# Impact of wildfire-related fine particulate matter on tuberculosis notifications in Brazil: a nationwide panel study, 2003–2023

**DOI:** 10.64898/2026.07.01.26356762

**Authors:** Thi Mui Pham, Thiago Mendonça, Yue Zhang, Derek Mallia, Julio Croda, Ted Cohen, Jason Andrews, Weeberb J. Réquia, Katharine S. Walter

## Abstract

Wildfire activity is intensifying with climate and land-use change, but its consequences for chronic infectious diseases remain poorly understood. We linked monthly tuberculosis notifications to wildfire-related fine particulate matter (PM_2.5)_ across 5,544 Brazilian municipalities (notifications 2005–2023; exposure 2003–2023), using a fixed-effects distributed-lag panel design that accounts for meteorology, diagnostic coverage, seasonality, and state-level trends. Fourteen additional high-exposure days (PM2.5>25 µg/m³) in a month were associated with an average increase of 2.7% [95% CI: 0.9–4.5%] in tuberculosis notifications in each of the following 24 months, peaking 11 months (IQR: 11–12) after exposure. Wildfire-related PM_2.5 e_xposure accounted for 2.0% [0.7–3.2%] of notifications in exposed municipality-months, with the largest attributable fractions in Pantanal (12.3%, 95% CI: 8.7–15.7%) and Amazônia (6.4%, 95% CI: 5.2–7.7%). These results suggest that wildfire-related PM_2.5 h_as a delayed effect on tuberculosis, extending health consequences of wildfire smoke beyond acute respiratory and cardiovascular outcomes and identifying a potential window for exposure-informed surveillance.

## Introduction

Tuberculosis (TB) remains the leading infectious disease cause of death globally, responsible for approximately 1.23 million deaths in 2024.^1^ Brazil carries the largest absolute TB burden in the World Health Organization (WHO) Region of the Americas, with roughly 84,000 new cases notified in 2025.^2^ Despite decades of investment in diagnosis and treatment, TB incidence in Brazil has plateaued, suggesting that conventional control strategies alone are insufficient and that upstream determinants of disease warrant greater attention.

Ambient air pollution is a near-universal exposure and major environmental determinant of health, with recent global estimates indicating that 99% of the entire world population is exposed to fine particulate matter (PM) concentrations exceeding WHO guideline levels.^3^ PM_2.5 c_ontributes substantially to cardiovascular and respiratory disease and premature mortality,^4,5^ and growing epidemiological and mechanistic evidence suggests that air pollution may also increase susceptibility to respiratory infections.^6–8^ For TB specifically, experimental evidence suggests that ambient PM_2.5 c_an compromise both innate and adaptive immune defences against *Mycobacterium tuberculosis* (*Mtb*), plausibly promoting progression to symptomatic disease.^9–13^ In studies of healthy Mexico City residents, higher in vivo PM burden in alveolar macrophages was associated with suppressed IL-1β and TNF-α production in bronchoalveolar cells in response to *Mtb* infection, and reduced IFN-γ production in peripheral blood mononuclear cells stimulated with mycobacterial antigens.^9,11^ These findings suggest that chronic inhalation of urban air pollution impairs both local lung and systemic antimycobacterial immune responses through PM-induced functional defects in alveolar macrophages. These mechanistic studies rely on primary human cells and cross-sectional exposure assessment and controlled in vivo evidence directly linking PM_2.5 e_xposure to accelerated TB progression in humans remains lacking.

Despite this biological plausibility, the epidemiological evidence base linking ambient PM_2.5 t_o TB remains sparse and constrained by study design limitations.^14–16^ A 2019 systematic review identified only 11 eligible studies, with inconsistent findings across pollutants and settings, although PM_2.5 w_as the pollutant most often linked to active TB.^14^ A subsequent meta-analysis found long-term PM_10,_ SO_2,_ and NO_2 a_ssociated with TB incidence, but no significant PM_2.5 a_ssociation.^8^ A 2022 review pooled 24 studies and found PM_2.5 a_ssociated with pulmonary TB (PTB) incidence, but not hospital admission or mortality, and graded the overall quality of evidence as low due to high risk of bias.^15^ A 2025 meta-analysis extended that pattern and reported a 26% higher PTB risk associated with long-term PM_2.5 e_xposure.^16^ Most studies were conducted in single cities, states, or provinces, predominantly from mainland China, with contributions from South Korea, Taiwan, and California.

Estimating the impact of air pollution on TB with epidemiological study designs is challenging because air pollution exposure shares social, environmental, and health-system factors that also shape TB risk, and because the lag from infection to TB disease onset is long and variable, spanning months to years.^17^ Case-crossover studies can reduce time-invariant confounding by using each individual as their own control,^18^ but the prolonged and variable latency of TB complicates the definition of the hazard and control periods. Moreover, such designs estimate the effect at a single pre-specified lag rather than flexibly estimating the timing of the exposure’s effect on the disease. Panel regression models have increasingly been used in climate and environmental health research to leverage within-location temporal variation in exposure while controlling for time-invariant differences between places, shared temporal shocks, and seasonality.^19^ Recent applications to infectious diseases have used panel designs to estimate delayed exposure-response relationships for outcomes including Lyme disease and dengue.^20,21^

Here, we aimed to estimate the population-level effect of wildfire-related air pollution on TB notification rates in Brazil between 2003 and 2023. We linked national TB notification data from 5,544 municipalities with source-specific PM_2.5 e_stimates from GEOS-Chem simulations. We used fixed-effects panel regression to leverage within-municipality changes in wildfire-related PM_2.5 o_ver time, while accounting for systematic differences between municipalities, state-level temporal trends, seasonality, and potential time-varying confounders. To capture delays consistent with TB progression and diagnosis, we estimated distributed lag effects of wildfire-related PM_2.5 o_ver the 1-24 months before notification using penalized spline smoothing. By combining within-municipality temporal variation, biologically motivated lag structures, and robustness analyses for key sources of bias, this design aims to support causal interpretation of the estimated effect between wildfire-related PM_2.5 a_nd TB notifications.

## Results

From Jan 1, 2005 to Dec 1, 2023, 1,504,346 TB cases (new and recurrent) were notified across 5,531 Brazilian municipalities (Table 1). The national mean annual notification rate was 39.5 per 100,000 population, with pronounced geographic heterogeneity: rates were highest in Amazonas (76.4 per 100k), Rio de Janeiro (75.7 per 100k), and Acre (54.0 per 100k), and lowest in Tocantins (13.6 per 100k), Goiás (14.6 per 100k), and the Federal District (16.2 per 100k) (Figure 1).

**Figure 1.**
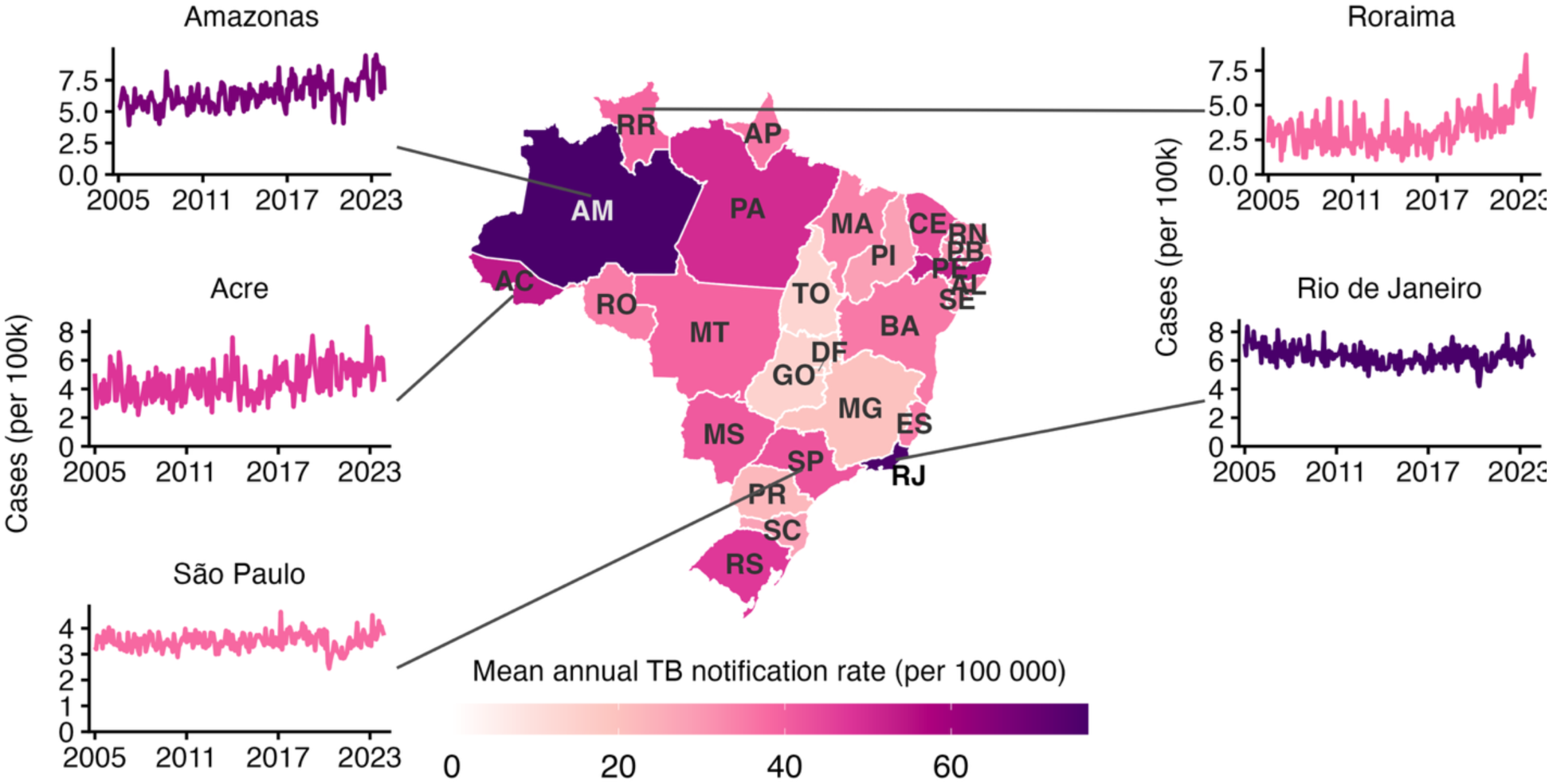
Spatiotemporal patterns of tuberculosis notification rates across Brazil, Jan 1, 2005–Dec 1, 2023. Map shows the average tuberculosis (TB) notification rate per 100,000 people over the study period for each Brazilian state and the Federal District. Insets display monthly TB notification rates for selected states representing distinct epidemiological settings: Amazonas and Roraima (high-burden northern states with substantial temporal variability), Acre (northern state with pronounced seasonality), Rio de Janeiro (consistently elevated urban burden), and São Paulo (moderate stable notification rates). Rates were calculated using population denominators from the Brazilian Institute of Geography and Statistics.^35^ Two-letter state abbreviations used in the map are defined in Supplementary Table 1.

**Table 1.**
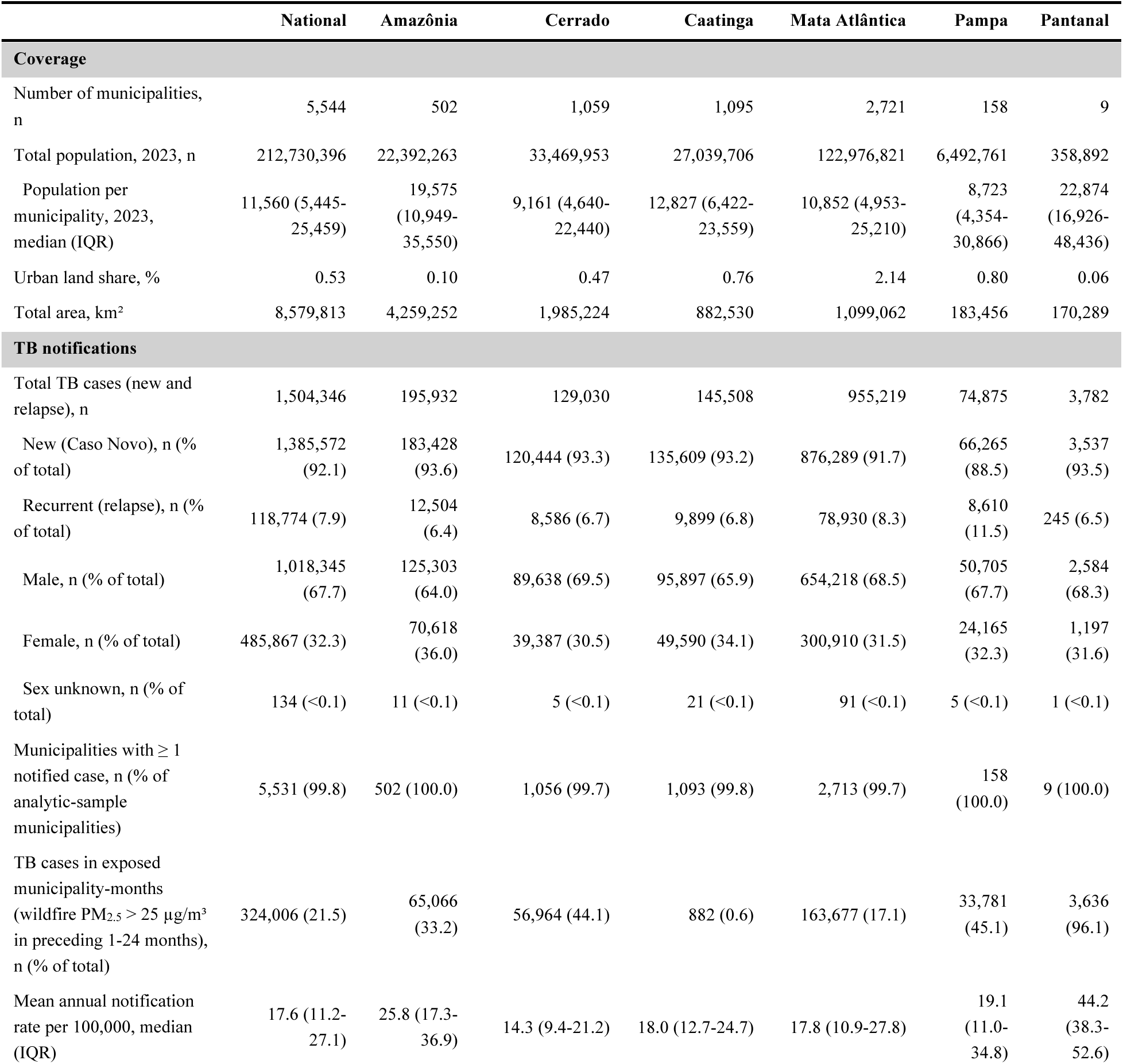
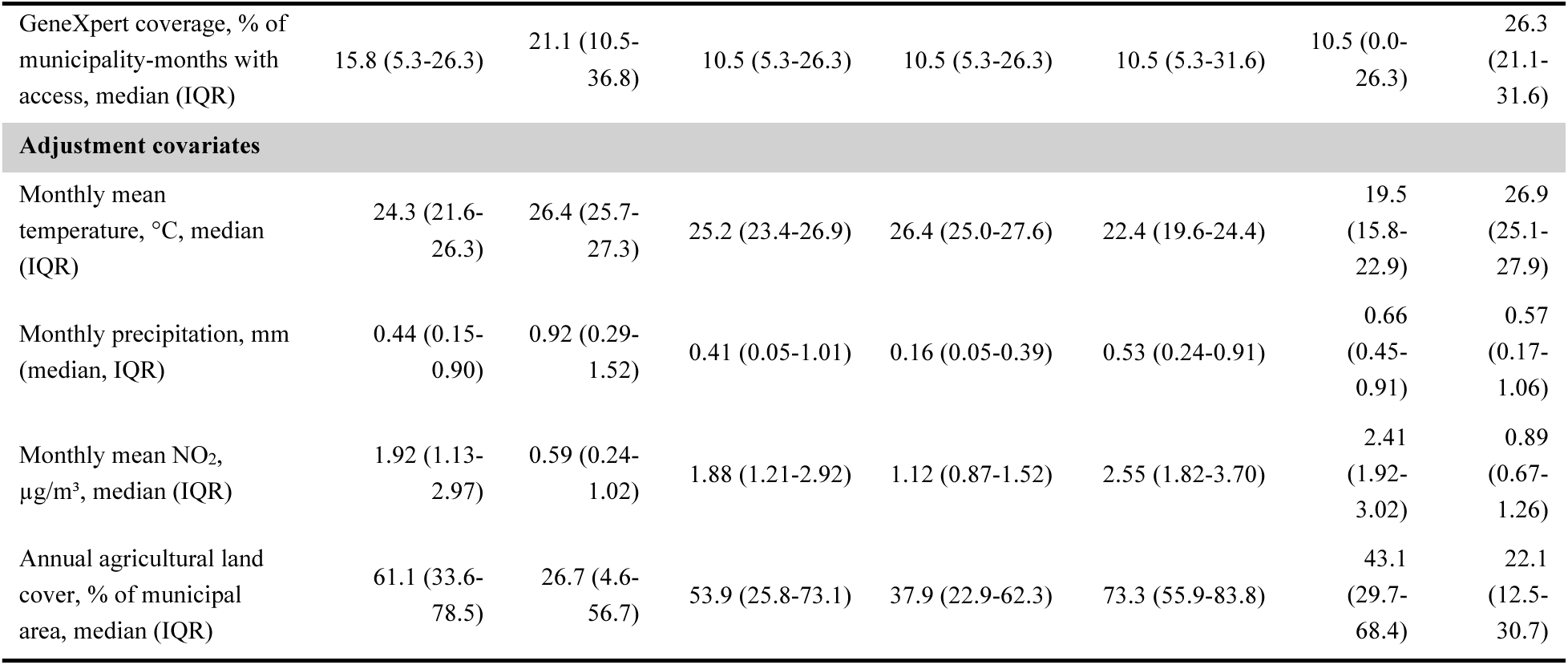
Descriptive statistics of TB notification and PM2.5 exposure data in Brazil, Jan 1, 2005 - Dec 1, 2023. Summary statistics describe the analytic cohort of municipalities with non-missing covariates over the outcome period, Jan 1, 2005 - Dec 1, 2023. Wildfire-related PM2.5 data from Jan 1, 2003 - Dec 1, 2023 are used, so that every outcome month has a complete 24-month exposure history. TB cases comprise new cases and recurrent cases (relapse); re-entries after loss to follow-up, transfers, post-mortem diagnoses and records with an unknown type of entry are excluded. Sex is as recorded in SINAN (male, female, or unknown). Column counts and totals (number of municipalities, total population in 2023, total area, total TB cases) are biome-level aggregates. The National column of the TB rows additionally includes notifications with an unknown or invalid municipality code, which cannot be assigned to a biome; the biome columns therefore sum to less than National. Municipalities with at least one notified case are expressed as a percentage of all Brazilian municipalities. Exposed-case counts are notifications occurring in exposed municipality-months, defined as municipality-months whose preceding 1 to 24 months contained at least one wildfire-PM2.5 day above the stated threshold. Urban land share is the area-weighted aggregate (total urban area divided by total municipal area), using each municipality’s most recent non-missing observation. Per-municipality summaries, i.e., population in 2023, mean annual TB notification rate per 100,000 (computed per municipality across years, then summarized across municipalities), GeneXpert coverage (% of municipality-months with prop_geneXpert > 0), and mean agricultural land cover, are reported as median (IQR) across municipalities within each biome. Monthly mean temperature, monthly precipitation, and monthly mean NO2 are summarized as median (IQR) across all municipality-months in the study period.

Wildfire-related PM_2.5 c_oncentrations showed strong spatial and seasonal patterns (Supplementary Fig. 3). Concentrations were highest in states along the arc of deforestation, encompassing Rondônia, northern Mato Grosso, Acre, Tocantins, southern Amazonas, and Pará. In these states, the average annual 90th percentile of daily PM_2.5 r_eached 28.3, 21.2, 15.2, 11.9, 7.6, and 5.3 µg/m³ (Supplementary Fig. 3). These same regions accumulated the greatest number of high-exposure days: municipalities in these states averaged 13.8 days per year above 25 µg/m³, compared with fewer than 1 day per year elsewhere (Figure 2A). Nationally, 20.4% of municipality-years recorded at least one day above 25 µg/m³. Wildfire-related PM_2.5 e_xposure was unevenly distributed across Brazil’s biomes (Figure 2B). Among municipality-months with at least one high-exposure day, Amazônia and Pantanal experienced the highest wildfire-related PM_2.5 e_xposure, with nearly half falling in the 21-30 high-exposure-day category (49% and 47%, respectively). In Amazônia, an additional 11% of exposed municipality-months exceeded 30 high-exposure days. By contrast, exposure in Mata Atlântica and Pampa were concentrated in the lowest category, with 1-5 high-exposure days accounting for 64% and 84% of exposed municipality-months, respectively, and almost no exposed months exceeding 21 days. Cerrado and Caatinga occupied an intermediate pattern, with exposure spread across both low- and high-exposure categories.

**Figure 2.**
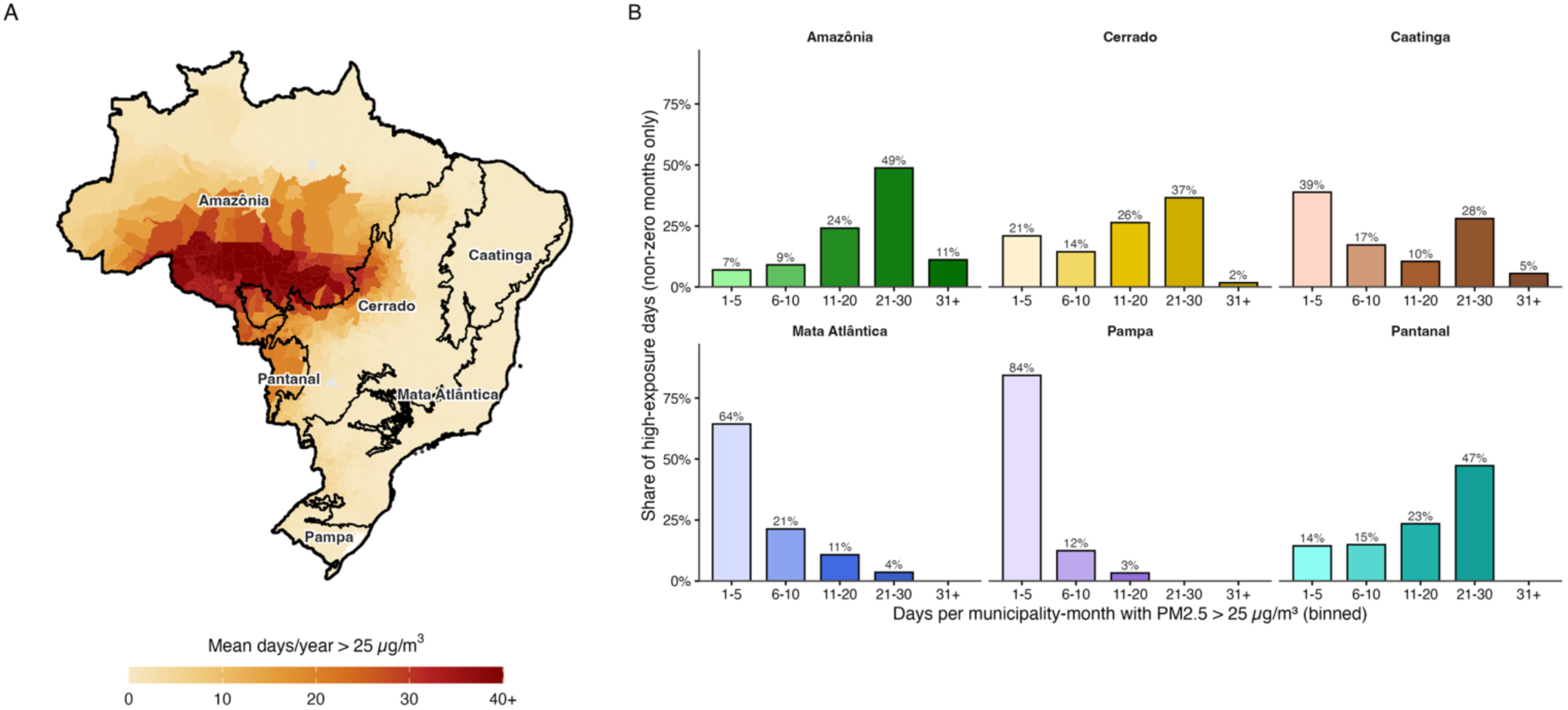
Spatial patterns of wildfire-related PM2.5 exposure in Brazil, Jan 1, 2003 – Dec 31, 2023. (A) Municipality-level choropleth of the mean number of days per year with wildfire-related PM2.5 above the primary threshold of 25 µg/m³. For each municipality, monthly exceedance counts were summed to annual totals and averaged across years. The upper limit is the 99th percentile of municipality values (rounded up to the nearest 10); municipalities above this cap are rendered in the top colour and the top legend tick is annotated with “+”. Biome boundaries (black, IBGE classification) are overlaid and labelled. (B) Distribution of binned high-exposure days per municipality-month, restricted to non-zero months and stratified by biome. Bars show the percentage of high-exposure days falling into each bin (1-5, 6-10, 11-20, 21-30, >=31 days/month above 25 µg/m³). Bars are coloured by the canonical biome palette (one colour family per biome, intensity increasing with the exceedance-day bin).

Over the whole study period, 324,006 (21.5%) TB cases had at least one high-exposure day (PM_2.5 >_ 25 µg/m³) 1-24 months prior to notification. In our primary analysis (>25 µg/m³), exposure to wildfire-related PM_2.5 i_ncreased TB notifications (Figure 3) such that 14 additional high-exposure days in a given month were associated with an average increase of 2.7% (95% CI: 0.9-4.5%) in TB notification rates in each of the following 24 months (Figure 3A). When we restricted exposures to those six to eighteen months prior to notification, the same exposure was associated with a mean monthly TB notification rate increase of 3.4% (95% CI: 1.7–5.2%; Figure 3A). The lag-response relationship was unimodal, with effects peaking at 11 months (IQR: 11-12) after exposure (Figure 3B, Supplementary Figs. 19 and 20). The estimated effect was smaller at a lower threshold of 15 µg/m³, where the 95% confidence interval included the null (1.5%, 95% CI: −0.2–3.1%; Supplementary Figs. 19 and 20), and larger at a higher threshold of 35 µg/m³ (3.7%, 95% CI: 1.2–6.4%; Supplementary Figs. 19 and 20).

**Figure 3.**
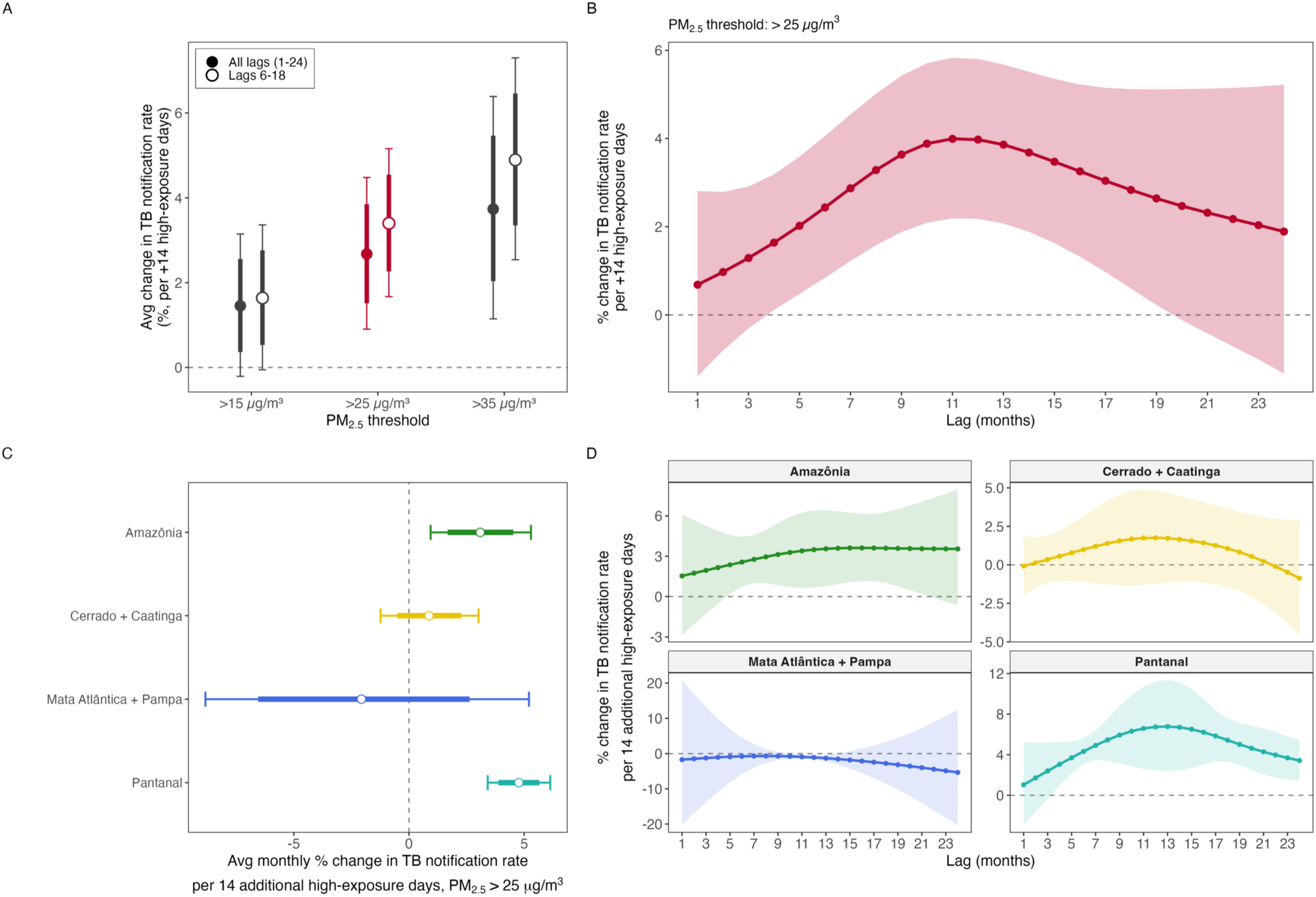
Lag-response relationship between wildfire-related PM2.5 exposure and TB notification rates, across high-exposure thresholds (A, B) and Brazilian biome groups (C, D), Jan 1, 2005 – Dec 1, 2023. (A) Average monthly percent change in TB notification rates per additional 14 high-exposure days, by PM2.5 high-exposure threshold (>15, >25, >35 µg/m³). Points are the geometric mean of per-month rate ratios across the lag window, expressed as percent change; thick bars show the 80% confidence interval and thin bars the 95% confidence interval, both from Conley spatial HAC standard errors (300 km cutoff). Filled circles use the full lag range (lags 1-24 months); open circles restrict the geometric mean to lags 6-18 months. The primary threshold (>25 µg/m³) is drawn in dark pink; the >15 and >35 µg/m³ sensitivity thresholds are drawn in dark grey. The dashed horizontal line indicates no effect. (B) Lag-response curve for the primary threshold (>25 µg/m³). The solid line shows the mean percent change in TB notification rates per +14 additional high-exposure days at each lag, with the shaded 95% confidence interval. The dashed line indicates no effect. (C) Forest plot of biome-stratified average monthly percent change in TB notification rate per 14 additional high-exposure days (PM2.5 > 25 µg/m³), reported as the geometric mean of lag-specific effects across lags 1-24. Points are point estimates; thick bars are 80% and thin bars 95% confidence intervals. Biomes were grouped a priori into Amazônia, Cerrado + Caatinga, Mata Atlântica + Pampa, Pantanal. (D) Lag-response curves by biome group, from the same biome-stratified penalized Poisson fixed-effect model as (C). Solid lines are mean estimates; shaded bands are 95% confidence intervals. Lag is shown in months from exposure (1-24).

In secondary age-stratified analyses, the association was strongest among adults aged 60 years or older and weakest among working-age adults, with evidence of heterogeneity across age groups (omnibus Wald p<0.0001; Supplementary Fig. 45).

We additionally conducted a concentration-based sensitivity analysis in which we defined exposures as the monthly 90th percentile of PM_2.5 c_oncentration rather than high-exposure days. The cumulative exposure–response relationship between wildfire-related PM_2.5 a_nd TB notifications was non-linear and convex, steepening at higher concentrations (Supplementary Figs. 16–18); the same 10 µg/m³ increase was associated with a 20% higher cumulative relative risk from a baseline of 15 µg/m³ (1.20, 95% CI 1.11–1.29) but a 72% higher risk from 25 µg/m³ (1.72, 1.42–2.08).

We predicted that the impact of wildfire-related PM_2.5 m_ight be greatest in biomes most impacted by fires. In a sub-group analysis, we found that effect magnitudes tracked exposure intensity more closely than case burden (Figure 3C,D). The average monthly effect per 14 additional high-exposure days was 3.1% (95% CI: 0.9–5.3%) in Amazônia and 4.8% (3.4–6.1%) in Pantanal. Point estimates for Cerrado + Caatinga were positive but overlapped the null, whereas the Mata Atlântica + Pampa stratum, which includes the cities Rio de Janeiro and São Paulo, contributed the largest share of TB cases but had the lowest wildfire exposure, showed little evidence of an association and wide confidence intervals.

We next quantified the static attributable fraction of TB notifications associated with wildfire-related PM_2.5 a_mong exposed municipality-months (Figure 4). The nationwide average AFE was 2.0% (95% CI 0.7–3.2%), corresponding 6,386 (95% CI: 2,263, 10,264) attributable cases in exposed municipalities. The AFE varied over time (Figure 4A), from 2.9% (95% CI: 1.1–4.6%) in 2005 to a low of 0.9% (95% CI: 0.0–1.8%) in 2010, rising to 3.8% (95% CI: 1.3–6.2) in 2022, although the year-specific confidence intervals were wide and overlapping. The AFE was highly heterogeneous across space (Figure 4B-C), with the highest fractions in the Pantanal (12.3%, 95% CI: 8.7–15.7%) and Amazônia (6.4%, 95% CI: 5.2–7.7%). Across microregions, the AFE ranged from 0.1% to 14.8%. Despite a lower AFE, the large case burden in Amazônia meant that most wildfire-related TB cases nationally occur in this biome, accounting for 4,187 (95% CI: 3,350–4,993) attributable cases, whereas the high AFE in the Pantanal translated into relatively few attributable cases of 446 (95%CI: 315–571) given its small population (Figure 4D).

**Figure 4.**
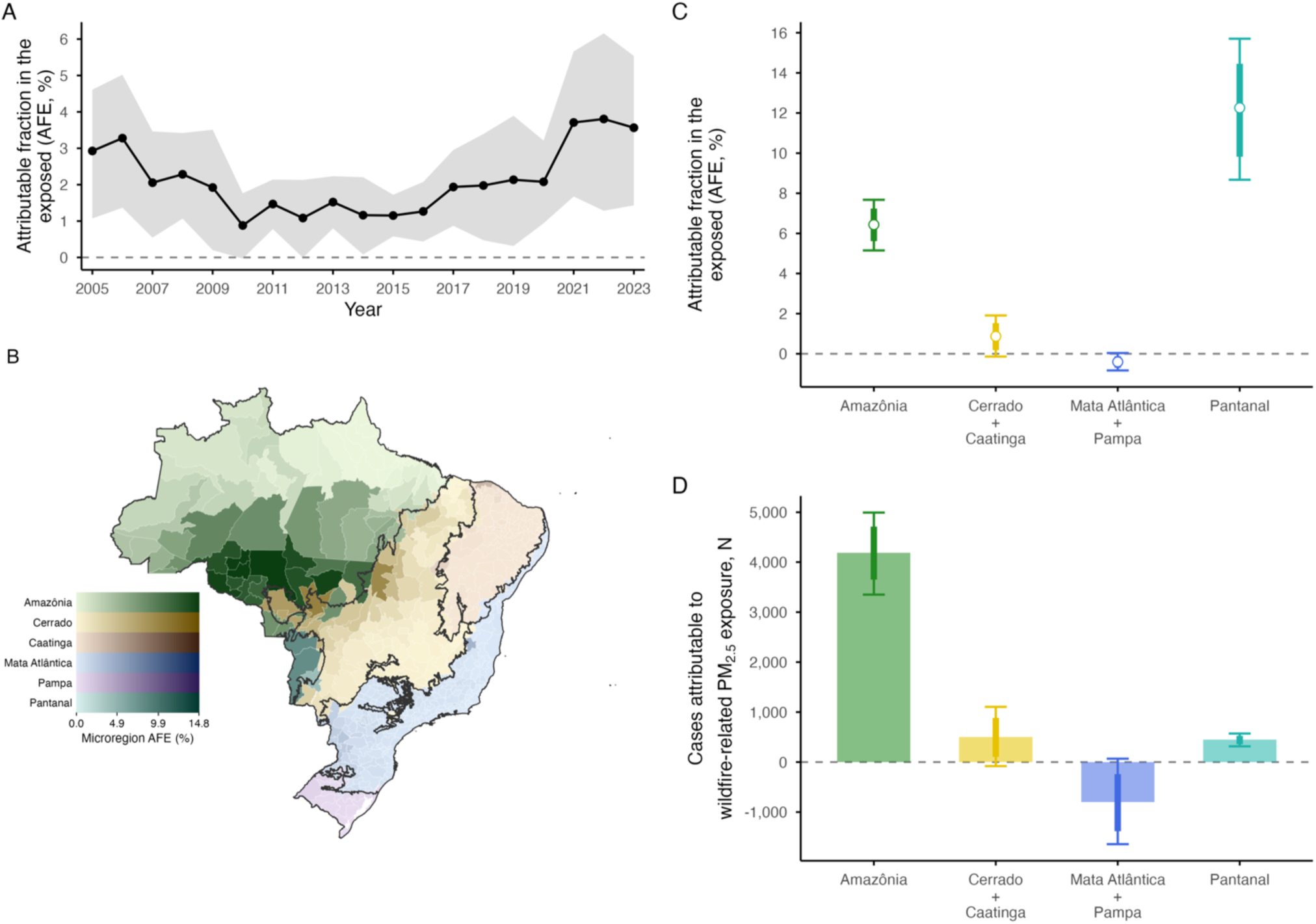
Attributable fraction in the exposed (AFE) of tuberculosis notifications to wildfire-related PM2.5 exposure above 25 µg/m³, Brazil, Jan 1, 2005 – Dec 1, 2023. (A) AFE by calendar year, estimated from the penalized spline distributed-lag model with municipality and state×year, state×month, state-by-pandemic-phase fixed effects and a log-population offset; line and points show the point estimate, the grey ribbon the 95% confidence interval based on Conley HAC standard errors (300 km cutoff). The dashed line marks AFE = 0. (B) Choropleth of AFE (%) by IBGE microregion. Each microregion is assigned to its dominant biome by largest-overlap spatial join and shaded on a biome-specific sequential ramp: colour encodes biome (Amazônia, Cerrado, Caatinga, Mata Atlântica, Pampa, Pantanal) and lightness encodes AFE magnitude, with all ramps sharing identical AFE limits so equivalent shading reflects equivalent AFE across biomes. Microregions never exposed above the 25 µg/m³ threshold are shown at the pale end of their biome ramp. Black outlines delineate biome boundaries. (C) AFE by biome from the biome-interaction model with merged groups (Cerrado + Caatinga, Mata Atlântica + Pampa); points show the point estimate, thick segments the 80% confidence interval, and thin whiskers the 95% confidence interval. (D) Number of TB cases attributable to wildfire-related PM2.5 exposure by biome from the same biome-stratified model; bars show point estimates, thick segments the 80% confidence interval, and thin whiskers the 95% confidence interval.

Our findings were robust across various sensitivity analyses. These included adjustment for NO₂ and O3 as a co-pollutants, agricultural expansion, drought index, and bacteriological confirmation (Supplementary Figs. 25–28, 31 and 32). We accounted for residual seasonality through state-by-month fixed effects and two additional tests. First, we extended the lag window to 32 months. If the estimated peak were due to residual seasonality, it would recur at the next annual lag at 24 months. We found no recurrence of the association at 24 months (Supplementary Section S3.10, Supplementary Fig. 34). Second, we replaced the exposure with its municipality-specific de-seasonalized exposure, i.e., the number of high-exposure days in a given month minus the municipality’s average for that calendar month over the study period 2003-2023. Although this approach removed 47% of the exposure variance, it left the association largely unchanged (average monthly increase 2.7% [95% CI: 0.8–4.6%] vs 2.7% [0.9–4.5%]), and the median peak remained at 11 months. Because migration from Venezuela into Brazil’s northern border states increased sharply after 2015,^22^ we also excluded TB cases recorded as immigrants, all municipalities in Roraima, or twin cities on the international border. None of these exclusions materially changed the estimates (average monthly increase: 2.7% [1.0–4.5%], 2.7% [0.9–4.5%] and 2.4% [0.2–4.7%], respectively; median peak 11–12 months; Supplementary Section S3.16, Supplementary Fig. 43). Adjustment for burned area within the municipality did not change the association (2.8% [1.0–4.6%]; Supplementary Section S3.15.1, Supplementary Fig. 40). Wildfire-related PM_2.5 w_as not associated with hospitalisations for malnutrition (−1.0% [−4.6% to 2.8%]; Supplementary Section S3.15.2, Supplementary Fig. 41), used here as a marker of fire-related food insecurity. A negative-control exposure, distant wildfire-related PM_2.5,_ drawn from a randomly selected municipality at least 1500 km from the focal municipality, was not associated with TB notification rate (Supplementary Fig. 37). Negative-control outcomes (leprosy, appendicitis) showed no association with wildfire-related PM_2.5 (_Supplementary Figs. 38 and 39).

## Discussion

In this nationwide panel study, high wildfire-related PM_2.5 e_xposure increased TB notifications, with effects peaking 11 months after exposure, a substantially longer lag than typically considered for acute respiratory outcomes. The attributable burden was concentrated along Brazil’s arc of deforestation, where recurrent dry-season smoke exposure overlaps with populations already at elevated risk of TB. Our findings identify wildfire smoke exposure as a previously under-recognized and modifiable environmental risk factor for TB in Brazil.

TB notification rates in Brazil have stagnated over the past decade despite sustained investment in the National Tuberculosis Control Programme, underscoring the importance of identifying modifiable upstream drivers of disease. Our findings point to wildfire-related air pollution as one such driver. The concentration of attributable burden in fire-prone regions and the delayed lag structure have direct implications for TB control. The 11-month peak suggests a predictable window for intensified TB surveillance, contact tracing, and preventive therapy in affected regions during the year following severe fire seasons. These efforts could be supported by the operational smoke forecasts already produced by Brazil’s National Institute for Space Research (INPE). More broadly, our results suggest that wildfire-related air pollution should be considered alongside established TB risk factors, including tobacco smoking, HIV, diabetes, and undernutrition, to guide prevention and targeting efforts.^23^ Our findings also point to upstream intervention opportunities: policies that influence wildfire activity, including deforestation control and regulation of agricultural burning, act directly on this risk pathway but are not currently integrated into TB control strategies.^24,25^ As climate change increases the frequency and severity of wildfires,^26,27^ this pathway is likely to become increasingly important not only in Brazil but in fire-prone regions globally with substantial latent TB reservoirs, including parts of sub-Saharan Africa and Southeast Asia.

The observed association between wildfire-derived PM_2.5 a_nd TB notifications could reflect increased susceptibility to new *Mtb* infections, transmission from individuals with active TB, progression from latent infection to active disease, or case detection. While analysis of TB notification rates alone cannot implicate a specific biological mechanism, a peak effect at 11 months prior to notification is consistent with either a PM_2.5-_associated increase in susceptibility to primary infection or progression to symptomatic disease. Although increased *Mtb* transmission during smoke episodes cannot be excluded,^18^ the larger effects observed in rural compared with urban municipalities, and in older adults are more consistent with a progression-based mechanism. This interpretation is supported by recent individual-level evidence linking long-term PM_2.5 e_xposure to increased risk of progression from latent TB infection to active disease.^28^ In addition, experimental studies demonstrated that PM_2.5 e_xposure impairs alveolar macrophage function, IFN-γ-responses, and granuloma maintenance disruptions – processes central to containing latent infection.^9–13^ Wildfire-specific PM_2.5 m_ay be particularly relevant because it has been associated with greater respiratory toxicity than PM_2.5 f_rom other sources.^29^ Detection biases due to increased respiratory symptoms are unlikely to explain our findings, because such bias would be expected to increase TB notifications within weeks following fire exposures, whereas our estimates indicate a delayed association emerging over several months and persisting across longer lags.

The estimated attributable burden of TB to wildfire exposures was highly variable across space. The attributable fraction reached up to 15% in exposed Amazônia and Pantanal micro-regions, where fire activity produces frequent and sustained smoke exposure during dry seasons. This pattern highlights potential gaps in current TB control policies. TB control efforts in Brazil are guided by priority municipalities defined by disease burden, social vulnerability, and high-risk populations, many of which are large urban municipalities and state capitals where a substantial share of TB notifications occur.^30,31^ In contrast, wildfire-related PM_2.5 a_re concentrated in fire-prone northern and central-western regions characterized by more dispersed rural and Indigenous populations, larger geographic distances and weaker healthcare infrastructures.^32^ The same populations therefore face both elevated environmental exposure and reduced programmatic reach.

Our study has several strengths. We linked national TB surveillance data to source-specific wildfire-related PM_2.5 e_stimates across more than 5,500 municipalities over two decades, providing broad spatial and temporal coverage to assess an environmental determinant of TB. The panel fixed-effects design exploits within-municipality variation in PM_2.5 w_hile controlling for time-invariant municipal characteristics, state-level long-term trends, and seasonal patterns. The distributed lag framework flexibly estimates the timing and duration of effects without imposing a pre-specified fixed lag. Finally, our results were consistent across a comprehensive set of sensitivity analyses, including alternative PM_2.5 t_hresholds, an independent fire-emissions inventory, and adjustment for co-pollutants and land use. The fixed-effects design, the delayed lag structure and these analyses support a causal interpretation of the estimates at the population level, under the assumption of no unmeasured time-varying confounding within municipalities. Negative-control analyses were consistent with this assumption: they did not detect residual bias from shared surveillance or health-system shocks (leprosy, appendicitis) or from spatially structured confounding (distant-municipality exposure).

Our results should be interpreted in light of several limitations. First, as an ecological analysis, our study is subject to the ecological fallacy: because both exposure and outcome were analysed at the municipality level, we cannot determine whether individuals notified with TB were those exposed to wildfire-related PM_2.5._ Our estimates should therefore be interpreted as population-level rather than individual-level effects. Second, although our fixed-effects design and extensive sensitivity analyses substantially reduce confounding, we cannot exclude residual bias from unmeasured time-varying factors associated with fire activity and independently influence TB risk. Fires may affect TB through pathways other than smoke inhalation, including loss of livelihood among workers affected by fires and disruption of local food systems leading to undernutrition, an established risk factor for TB.^33^ In sensitivity analyses, we adjusted for agricultural land use and drought, and found no association between wildfire-related PM₂.₅ and malnutrition hospitalisations. However, these hospitalisations capture only severe undernutrition, so we cannot fully exclude a contribution of these pathways to the observed association. Third, our analysis assumes that municipal populations were broadly stable over the lag window. Our estimates were robust to excluding immigrant cases and border municipalities, but movement within Brazil could not be assessed, as SINAN does not record short-term residential history. Any resulting exposure misclassification would most likely bias our estimates towards the null. Fourth, SINAN captures diagnosed cases rather than true incidence, and our estimates therefore reflect the combined effects of wildfire-related PM_2.5 o_n TB diagnosis and case detection. Although differential diagnosis could theoretically bias our estimates, several findings make this explanation less likely, including the persistence of results when restricted to microbiologically confirmed cases and the delayed lag structure. Fifth, wildfire-related PM_2.5 c_oncentrations were estimated using atmospheric modelling rather than direct measurements. Any resulting exposure misclassification is likely to be non-differential with respect to TB notifications and would therefore be expected to attenuate the estimated effects. Our sensitivity analysis using QFED2.5 as an alternative fire emissions inventory yielded a similar lag-response pattern, albeit with smaller effect sizes. Lastly, our analysis does not explicitly model dynamic spatial dependence. Although Conley standard errors account for spatial correlation in uncertainty, they do not capture spillover mechanisms such as cross-municipality TB transmission. Attributable-burden estimates should therefore be interpreted as static municipality-level quantities.

Together, our findings extend the health consequences of wildfire smoke beyond acute respiratory and cardiovascular outcomes and show that the effects on infectious disease burden can extend over months and years rather than days or weeks. As wildfire activity intensifies globally, understanding these delayed effects will be increasingly important for anticipating and preventing the full health burden of climate-driven environmental change.

## Methods

### Study design and setting

We conducted a nationwide panel study of 5544 Brazilian municipalities with the aim to estimate the population-level effect of wildfire-related PM_2.5 e_xposure on TB notification rates across Brazil between Jan 1, 2005 and Dec 31, 2023. Leveraging spatiotemporal variation in wildfire-related PM_2.5 e_xposure, our design approximates a natural experiment, and we interpret the results as supporting a population-level causal interpretation, conditional on fixed effects and time-varying covariates described below.

### Data sources

#### TB notification data

Individual-level TB notification records (new and recurrent) from Jan 1, 2005 to Dec 31, 2023 were obtained from the Brazilian Notifiable Disease Information System (SINAN), which compiles compulsory TB notifications from all healthcare facilities nationwide.^34^ Records were aggregated to municipality-month counts using the date of notification and the municipality of residence. Age, sex, HIV co-infection status, and method of laboratory confirmation were extracted for stratified analyses.

#### Population denominators

Population denominators were obtained from annual intercensal estimates produced by the Brazilian Institute of Geography and Statistics and assigned to all months within each year.^35^

#### Source-specific PM_2.5 d_ata

Daily wildfire-related PM_2.5 c_oncentrations at 0.25°× 0.25° resolution were obtained from a global source-specific PM_2.5 d_ataset based on GEOS-Chem chemical transport model simulations for Jan 1, 2003 to Dec 31, 2023.^36^ In this dataset, fire-attributable PM_2.5 w_as estimated from paired GEOS-Chem simulations with and without biomass-burning emissions; the resulting fire- to-total PM_2.5 r_atio was then applied to machine-learning bias-corrected total PM_2.5 c_oncentrations (Supplementary Methods S1.4). Estimates include transboundary smoke from fires outside Brazil. We use the term “wildfire-related PM_2.5”_ to refer to PM_2.5 a_ttributed to biomass-combustion fire emissions, including natural wildfires, deforestation fires, and agricultural burning, consistent with terminology used in previous epidemiological studies of fire-attributable PM_2.5._^37^ Our primary analysis used the Global Fire Emissions Database version 4.1 with small fires (GFED4.1s) which derives emissions from satellite-observed burned areas combined with biogeochemical modelling of fuel consumption. Concentrations were spatially aggregated to municipalities using area-weighted zonal statistics. Sensitivity analyses used the Quick Fire Emissions Dataset (QFED2.5), which derives fire emission estimates from satellite-derived fire radiative power.

#### Ecological biomes

Each municipality was assigned to one of Brazil’s six terrestrial biomes (Amazônia, Cerrado, Caatinga, Mata Atlântica, Pampa, Pantanal), based on 2019 definition of the Brazilian Institute of Geography and Statistics (IBGE) and on majority area overlap.^38^ IBGE draws biome boundaries mainly from dominant vegetation, climate, relief, and geology. For effect-modification analyses, biomes were grouped a priori according to wildfire-related PM_2.5 e_xposure distributions and sample size considerations into three strata: Amazônia; Cerrado and Caatinga; Pantanal; Mata Atlântica and Pampa. Caatinga was grouped with Cerrado because it had too little within-municipality variation in exposure to be analysed alone (Supplementary Fig. 9D), and the two biomes share a savanna- and shrubland-dominated burned-area profile and a September–October exposure peak (Supplementary Fig. 9A,B). More information on the biomes can be found in the supplement (section S1.9).

#### Climate and land use data

Daily mean temperature and total precipitation were derived from the Copernicus ERA5-Land reanalysis and aggregated to municipality-month means using area-weighted zonal statistics.^39^ Nitrogen dioxide (NO_2)_ concentrations were obtained from the Copernicus Atmosphere Monitoring Service (CAMS) global reanalysis (EAC4) and included in co-pollutant sensitivity analyses.^40^ Annual municipal proportions and corresponding annual change rates of agricultural land were extracted from MapBiomas Collection 10.^41^

#### Outcome and exposure

The primary outcome was the monthly count of newly notified and recurrent active TB cases (all clinical forms), aggregated to the municipality of residence. For descriptive analyses, TB notification rates per 100,000 people were calculated at the state-level.

The primary exposure was the monthly count of high-exposure wildfire-related PM_2.5 d_ays per municipality, defined as days on which the municipality-level wildfire-related PM_2.5 c_oncentration exceeded 25 µg/m³. This threshold corresponds to the final 24-hour air quality target under Brazil’s national air quality framework.^42^ Alternative thresholds of >15 µg/m³ and >35 µg/m³ were examined in sensitivity analyses, corresponding to the World Health Organization 24-hour PM_2.5 g_uideline and the U.S. Environmental Protection Agency 24-hour PM_2.5 s_tandard, respectively.^43,44^ We included lagged exposure windows from 1–24 months prior to TB notification to capture delayed effects of wildfire-related PM_2.5 o_n TB progression.^17^

### Statistical analysis

We estimated the effect of high-exposure days on TB notifications using Poisson panel regression. Fixed effects for municipality, state-by-year, state-by-month, and state-by-pandemic phase controlled for time-invariant differences between municipalities, state-level long-term trends, seasonality, and COVID-19-related disruptions. We adjusted for time-varying covariates, including GeneXpert MTB/RIF diagnostic coverage (SINAN) and meteorological conditions (temperature and precipitation) at corresponding lag months. To characterize the lag-response relationship, we estimated a penalized distributed lag model (DLM) over lag months 1 to 24 with basis dimension k = 8 and second-order penalty (m = 2), where the smoothing parameter was estimated via fast restricted maximum likelihood (fREML).^45,46^

Effect estimates are reported as average monthly notification rate ratios per 14 additional high-exposure days in a single month. This measure is the geometric mean of the 24 lag-specific rate ratios, that is, the cumulative effect over the 1–24-month lag window re-expressed as a single constant per-month rate. Lag-specific rate ratios and pointwise 95% confidence intervals were recovered by projecting the basis coefficients onto the original lag grid; the average per-lag-month effect was reported as a summary measure. Standard errors and 95% confidence intervals were computed using the Conley spatial heteroskedasticity- and autocorrelation-consistent (HAC)^47^ estimator with a primary distance cutoff of 300 km. Additional cutoffs were examined in sensitivity analyses.

### Attributable burden

We estimated the burden of TB notifications attributable to wildfire-related PM_2.5 h_igh-exposure days using a backward attributable fraction framework adapted to distributed-lag models.^48^ For each municipality-month, we defined a counterfactual in which exposure was set to zero days above 25 µg/m³ over the preceding 24 months, while holding all other covariates and fixed effects at their observed values. Attributable fractions were derived by evaluating the fitted cross-basis at the observed and counterfactual exposure profiles and taking their difference.

Attributable case numbers were obtained by multiplying these fractions by observed counts. Our primary estimand was the attributable fraction in the exposed (AFE)^49^ defined as the attributable fraction among municipality-months with at least one high-exposure day in the 24-month lag window. Overall estimates were obtained by aggregating attributable cases and observed counts across all exposed observations; biome- and microregion-year estimates were computed analogously. Uncertainty was propagated by sampling from the posterior distribution of the cross-basis coefficients using the Conley spatial-HAC covariance matrix, recomputing attributable cases and AFE for each draw, and reporting 95% empirical intervals.

### Sensitivity analyses

We conducted pre-specified sensitivity analyses using alternative PM_2.5 t_hresholds, fire emission inventory (QFED2.5), NO2 co-pollutant adjustment (25 µg/m³ threshold), spatial HAC distance cutoff (500 km); and negative control exposure and outcome.^50^ Further details are provided in Supplementary Information sections S3–S5.

The funder had no role in study design, data collection, data analysis, data interpretation, writing of the manuscript, or the decision to submit for publication. All authors had full access to all data in the study and accept responsibility for the decision to submit for publication.

## Data Availability

All data produced in the present study are available upon reasonable request to the authors.

https://doi.org/10.5281/zenodo.15493914

https://datasus.saude.gov.br/

## Data availability

Wildfire-related fine-particulate-matter data are available from Zenodo (GFED4.1s: https://doi.org/10.5281/zenodo.15493914; QFED2.5: https://doi.org/10.5281/zenodo.15496596). Individual-level tuberculosis notification data are available from the Brazilian Ministry of Health (https://portalsinan.saude.gov.br). Population estimates and geographic boundaries are available from IBGE (https://www.ibge.gov.br); meteorological and atmospheric-composition data from the Copernicus Climate and Atmosphere Data Stores; and land-use data from MapBiomas Collection 10 (https://brasil.mapbiomas.org).

## Code availability

Code for data processing and statistical analyses is available at https://github.com/tm-pham/tb-wildfire-pm25.git.

## Acknowledgements

We thank the Brazilian Ministry of Health for providing public access to the health data used in this study. This work was supported by the National Institutes of Health (K01AI173385) and the University of Utah 3i Initiative. The funders had no role in study design, data collection, analysis, interpretation, manuscript preparation or the decision to submit the work for publication.

## Author contributions

KSW and TMP conceived the study. TMP and KSW designed the study. TMP and TM curated the data. TMP conducted the formal analysis. TMP, KSW, WJR, YZ, JC, JA, TC, and DM interpreted the findings. TMP drafted the manuscript. All authors revised the content of the manuscript critically and approved the final version. TMP, TM, WJR, and KSW directly accessed and verified the underlying data. All authors had full access to the data in the study and accept responsibility for the decision to submit for publication.

## Competing interests

The authors declare no competing interests.

## Notes

### Competing Interest Statement

The authors have declared no competing interest.

### Author Declarations

This study used ONLY openly available tuberculosis notification data from the Brazilian National Notifiable Diseases Information System (SINAN), available through the Brazilian Ministry of Health (DATASUS: https://datasus.saude.gov.br/). Wildfire-related PM2.5 exposure estimates were derived from publicly available GEOS-Chem simulations driven by the Global Fire Emissions Database version 4.1 (GFED4.1) (https://doi.org/10.5281/zenodo.15493914). All source data were publicly available before the initiation of the study. For this analysis, the individual-level records were de-identified prior to use and were aggregated to municipality-month counts by date of notification and municipality of residence. The primary analytic dataset, therefore, contained aggregated summary outcome data at the municipality-month level, not individually identifiable data.

### Summary of Updates

Updated analyses according to revised inclusion criteria and improved clarity of the figures. Added several additional sensitivity analyses.

