## Supplementary material for "Impact of wildfire-related fine particulate matter on tuberculosis notifications in Brazil: a nationwide panel study, 2003–2023"

#### Table of contents

S1. Data sources and processing ... 3
S1.1 SINAN tuberculosis notifications ... 3
S1.2 Population denominators (IBGE) ... 3
S1.3 Administrative boundaries and projection ... 3
S1.4 Wildfire-related $PM_{2.5}$ ... 5
S1.4.1 Data source ... 5
S1.4.2 Threshold-based high-exposure days ... 6
S1.4.3 Wildfire-related $PM_{2.5}$ event durations ... 8
S1.5 Meteorological data (ERA5-Land) ... 10
S1.6 Nitrogen dioxide co-pollutant (CAMS EAC4) ... 11
S1.7 Ozone co-pollutant (CAMS EAC4) ... 12
S1.8 Land use and land cover (MapBiomas Collection 10) ... 13
S1.9 Biome fire regimes and exposure characteristics ... 13
S1.10 Drought index ... 15
S1.11 Leprosy ... 17
S1.12 Appendicitis ... 17
S1.13 Malnutrition ... 18
S1.14 Spatial assignment of gridded exposures ... 19
S1.15 Lag variable construction ... 20
S1.16 GeneXpert diagnostic coverage ... 20
S1.17 Pandemic-phase definition ... 21
S2. Statistical methods ... 22
S2.1 Panel fixed-effects design and confounding control overview ... 22
S2.2 Fixed-effects specification ... 22
S2.3 Covariates ... 23
S2.4 Poisson regression ... 24
S2.5 Conley spatial HAC standard errors ... 25
S2.6 Penalized P-spline distributed lag model ... 25
S2.7 Backward attributable-fraction g-computation ... 25

S2.8 Concentration-based DLNM specification ... 26
S3. Sensitivity and robustness analyses ... 29
S3.1 Alternative $PM_{2.5}$ thresholds ... 29
S3.2 QFED2.5 alternative fire-emission inventory ... 31
S3.3 $NO_2$ co-pollutant adjustment ... 35
S3.4 $O_3$ co-pollutant adjustment ... 36
S3.5 Drought and SPEI adjustment ... 37
S3.6 Agricultural-expansion adjustment ... 38
S3.7 Microregion aggregation ... 39
S3.8 Bacteriologically confirmed cases only ... 40
S3.9 Sensitivity to adjustment for GeneXpert diagnostic ... 44
S3.10 Testing for residual seasonality by extending the lag window ... 45
S3.11 Testing for residual seasonality with a de-seasonalized exposure variable ... 46
S3.12 Conley distance-cutoff sensitivity ... 47
S3.13 Negative control exposure ... 48
S3.14 Negative control outcomes ... 50
S3.14.1 Leprosy ... 50
S3.14.2 Appendicitis ... 51
S3.15 Sensitivity analysis of other wildfire-related effects ... 52
S3.15.1 Local fire activity vs transported smoke ... 52
S3.15.2 Malnutrition hospitalisations as an alternative-pathway outcome ... 54
S3.16 Exclusion of migrant TB cases and border municipalities ... 55
S3.17 Consideration of additional time-varying confounders ... 57
S4. Supplementary heterogeneity results ... 58
S4.1 Effect modification by age group ... 58
S4.2 Effect modification by HIV status ... 61
S4.3 Effect modification by biological sex ... 62
S5. Model diagnostics ... 63
S5.1 QQ and residual-vs-predicted diagnostics ... 63
S5.2 Basis adequacy and smoothing parameter ... 64
S5.3 Residual Temporal Autocorrelation ... 65
S5.4 Spatial correlation residuals ... 66
S5.5 Residual calibration against population ... 67
S6. Reporting and reproducibility ... 69
S6.1 Data and code availability details ... 69
S6.2 Software versions ... 69

### S1. Data sources and processing

#### S1.1 SINAN tuberculosis notifications

Individual-level tuberculosis (TB) notification records were obtained from the Brazilian Notifiable Disease Information System (Sistema de Informação de Agravos de Notificação, SINAN), administered by the Brazilian Ministry of Health.[1] SINAN compiles compulsory notifications from all healthcare facilities nationwide and is the primary surveillance system for TB in Brazil. The system captures demographic, clinical, and diagnostic information for each notified case, including date of notification, municipality of residence, whether the patient was identified as an immigrant (foreign national living in Brazil for less than 2 years), age, sex, HIV co-infection status, and method of laboratory confirmation.

Notifications were aggregated to municipality-month counts by date of notification and municipality of residence. Municipality-months with no notifications were assigned a count of zero. The analysis dataset includes 1,504,513 TB cases (new and recurrent) across 5,544 municipalities for the period January 1, 2005 to December 1, 2023.

We use the term 'notification rate' because SINAN captures diagnosed and reported cases rather than true incidence. A spatial-mechanistic analysis using routine SINAN notifications and mortality data for 2016–2018 estimated that 86% (uncertainty interval: 82–89%) of incident TB cases in Brazil received diagnosis and treatment, with substantial municipal heterogeneity ranging from 73% to 95%.[2] Differential detection (where wildfire exposure itself might alter the probability of diagnosis) is addressed in Sections S2.1 and S3.8.

#### S1.2 Population denominators (IBGE)

Annual municipal population estimates were obtained from the Brazilian Institute of Geography and Statistics (Instituto Brasileiro de Geografia e Estatística, IBGE).[3] IBGE produces annual intercensal estimates using demographic methods anchored to decennial censuses. Each annual estimate was applied to all twelve months of the corresponding calendar year, so that monthly population denominators within a year were assumed constant.

#### S1.3 Administrative boundaries and projection

Municipality and microregion boundaries were obtained from the geobr R package, which serves IBGE official cartographic boundaries. Municipalities nest within Brazil's 27 federative units (26 states and the Federal District), which IBGE groups into five macro-regions (North, Northeast, Central-West, Southeast, and South); the two-letter state abbreviations used throughout the figures and tables of this manuscript are defined in Supplementary Table 1. Each municipality was assigned to one of Brazil's six IBGE terrestrial biomes (Amazônia, Cerrado, Caatinga, Mata Atlântica, Pampa, Pantanal) based on majority area overlap (Supplementary Fig. 1). For analyses requiring distance computations, all spatial operations used the SIRGAS 2000 / Brazil Polyconic projected coordinate reference system (EPSG:5880).

Municipalities in the border strip (*faixa de fronteira*), the 150 km zone along Brazil's international land border, and the subset of twin cities (cidades gêmeas) were identified from the 2022 IBGE list (Supplementary Fig. 42A). The list includes 588 border-strip municipalities, 33 of which are twin cities. All municipalities of Roraima lie within the border strip.

**Supplementary Table 1. Abbreviations and full names of the 27 Brazilian federative units (states and the Federal District).** Two-letter abbreviations are the official IBGE codes used throughout the figures and tables of this manuscript. States are grouped by IBGE macro-region (North, Northeast, Central-West, Southeast, South) and ordered approximately North to South within each region; the Federal District (DF) is listed under the Central-West region.

| Abbreviation | State |
|---|---|
| **North** | |
| AC | Acre |
| AM | Amazonas |
| RR | Roraima |
| RO | Rondônia |
| PA | Pará |
| AP | Amapá |
| TO | Tocantins |
| **Northeast** | |
| MA | Maranhão |
| PI | Piauí |
| CE | Ceará |
| RN | Rio Grande do Norte |
| PB | Paraíba |
| PE | Pernambuco |
| AL | Alagoas |
| SE | Sergipe |
| BA | Bahia |
| **Central-West** | |
| MT | Mato Grosso |
| MS | Mato Grosso do Sul |
| GO | Goiás |
| DF | Distrito Federal |
| **Southeast** | |
| MG | Minas Gerais |
| ES | Espírito Santo |
| RJ | Rio de Janeiro |
| SP | São Paulo |
| **South** | |
| PR | Paraná |
| SC | Santa Catarina |
| RS | Rio Grande do Sul |

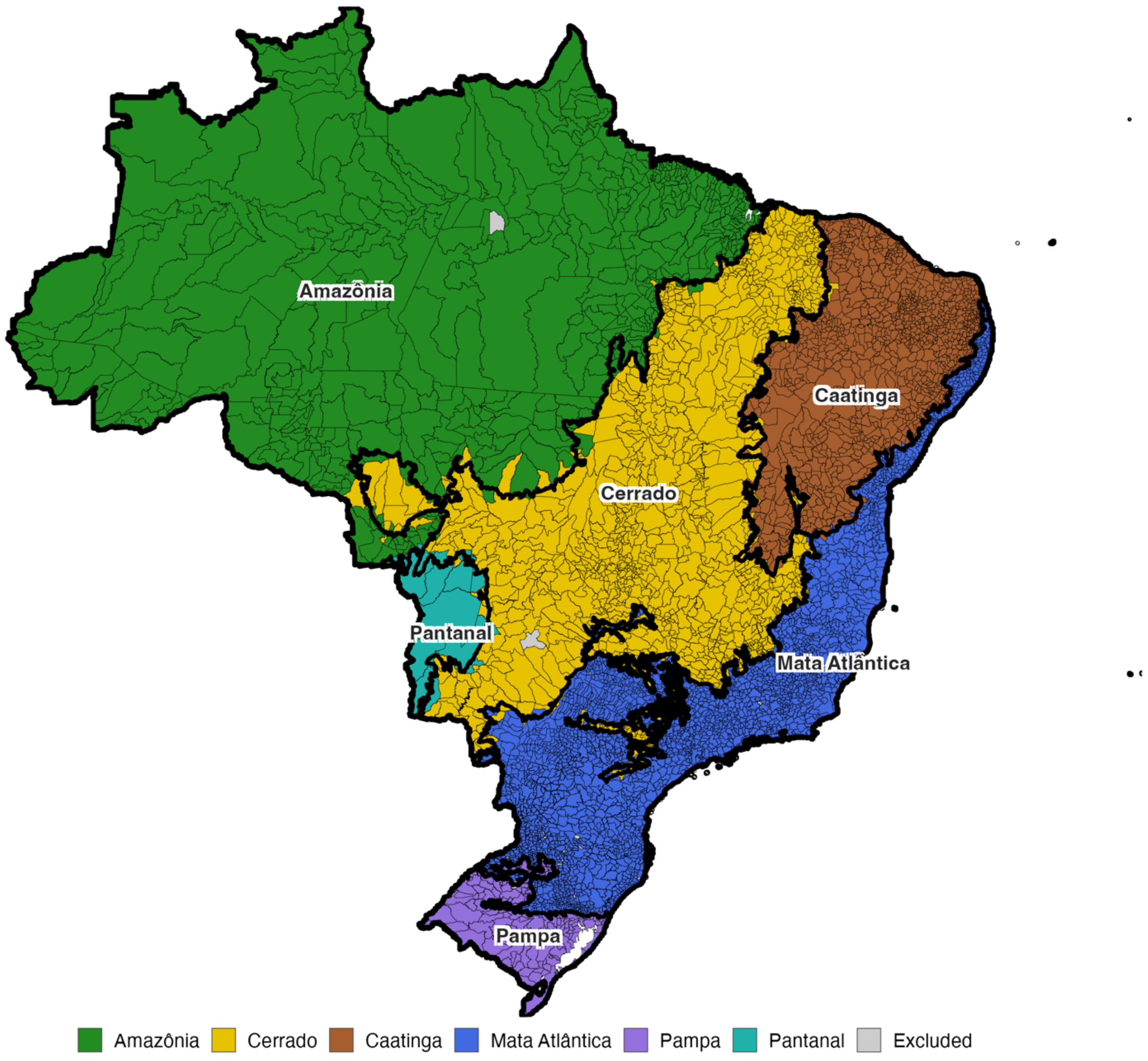


**Supplementary Fig. 1. Map of municipalities included in the analytic sample (n = 5,544), with included municipalities shaded by IBGE biome assignment.** Excluded municipalities (those with missing population denominators or boundary mismatches across census years) are shown in grey.

#### S1.4 Wildfire-related $PM_{2.5}$

##### *S1.4.1 Data source*

Daily wildfire-related $PM_{2.5}$ concentrations (0.25° spatial resolution) were obtained from the global dataset of Hu and colleagues.[4] The dataset is generated through a four-step pipeline:

1) GEOS-Chem chemical transport model simulations (v12.0.0) at 2.5° × 2.5° horizontal resolution with 47 vertical layers, driven by MERRA-2 meteorology. For each fire-emission inventory, paired simulations are run with and without biomass-burning emissions; the ratio of fire-sourced to total $PM_{2.5}$ at each grid cell is computed from the difference.

2) Machine-learning bias correction of simulated total $PM_{2.5}$ using an XGBoost gradient-boosted regression tree trained on ground-level observations from more than 9,000 monitoring stations worldwide. The correction improves the coefficient of determination of simulated $PM_{2.5}$ from 0.22 to 0.75.
3) Application of the simulated fire-to-total ratio to the bias-corrected total field to yield the final wildfire-related $PM_{2.5}$ concentration at each grid cell and day.
4) Provision of independent estimates from two fire-emission inventories: the Global Fire Emissions Database version 4.1 with small fires (GFED4.1s)[5] and the Quick Fire Emission Dataset version 2.5 (QFED2.5). [6]

**Choice of GFED4.1s as primary inventory.** We selected GFED4.1s as the primary fire-emission inventory, in line with prior epidemiological studies of wildfire $PM_{2.5}$.[7,8] GFED4.1s derives emissions from satellite-observed burned area combined with biogeochemical modelling of fuel consumption and explicitly includes small fires below the Moderate Resolution Imaging Spectroradiometer (MODIS) burned-area detection limit.[5] $QFED_{2.5}$ was used as a sensitivity analysis (Section S3.2).[6]

**Estimand and inventory scope.** GFED4.1s captures a mixture of natural wildfires, deforestation fires, and agricultural burning. Accordingly, wildfire-related $PM_{2.5}$, should be interpreted as biomass-combustion smoke from mixed sources, not smoke from a single ignition type. We follow the terminology used in previous epidemiological studies of fire-attributable $PM_{2.5}$.[7,8]

##### *S1.4.2 Threshold-based high-exposure days*

For each municipality-month, we counted the number of days on which the daily wildfire-related $PM_{2.5}$ concentration exceeded a specified threshold. The primary threshold was 25 μg/m³, corresponding to the 24-hour *Padrão Final* target under Brazilian National Environment Council resolution CONAMA 491/2018, the final stage of the national air-quality framework.[9] Sensitivity analyses used alternative thresholds of 15 μg/m³ and 35 μg/m³, corresponding to the World Health Organization 24-hour $PM_{2.5}$ guideline and the U.S. Environmental Protection Agency 24-hour $PM_{2.5}$ standard, respectively (Section S3.1). The municipalities with at least one high-exposure day at each threshold are shown in Supplementary Fig. 2.[10,11]

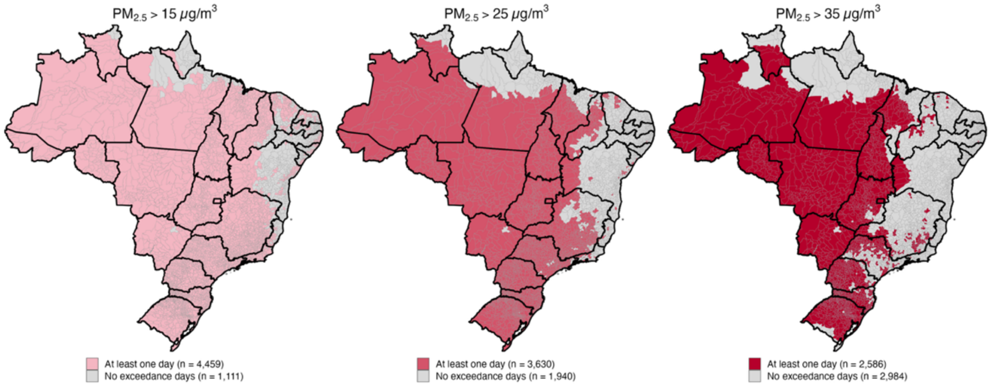


**Supplementary Fig. 2. Brazilian municipalities ever exposed to at least one day with wildfire-related $PM_{2.5}$ above a given threshold, Jan 1, 2003–Dec 1, 2023.** Each polygon is a Brazilian municipality. Within each panel, municipalities are coloured if they had at least one day during the study period with GFED4.1s wildfire-related $PM_{2.5}$ above the indicated threshold; the remaining municipalities are shown in light grey. Daily $PM_{2.5}$ exceedances were aggregated from monthly municipality-level counts. Across thresholds: 4,459 municipalities exceeded 15 μg/m³ at least once; 3,630 exceeded 25 μg/m³; 2,586 exceeded 35 μg/m³. State boundaries are drawn in black.

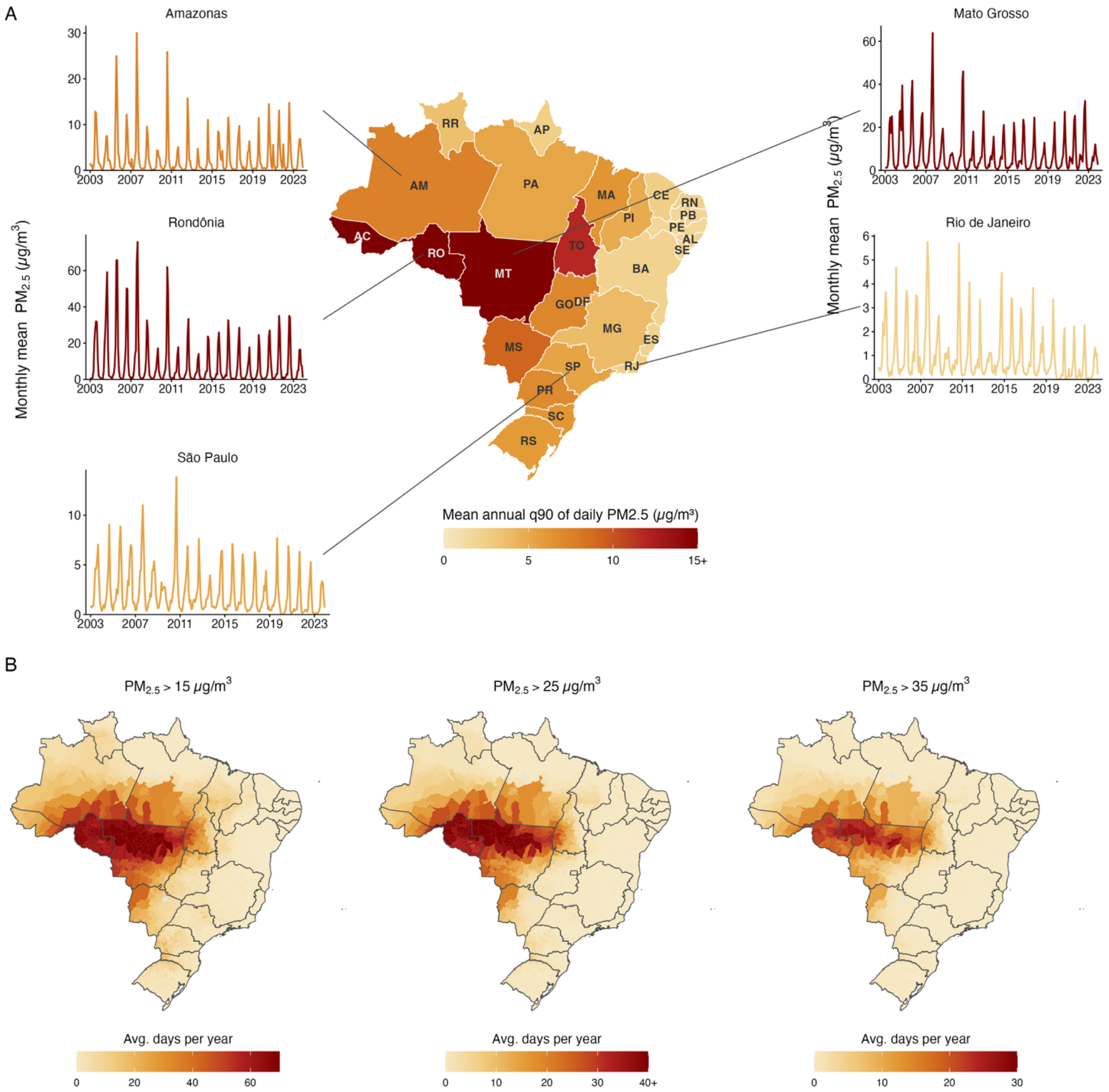


**Supplementary Fig. 3. Spatiotemporal patterns of wildfire-related $PM_{2.5}$ in Brazil, Jan 1, 2003–Dec 1, 2023.** (A) Choropleth of the mean of annual 90th-percentile daily wildfire-related $PM_{2.5}$ per state (µg/m³). For each state-year, the 90th percentile (q90) is computed over all daily municipality-level $PM_{2.5}$ observations within that state; the map shows the mean of these annual q90 values across the study period. Time-series insets show the monthly mean of GFED4.1s wildfire-related $PM_{2.5}$ for selected states (Amazonas, Rondônia, São Paulo, Mato Grosso, Rio de Janeiro), where each monthly value is the monthly mean over all municipality × day observations within that state-month. (B) Municipality-level choropleth of the average number of days per year with wildfire-related $PM_{2.5}$ concentrations exceeding 15, 25, and 35 µg/m³. Monthly high-exposure day counts per municipality were summed to annual totals and then averaged across years. Each panel uses an independently scaled colourbar; the upper limit is set to the 99th percentile of municipality values (rounded up to the nearest 10). Municipalities above this cap are rendered in the top colour of the ramp, and the top legend tick is annotated with "+" to indicate this.

##### *S1.4.3 Wildfire-related $PM_{2.5}$ event durations*

We characterized the empirical distribution of smoke-event durations, where an event is a maximal run of consecutive days exceeding 25 µg/m³ within a municipality (Supplementary Fig. 4). Events separated by a single non-exceedance day were merged into one event (gap tolerance of 1 day). One sub-threshold day between two exceedance runs typically reflects short-term meteorological variability, a transient wind shift or brief precipitation, within a single ongoing smoke episode rather than the end of one episode and the onset of another. Merging across such gaps prevents one sustained episode from being split into several short fragments, which would otherwise bias the duration distribution downward and understate the contribution of prolonged smoke. The distribution is strongly right skewed: approximately 46% of smoke events resolved within 2 days and roughly 90% within 1 week, while about 95% of events were contained within 2 weeks. The 14-day reference used in our main analysis therefore approximates the upper bound of a single sustained wildfire smoke episode. The distribution was stable across the study period (year-specific curves in Supplementary Fig. 4 largely overlap the pooled curve), indicating that the 14-day contrast carries a consistent physical meaning across 2003–2023 despite the secular increase in fire activity.

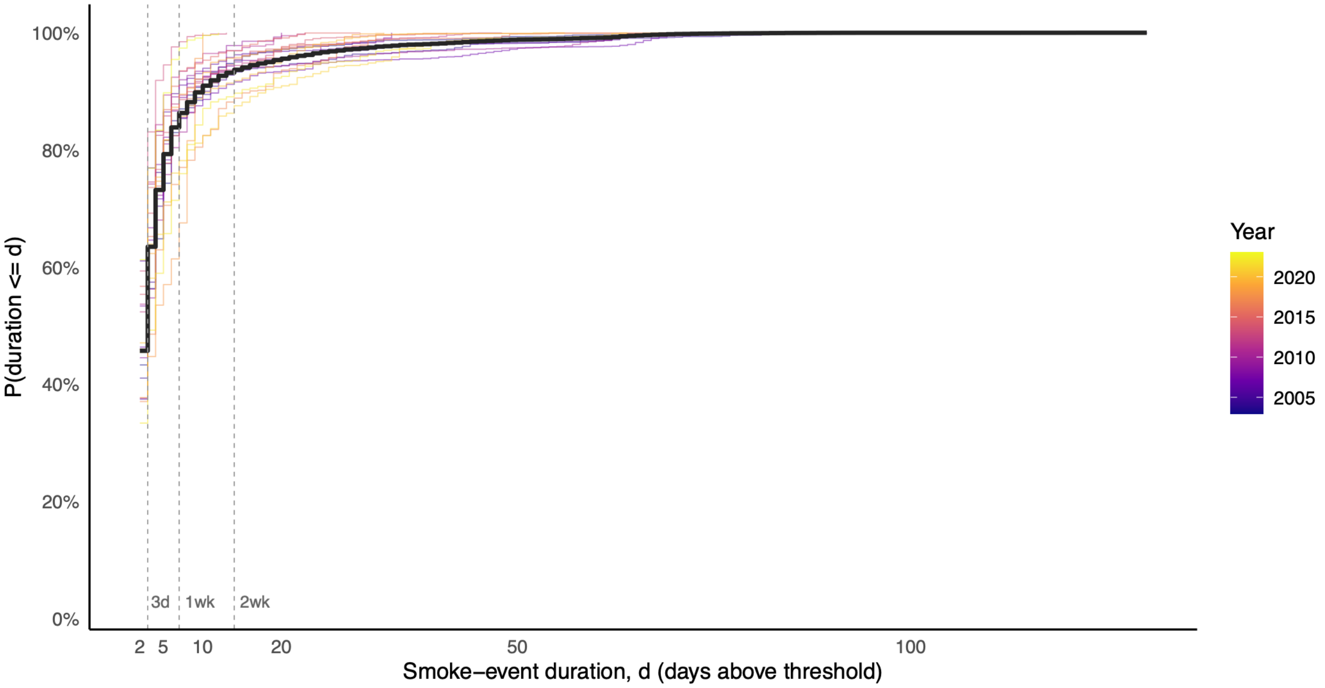


**Supplementary Fig. 4. Empirical cumulative distribution of wildfire smoke-event durations across Brazil, Jan 1, 2003 – Dec 1, 2023.** A smoke event is defined as a maximal run of consecutive days on which the daily wildfire-related $PM_{2.5}$ concentration exceeded 25 µg/m³ within a municipality, allowing a gap tolerance of 1 day: two runs separated by a single non-exceedance day were merged into one event. For each event, duration d is the number of consecutive exceedance days. The curve gives the empirical probability that an event lasts no more than d days, P(duration ≤ d), with d on a log scale. Thin coloured lines show the year-specific distributions (colour denotes year, 2003–2023); the bold black line shows the pooled distribution across all events and years. Vertical dashed reference lines mark durations of 3 days, 1 week (7 days), and 2 weeks (14 days).

**Supplementary Table 2. Wildfire-related $PM_{2.5}$ summary statistics by state, Brazil, Jan 1, 2003 – Dec 1, 2023.**
Summary statistics cover the study period Jan 1, 2003 – Dec 1, 2023. Mean $PM_{2.5}$ and 90th-percentile $PM_{2.5}$ are computed per state as the mean across years of the annual state-level average and 90th-percentile wildfire-related $PM_{2.5}$. Days per year exceeding each threshold are computed as the mean across municipalities within each state-year, then averaged across years. The last three columns give the percentage of municipality-years with at least one day above each threshold. The 'Overall' block aggregates the same municipality-level quantities across all states (National), across the arc-of-deforestation states (Rondonia, Amazonas, Mato Grosso, Acre, Tocantins, Pará), and across the remaining states (Rest of country). States in the 'By state' block are sorted by 90th-percentile $PM_{2.5}$. wildfire-related $PM_{2.5}$ is from Hu et al. 2025 (GFED4.1s).

| Region | Mean $PM_{2.5}$ (μg/m³) | 90th percentile $PM_{2.5}$ (μg/m³) | Days/yr > 15 μg/m³ | Days/yr > 25 μg/m³ | Days/yr > 35 μg/m³ | % muni-years > 15 μg/m³ | % muni-years > 25 μg/m³ | % muni-years > 35 μg/m³ |
|---|---|---|---|---|---|---|---|---|
| **Overall** | | | | | | | | |
| National | 2.4 | 6.5 | 5.8 | 2.0 | 0.9 | 43.2 | 20.4 | 8.7 |
| Arc of deforestation | 4.9 | 14.9 | 29.1 | 13.8 | 7.1 | 72.3 | 53.4 | 35.9 |
| Rest of country | 1.6 | 4.1 | 3.2 | 0.7 | 0.2 | 40.0 | 16.7 | 5.6 |
| **By state** | | | | | | | | |
| Rondônia (RO) | 8.2 | 28.3 | 63.9 | 37.3 | 20.9 | 100.0 | 95.6 | 81.7 |
| Mato Grosso (MT) | 7.4 | 21.2 | 52.2 | 24.3 | 12.4 | 95.7 | 82.1 | 56.4 |
| Acre (AC) | 4.5 | 15.2 | 32.9 | 16.8 | 8.4 | 92.2 | 80.1 | 56.7 |
| Tocantins (TO) | 4.3 | 11.9 | 21.6 | 8.3 | 3.9 | 79.3 | 45.7 | 25.5 |
| Mato Grosso do Sul (MS) | 3.8 | 9.2 | 14.4 | 4.1 | 1.6 | 89.5 | 54.0 | 19.9 |
| Amazonas (AM) | 2.8 | 7.6 | 15.6 | 6.7 | 3.1 | 76.7 | 51.8 | 30.0 |
| Goiás (GO) | 2.7 | 7.2 | 7.2 | 1.4 | 0.4 | 73.6 | 28.7 | 9.2 |
| Paraná (PR) | 3.0 | 7.0 | 8.0 | 1.9 | 0.6 | 79.0 | 40.3 | 14.1 |
| Maranhão (MA) | 2.3 | 6.3 | 4.7 | 1.0 | 0.2 | 31.4 | 8.8 | 3.9 |
| Santa Catarina (SC) | 2.5 | 6.2 | 6.7 | 1.8 | 0.6 | 67.6 | 39.2 | 17.3 |
| Distrito Federal (DF) | 2.1 | 6.0 | 3.4 | 0.3 | 0.0 | 71.4 | 14.3 | 4.8 |
| Rio Grande do Sul (RS) | 2.5 | 5.9 | 5.8 | 1.4 | 0.4 | 62.8 | 38.0 | 17.2 |
| São Paulo (SP) | 2.1 | 5.4 | 3.3 | 0.5 | 0.1 | 63.1 | 21.2 | 3.9 |
| Pará (PA) | 2.2 | 5.3 | 6.4 | 2.7 | 1.5 | 27.3 | 13.7 | 8.6 |
| Piauí (PI) | 1.8 | 4.9 | 2.3 | 0.3 | 0.1 | 22.7 | 5.5 | 1.1 |
| Minas Gerais (MG) | 1.5 | 3.9 | 1.4 | 0.2 | 0.0 | 38.6 | 9.6 | 1.5 |
| Roraima (RR) | 1.4 | 3.6 | 4.1 | 1.0 | 0.1 | 28.6 | 11.4 | 3.8 |
| Ceará (CE) | 1.0 | 2.5 | 0.2 | 0.0 | 0.0 | 4.1 | 0.9 | 0.3 |
| Amapá (AP) | 0.9 | 2.5 | 0.0 | 0.0 | 0.0 | 0.3 | 0.0 | 0.0 |
| Rio de Janeiro (RJ) | 0.9 | 2.4 | 0.6 | 0.0 | 0.0 | 22.5 | 2.3 | 0.2 |
| Rio Grande do Norte (RN) | 0.8 | 2.2 | 0.0 | 0.0 | 0.0 | 1.9 | 0.0 | 0.0 |
| Bahia (BA) | 0.9 | 2.1 | 0.3 | 0.0 | 0.0 | 4.8 | 0.6 | 0.2 |
| Espírito Santo (ES) | 0.9 | 2.1 | 0.1 | 0.0 | 0.0 | 4.9 | 0.8 | 0.2 |
| Alagoas (AL) | 0.8 | 2.0 | 0.1 | 0.0 | 0.0 | 2.9 | 0.0 | 0.0 |
| Paraíba (PB) | 0.7 | 1.9 | 0.0 | 0.0 | 0.0 | 1.2 | 0.0 | 0.0 |
| Sergipe (SE) | 0.8 | 1.8 | 0.1 | 0.0 | 0.0 | 3.1 | 0.5 | 0.0 |
| Pernambuco (PE) | 0.7 | 1.8 | 0.0 | 0.0 | 0.0 | 1.4 | 0.1 | 0.1 |

#### S1.5 Meteorological data (ERA5-Land)

Daily mean 2-metre air temperature and total precipitation were obtained from the Copernicus ERA5-Land reanalysis at native 0.1° resolution and aggregated to municipality-month means using the area-weighted zonal-statistics approach described in Section S1.14 (Supplementary Fig. 5).

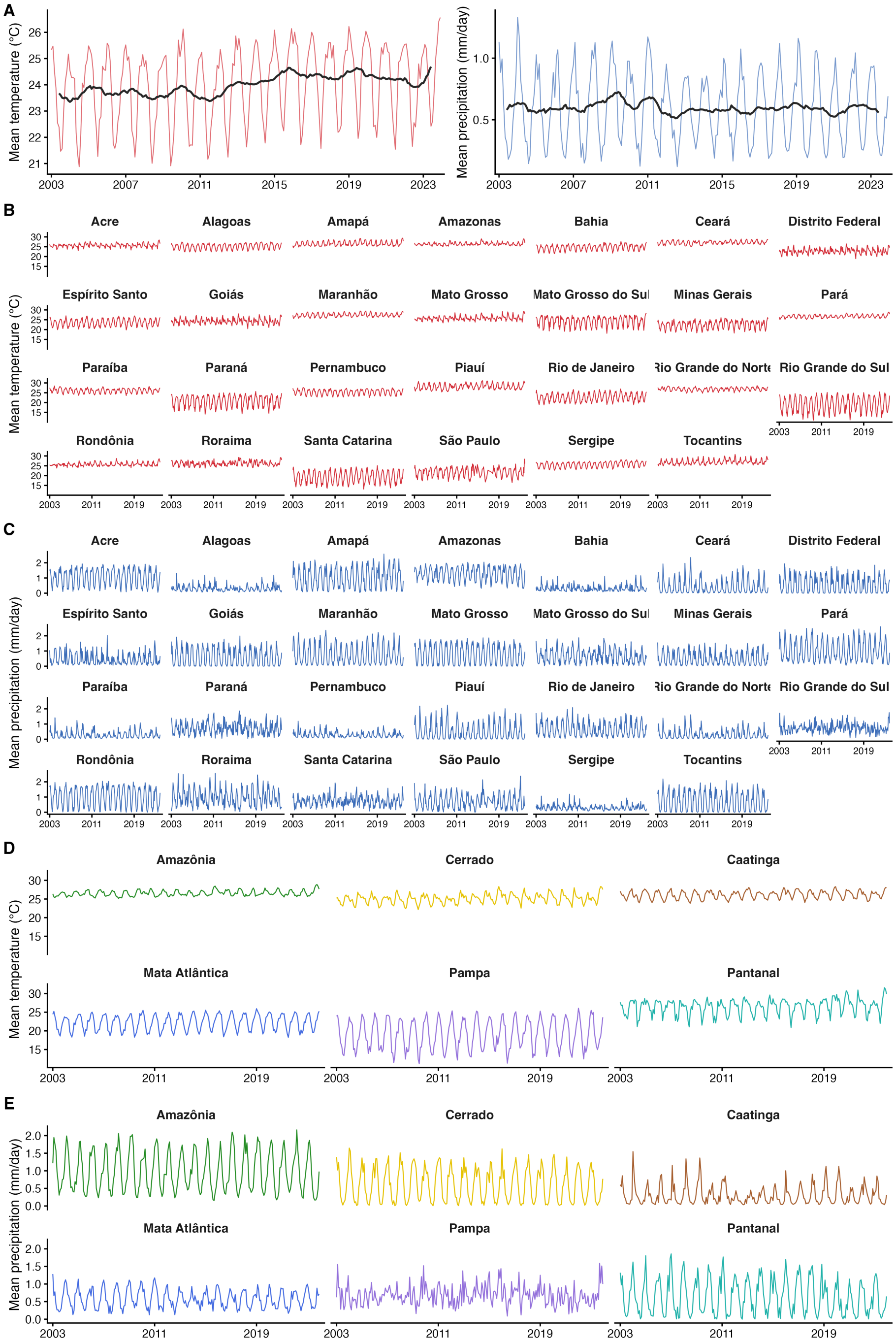


**Supplementary Fig. 5. Mean monthly air temperature and total precipitation across the analytic sample of Brazil, Jan 1, 2003 – Dec 1, 2023.** (A) Brazil overall - unweighted spatial mean across all 5,544 sampled municipalities, by calendar month (light red/blue lines). The dark line is a centred 12-month rolling mean highlighting the long-run trend. (B) and (C) Per state, showing the unweighted spatial mean across municipalities in that state. (D) and (E) Per IBGE biome (Amazonia, Cerrado, Caatinga, Mata Atlantica, Pampa, Pantanal), showing the unweighted spatial mean across municipalities in that biome; panel colour matches the biome palette used elsewhere in the manuscript.

#### S1.6 Nitrogen dioxide co-pollutant (CAMS EAC4)

Monthly mean surface $NO_2$ concentrations were obtained from the Copernicus Atmosphere Monitoring Service global reanalysis (EAC4).[12] $NO_2$ shares combustion sources with $PM_{2.5}$ but has distinct atmospheric chemistry and a markedly different spatial signature, with high concentrations in urban centres rather than at the agricultural frontier. It is therefore used to test whether the $PM_{2.5}$ signal is distinct from general combustion-related pollution (Supplementary Fig. 6).

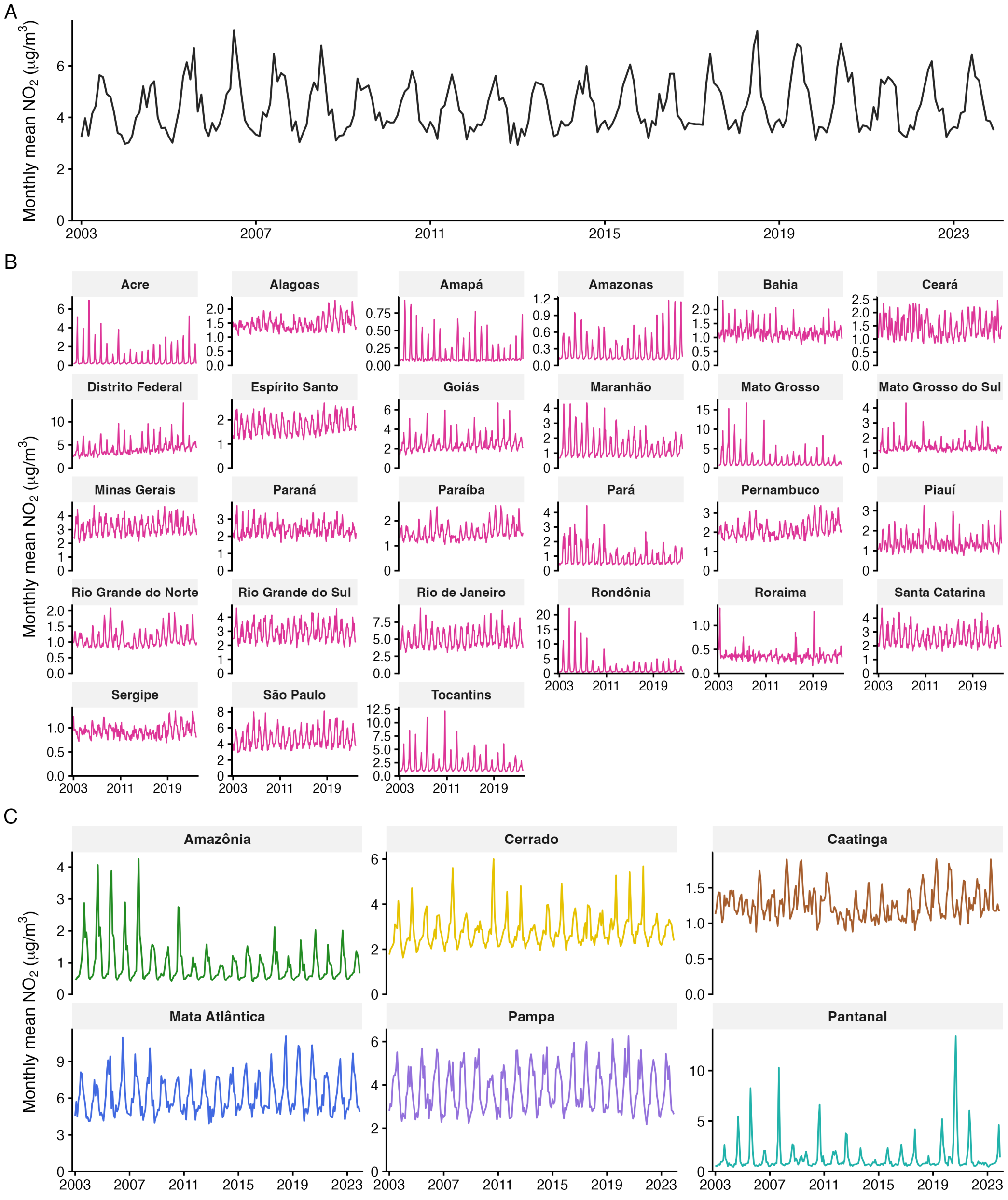


**Supplementary Fig. 6. Monthly mean ambient NO2 concentration across Brazil, Jan 1, 2003 – Dec 1, 2023.** (A) Brazil-wide monthly mean NO2 (μg/m³), computed as the population-weighted average of municipality monthly means across all municipalities in the analytic sample (n = 5,544). Weights are annual IBGE municipality population estimates. (B) Monthly mean NO2 for each of the 27 federation units (26 states + Federal District), from state-level monthly aggregates (area-weighted average across grid cells within state). Y-axes are independent across panels to reveal within-state temporal patterns. (C) Monthly mean NO2 by IBGE biome (Amazonia, Cerrado, Caatinga, Mata Atlantica, Pampa, Pantanal), computed as the population-weighted average of municipality monthly means across municipalities assigned to each biome (IBGE 2019 classification) and present in the analytic sample. Y-axes are independent across biome panels.

#### S1.7 Ozone co-pollutant (CAMS EAC4)

Monthly mean gas-phase $O_3$ concentrations were obtained from the Copernicus Atmosphere Monitoring Service global reanalysis (EAC4).[12] Unlike $PM_{2.5}$ and $NO_2$, ozone is not directly emitted but is a secondary pollutant formed photochemically downwind of fires from co-emitted precursors (nitrogen oxides and volatile organic compounds), giving it distinct atmospheric chemistry, a longer atmospheric lifetime, and a spatial signature only loosely coupled to primary smoke emissions. It is therefore used to test whether the $PM_{2.5}$ signal is distinct from co-varying photochemical (secondary) pollution (Supplementary Fig. 7).

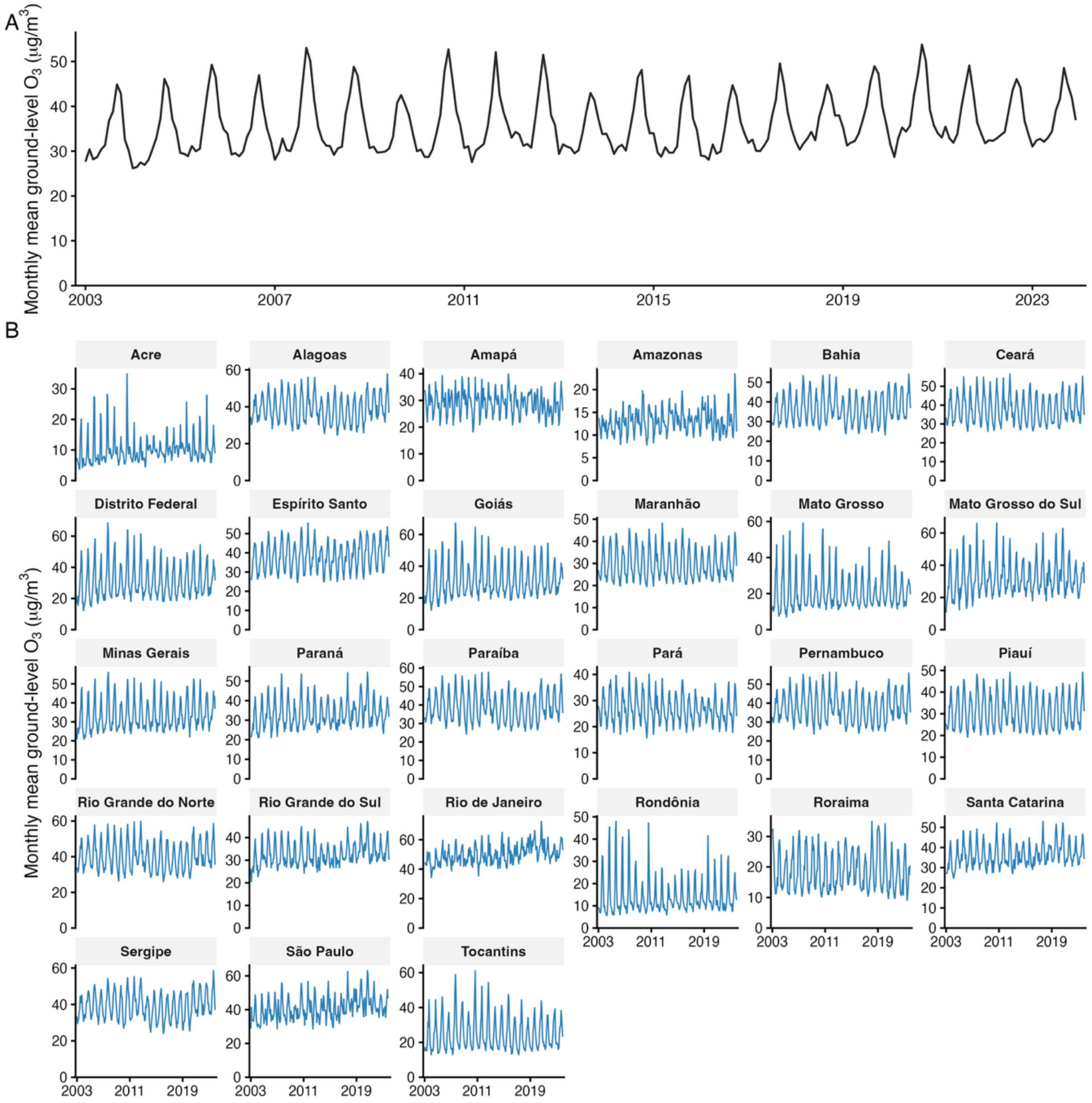


**Supplementary Fig. 7. Monthly mean gas-phase ozone (O3) concentration across the analytic sample, Jan 1, 2003 – Dec 1, 2023.** (A) Brazil-wide monthly mean O3 (μg/m³), computed as the population-weighted average of municipality monthly means across all municipalities in the analytic sample (n = 5,544). Weights are annual IBGE municipality population estimates. (B) Monthly mean O3 for each of the 27 federation units (26 states + Federal District), computed as the population-weighted average of municipality monthly means across municipalities within each unit.

#### S1.8 Land use and land cover (MapBiomas Collection 10)

Annual land-use and land-cover data at 30-metre resolution were obtained from MapBiomas Collection 10. Classification was performed using random-forest algorithms on Google Earth Engine with biome-specific training samples.[13] For each municipality and calendar year we extracted the proportion of total area classified to the MapBiomas Level 1 'Farming' class (pasture, cropland, and mixed agriculture/pasture), and the first difference between consecutive years to capture the pace of agricultural-frontier expansion. This data was used for the agricultural expansion sensitivity analysis (Section S3.6; Supplementary Fig. 8).

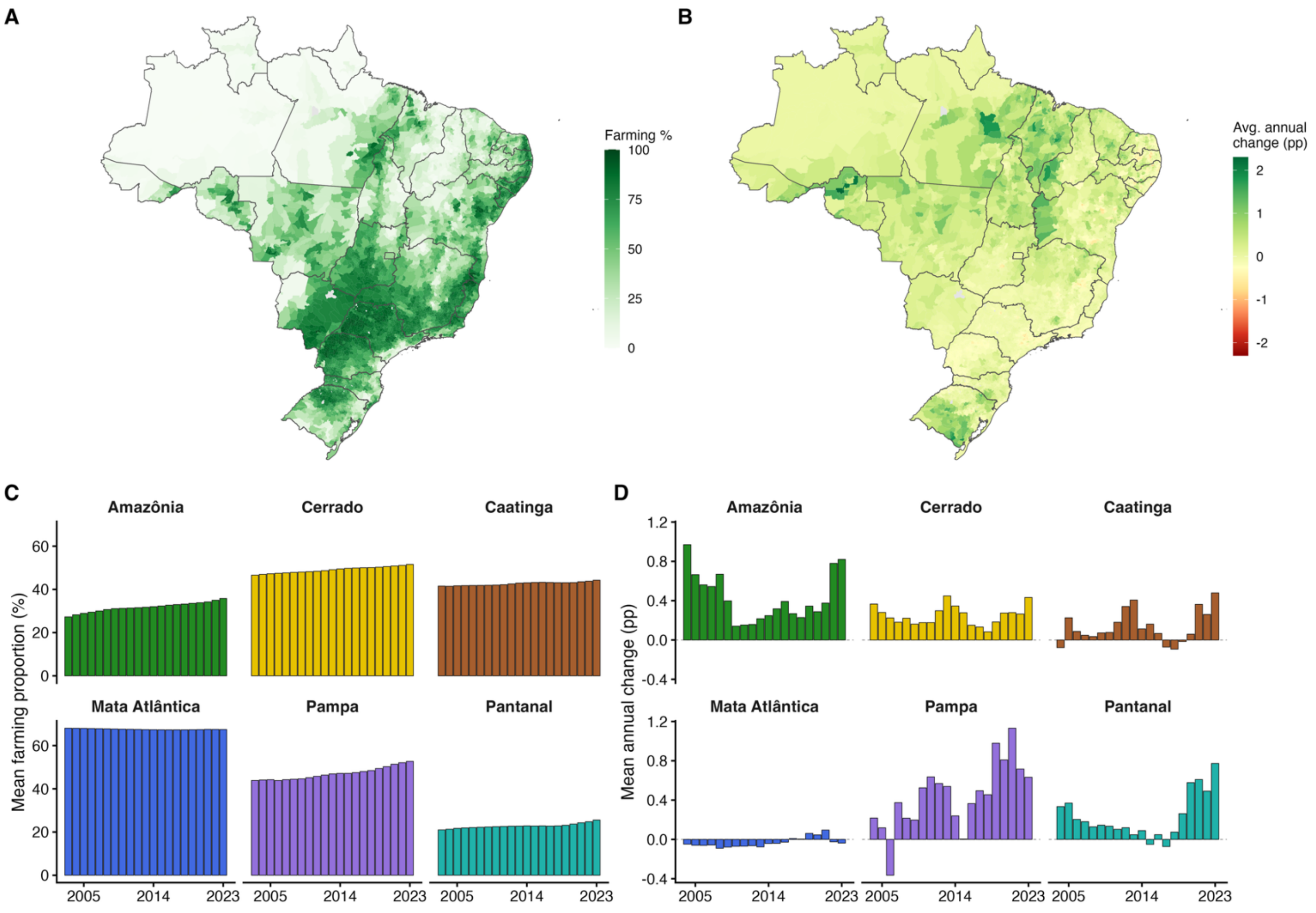


**Supplementary Fig. 8. Farming land-use proportion and year-to-year change across the analytic sample of Brazilian municipalities, 2003–2023.** (A) Choropleth of the average farming proportion (percent of municipal area classified as MapBiomas Level 1 'Farming') per municipality, averaged across all years in the study period. Greys indicate municipalities outside the analytic sample or with no farming observations. (B) Choropleth of the average annual change (delta, percentage points) in farming proportion per municipality. The diverging palette is symmetric around zero; reds indicate average annual losses; greens indicate average annual gains. (C) Mean farming proportion over time, per IBGE biome (Amazônia, Cerrado, Caatinga, Mata Atlântica, Pampa, Pantanal). Each series is the unweighted spatial mean across municipalities in that biome; panel colour matches the biome palette used elsewhere in the manuscript. (D) Mean annual change in farming proportion (percentage points) over time, one small multiple per IBGE biome. The dashed grey line marks zero (no year-to-year change).

#### S1.9 Biome fire regimes and exposure characteristics

Brazil's six biomes differ in vegetation, in their ecological relationship with fire, and in the land types that burn.[14] The grass-dominated savannas and grasslands of the Cerrado and Pampa are fire-adapted systems in which burning is a long-standing ecological process, whereas the tropical forests of Amazônia and Mata Atlântica are fire-sensitive. In Amazônia, fire is predominantly human-ignited and linked to deforestation and

pasture management. The Pantanal, a seasonally flooded mosaic of savanna, forest and grassland, is also fire-adapted, whereas natural fire is rare in the Caatinga, where dry shrubland and forest lack continuous flammable fuel.
We summarized these differences using four descriptive measures (Supplementary Fig. 9).

Savanna, grassland and shrubland [including pasture] accounted for most burned area in every biome, with forest contributing 32% in Amazônia and cropland 48% in Mata Atlântica (Supplementary Fig. 9A). High-exposure days were concentrated in August–October in all biomes (Supplementary Fig. 9B). In Mata Atlântica and Pampa, most exposed municipality-months had no detected fire in the municipality or its neighbours, indicating smoke transported from fires elsewhere (Supplementary Fig. 9C; Section S3.15.1).

Within-municipality variation in annual high-exposure days, which identifies the association under the fixed-effects design (Section S2.1), was largest in Pantanal and Amazônia and smallest in Caatinga, which contributed only 144 exposed municipality-months (Supplementary Fig. 9C,D). Caatinga could therefore not support a biome-specific lag–response and was grouped with Cerrado, with which it shares a burned-area profile dominated by savanna, grassland and shrubland and a late dry-season exposure peak. Mata Atlântica and Pampa were grouped because both had little local burning (less than 1% of biome area per year), low within-municipality exposure variation, and exposure arising predominantly from transported smoke (Supplementary Fig. 9A,C,D).

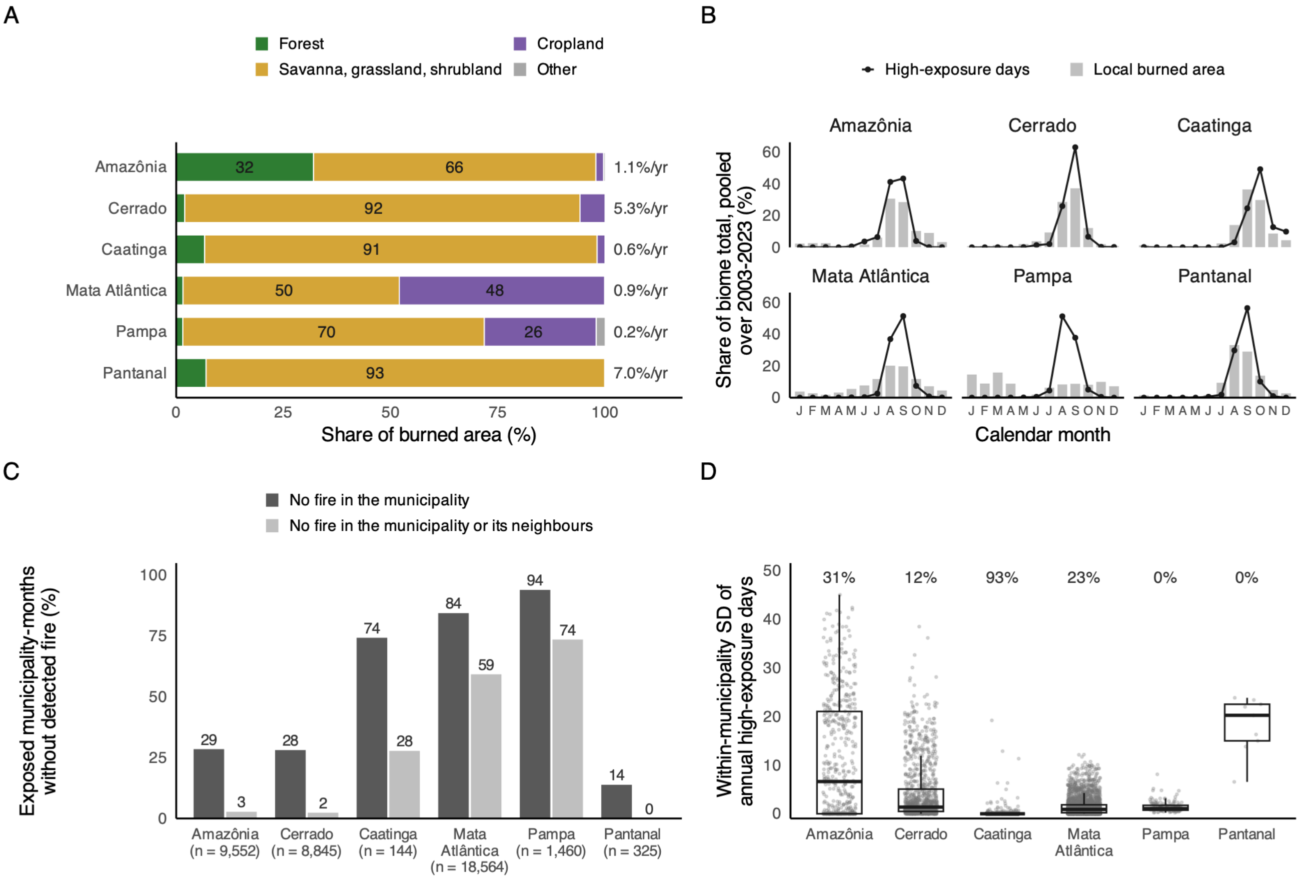


**Supplementary Fig. 9. Fire regime and wildfire-related $PM_{2.5}$ exposure by biome, Brazil, Jan 1, 2003 – Dec 31, 2023.** (A) Burned area by MODIS land-cover class at ignition, as a percentage of each biome's total burned area (Global Fire Atlas); labels at right give the mean annual burned area as a percentage of biome area. (B) Seasonality of local burning and of exposure: bars show the share of each biome's burned area by calendar month, and lines the share of its high-exposure days (wildfire-related $PM_{2.5}$ above 25 µg/m³, GFED4.1s) by calendar month; both are pooled over all municipalities in the biome and all years of the study period (each month's total divided by the biome's total over the period). (C) Percentage of exposed municipality-months (at least one high-exposure day) in which no fire was detected in the municipality (dark) or in the municipality or any neighbouring municipality (light); n is the number of exposed municipality-months. (D) Within-municipality standard deviation of annual high-exposure days, the year-to-year variation that identifies the exposure effect after municipality fixed effects; each point is a municipality, boxes show the median and IQR, and numbers above give the percentage of municipalities with no high-exposure day over the study period. Fires smaller than one MODIS pixel (about 0.21 $km^2$) are not detected, so local agricultural burning is under-counted in (A)-(C).

#### S1.10 Drought index

Drought conditions in Brazil can be characterized using the Standardized Precipitation–Evapotranspiration Index at the 12-month accumulation scale (SPEI-12) from the CSIC SPEIbase v2.11 (https://spei.csic.es/spei_database_2_11), aggregated to municipalities by coverage-weighted zonal means (Supplementary Fig. 10). SPEI can be used for determining the onset, duration and magnitude of drought conditions with respect to normal conditions in a variety of natural and managed systems such as crops, ecosystems, rivers, water resources, etc.

We included SPEI-12 in a sensitivity analysis because drought is a plausible shared driver of wildfire-related $PM_{2.5}$ exposure and TB notification rates: dry conditions promote fire spread and smoke production, while prolonged drought may independently influence TB risk through food insecurity, undernutrition, population displacement, and disruption to health-care access. Thus, adjustment for SPEI-12 allowed us to assess whether the wildfire-related $PM_{2.5}$ association was robust to confounding by sustained drought conditions not fully captured by monthly temperature and precipitation.

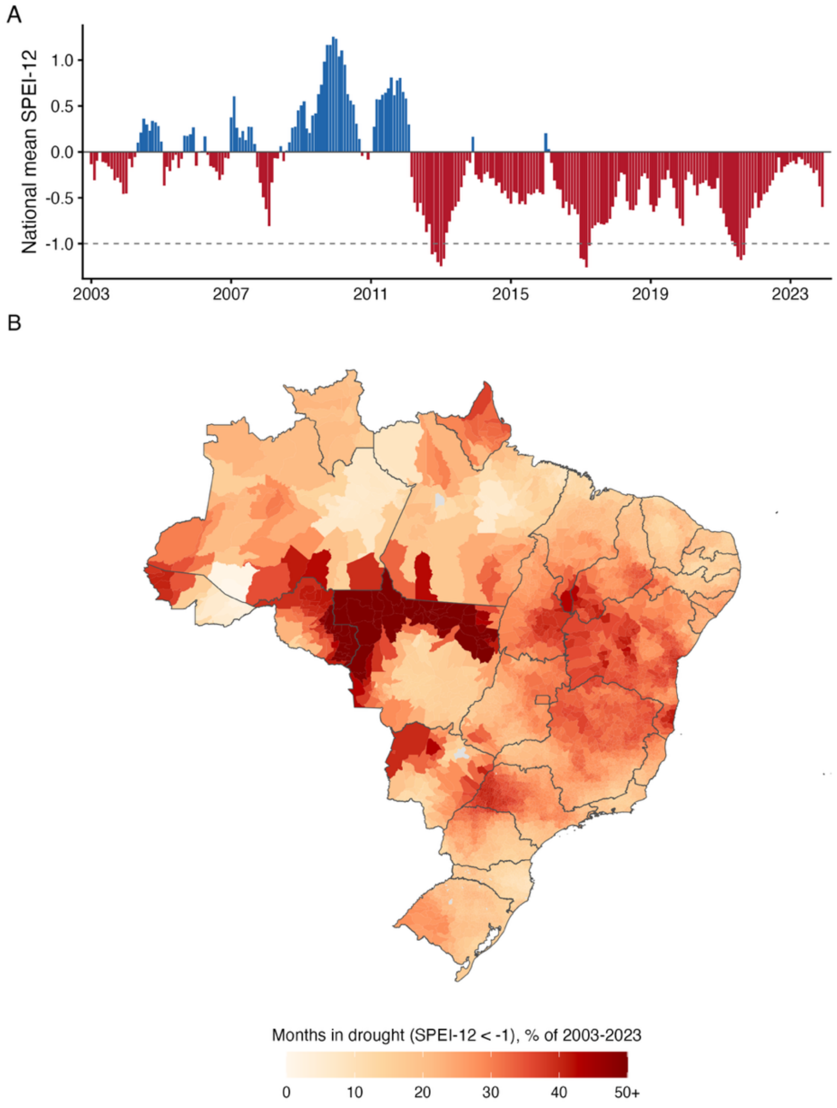


**Supplementary Fig. 10. SPEI-12 (12-month) drought index across the analytic sample of Brazilian municipalities, Jan 1, 2003 – Dec 1, 2023.** The Standardized Precipitation-Evapotranspiration Index (SPEI; CSIC SPEIbase v2.11) is a z-scored, multi-scale drought index defined relative to the 1901-2020 climatology: negative values indicate drier than normal conditions (hydrological (approximately annual) drought), positive values wetter. The 12-month accumulation integrates the water balance over that window. (A) Unweighted spatial mean SPEI-12 across the 5,544 analytic-sample municipalities, by calendar month. Bars below zero (drier than climatology) are red; bars above zero (wetter) are blue. The dashed line at SPEI-12 = -1 marks the moderate-drought threshold. (B) Municipality-level choropleth of the percentage of study-period months in moderate-or-worse drought (SPEI-12 < -1). Municipalities above the colour-scale cap (50%) are drawn in the top colour and the top legend tick is annotated with "+". Municipalities outside the analytic sample (or with only ocean grid-cell coverage) are shown in grey; state borders are overlaid in grey.

#### S1.11 Leprosy

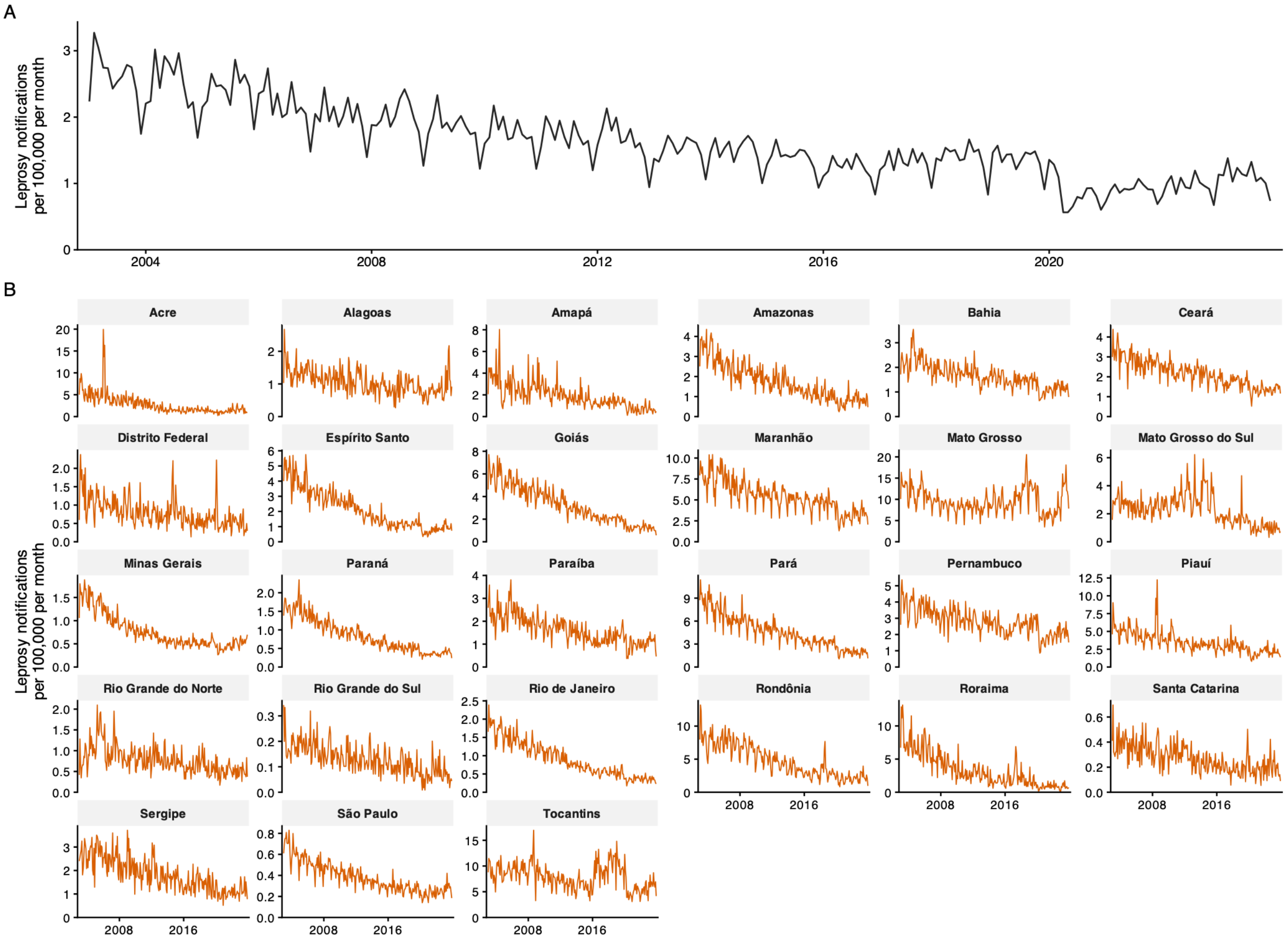


**Supplementary Fig. 11. Monthly leprosy (hanseniase) notification rate in Brazil, Jan 1, 2003 – Dec 1, 2023.** (A) Brazil-wide monthly leprosy notification rate per 100,000 population, computed as the national monthly notification count divided by that year's national population, x 100,000. (B) Monthly leprosy notification rate per 100,000 for each of the 27 federation units (26 states + Federal District), using state population as the denominator.

#### S1.12 Appendicitis

Appendicitis data are used for a negative control outcome analysis described in Section S3.14.2. Across January 1, 2003 to December 1, 2023, the Hospital Information System (SIH-RD) recorded 1,879,350 hospital admissions for acute appendicitis (primary diagnosis ICD-10 K35) across Brazilian municipalities. The national monthly count rose from roughly 4,400 admissions per month at the start of the period to about 10,200 by the end, corresponding to a crude annual rate of approximately 31.8 per 100,000 in 2003 and 54.5 per 100,000 in 2019; expressed as the monthly rate, appendicitis admissions ranged from about 2.5 to 4.9 per 100,000 per month (Supplementary Fig. 12). Part of the early-period increase reflects the expanding municipal coverage of SIH rather than a true rise in incidence, as the number of municipalities recording at least one admission grew through the 2000s. A marked, transient decline in 2020 coincides with COVID-19 disruption to hospital care. Absolute admission counts are dominated by the most populous states (Sao Paulo, Minas Gerais) and therefore track state population, whereas the population-standardized admission rate is considerably more uniform across the 27 federation units, consistent with appendicitis being a common acute surgical condition with broadly similar occurrence nationwide. Admissions are restricted to the public (SUS) system, are attributed to the patient's municipality of residence, and are dated by month of admission.

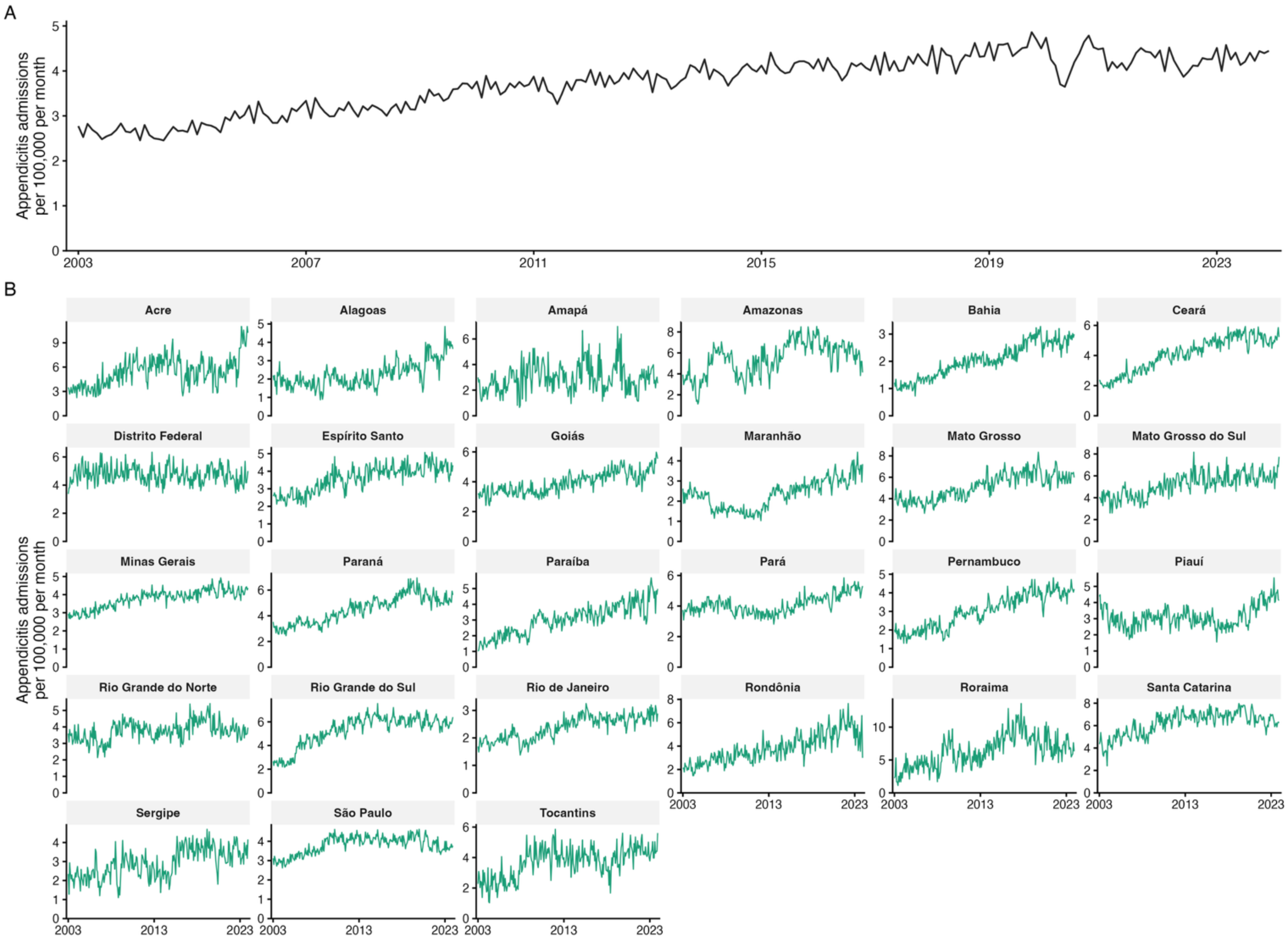


**Supplementary Fig. 12. Monthly appendicitis hospital admission rate in Brazil, Jan 1, 2003 – Dec 1, 2023.** (A) Brazil-wide monthly appendicitis admission rate per 100,000 population, computed as the national monthly admission count divided by that year's national population, x 100,000. (B) Monthly appendicitis admission rate per 100,000 for each of the 27 federation units (26 states + Federal District), using state population as the denominator. Y-axes are independent across panels (note scales); per-capita admission rates are far more uniform across states than the absolute counts in the companion figure, since the population denominator removes the dominance of the most populous states. Admissions are Sistema de Informacoes Hospitalares do SUS (SIH-RD) hospitalizations for a primary diagnosis of acute appendicitis (ICD-10 K35) provided by Brazilian Ministry of Health, counted at the patient's municipality of residence and dated by month of admission. SIH-RD covers SUS (public-system) hospitalizations only, so the private sector is not represented. Municipality-to-state assignment uses the first two digits of the IBGE municipality code. One state-month is absent at source (Amapa, October 2007).

#### S1.13 Malnutrition

Malnutrition data are used for the alternative-pathway analysis described in Section S3.15.2. Admissions were obtained from the Hospital Information System (SIH-RD)[15] using the microdatasus R package[16] and restricted to those with a principal diagnosis of protein-energy malnutrition (ICD-10 E40–E46).[17] These codes are consistent with previous national analysis of malnutrition admissions in Brazil.[18] Between January 1, 2005 and December 1, 2023, SIH-RD recorded 803,522 such admissions, 85% of them in patients aged 15 years or older. The crude admission rate fell from 31.8 per 100,000 in 2005 to 12.3 per 100,000 in 2023, with an additional drop in 2020 that coincides with COVID-19 disruption to hospital care (Supplementary Fig. 13). Several states show abrupt shifts in level between consecutive years (for example, Bahia around 2016 and Paraíba around 2013). These more likely reflect changes in hospital coding or service provision than changes in incidence and are absorbed by the state × year fixed effects in our panel regression. These malnutrition admissions are those funded by SUS, are attributed to the patient's municipality of residence, and are dated by the month of admission. Municipality-months without an admission were treated as zero counts; 76% of municipality-months had no admissions.

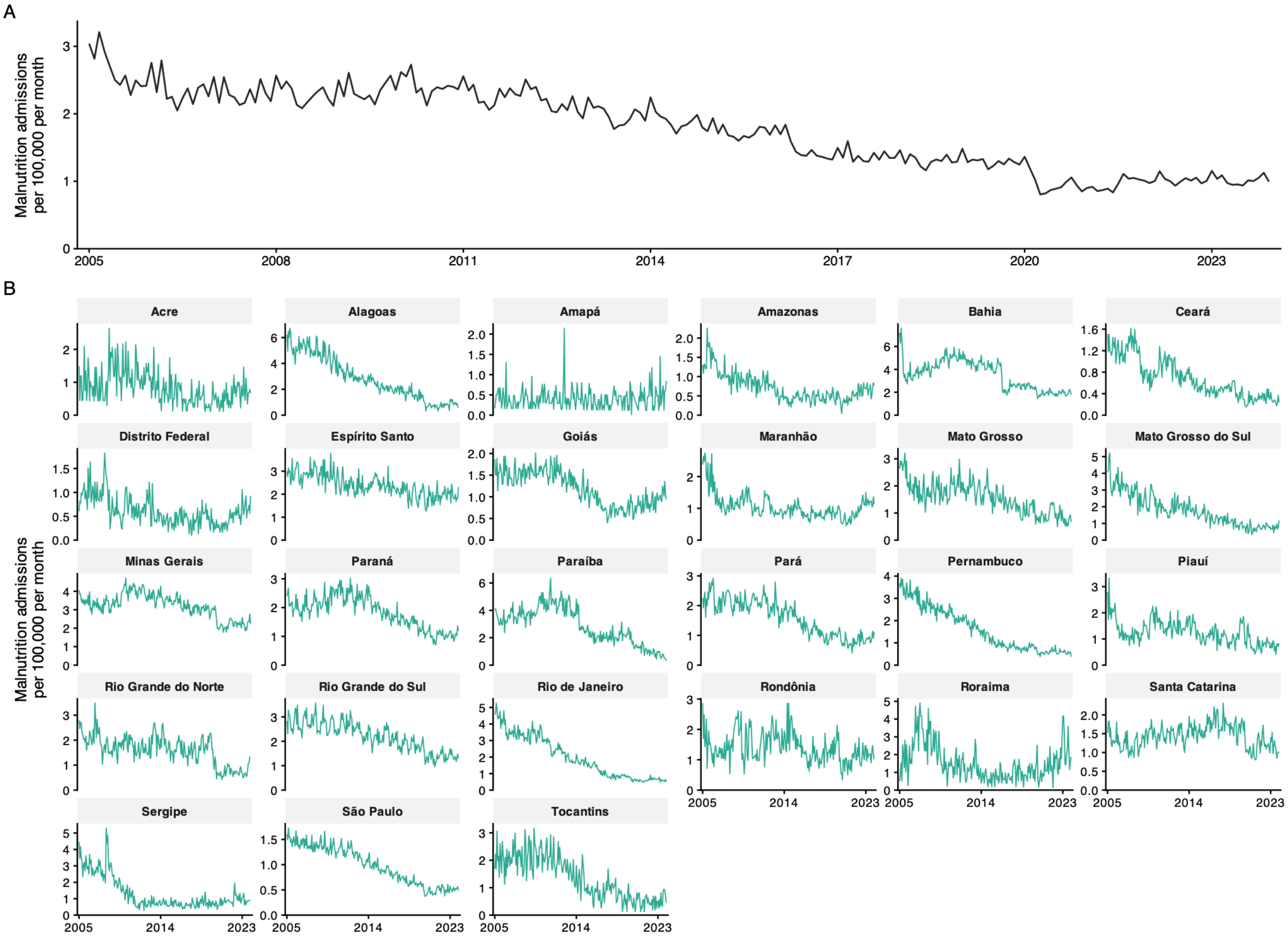


**Supplementary Fig. 13. Monthly malnutrition hospital admission rate in Brazil, Jan 1, 2005 – Dec 1, 2023.** (A) Brazil-wide monthly admission rate per 100,000 population, computed as the national monthly admission count divided by that year's national population, x 100,000. (B) Monthly admission rate per 100,000 for each of the 27 federation units (26 states and the Federal District), using state population as the denominator; y-axes are independent across panels. Admissions are SIH-RD hospitalizations with a principal diagnosis of protein-energy malnutrition (ICD-10 E40-E46), counted at the patient's municipality of residence and dated by month of admission. SIH-RD covers hospitalizations funded by the Unified Health System (SUS) only. Rates are monthly; denominators are annual IBGE municipal population estimates, summed to the state and national levels.

#### S1.14 Spatial assignment of gridded exposures

Daily gridded wildfire $PM_{2.5}$ values at 0·25° resolution were assigned to Brazilian municipalities using area-weighted zonal statistics implemented via the exactextractr R package.[19] For each municipality polygon, the contribution of each overlapping grid cell was weighted by the fraction of the municipality's area falling within that cell. This avoids the bias that arises from simple centroid-based assignment when municipalities span multiple grid cells or when grid cells partially overlap municipal boundaries. The same procedure was applied to ERA5-Land meteorology and CAMS NO2 fields.

#### S1.15 Lag variable construction

Monthly exceedance-day counts were lagged from 1 to 24 months prior to the notification month. The 24-lag window was selected to represent a plausible biological interval from $PM_{2.5}$ exposure to TB notification, including both proximal effects on innate immunity and accelerated progression from latent infection to active disease over many months.

#### S1.16 GeneXpert diagnostic coverage

The GeneXpert MTB/RIF molecular diagnostic platform was introduced in Brazil from approximately 2014, with phased state-by-state rollout.[20] GeneXpert has substantially higher sensitivity than smear microscopy, and its introduction could increase TB notification rates independently of true incidence. To account for this time-varying source of differential case detection, we computed the yearly proportion of TB notifications in each municipality that were diagnosed using GeneXpert and included this measure as a proxy for local diagnostic coverage (Supplementary Fig. 14).

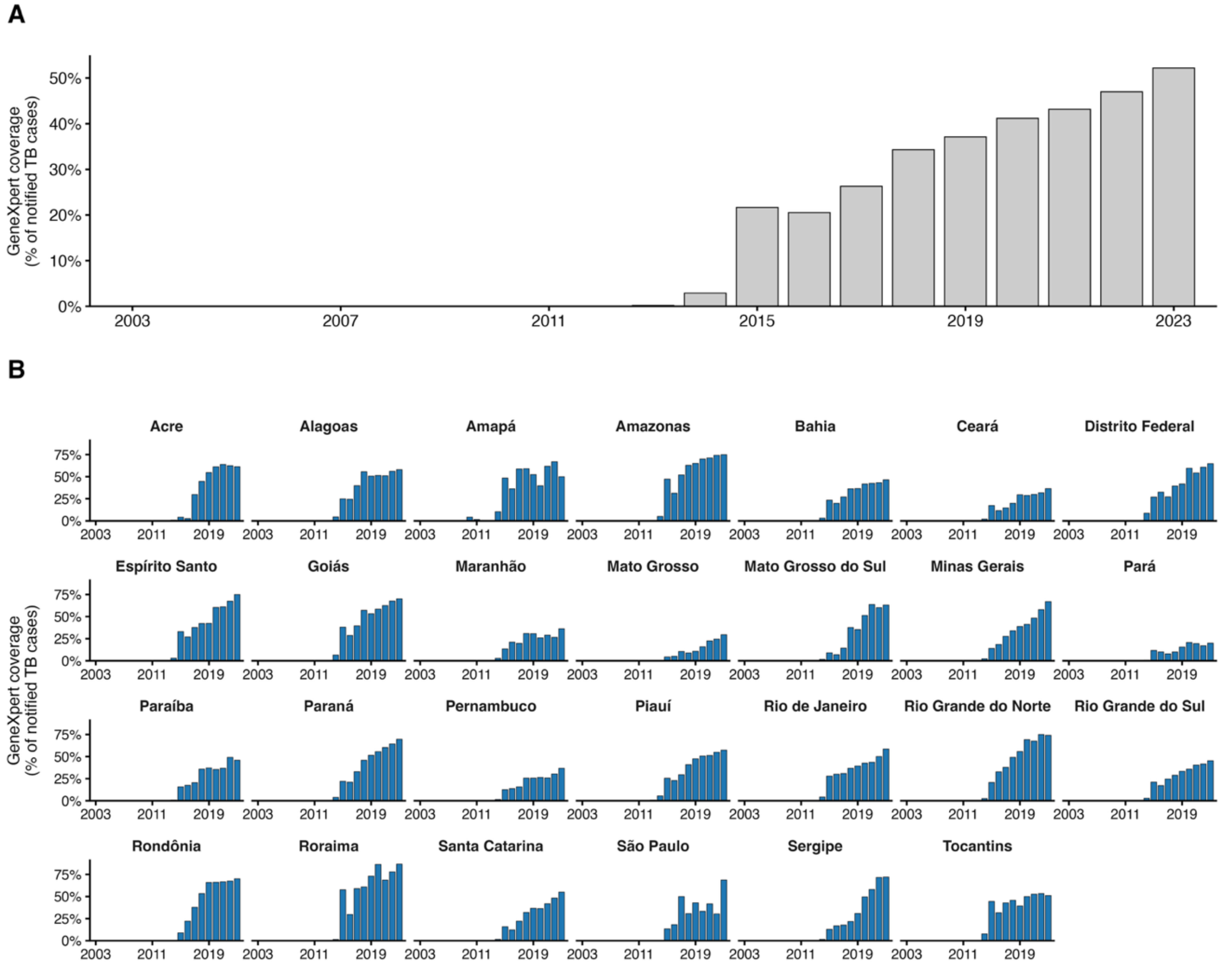


**Supplementary Fig. 14. Annual GeneXpert (rapid molecular test) coverage among notified TB cases across the analytic sample of Brazilian municipalities, Jan 1, 2003–Dec 1, 2023.** Coverage is approximated by the share of notified TB cases (SINAN, TEST_MOLEC variable) for which a rapid molecular test result was recorded, i.e., TEST_MOLEC coded as "Detectable — sensitive to rifampicin", "Detectable — resistant to rifampicin", "Not detectable", or "Inconclusive". Cases with TEST_MOLEC coded "Not performed" or missing are treated as not covered. Aggregation is case-weighted (sum of cases with a recorded result divided by sum of notified cases within each year-scope). (A) Brazil overall: national case-weighted coverage across all 5,544 sampled municipalities, by calendar year. (B) Case-weighted coverage across municipalities per state.

#### S1.17 Pandemic-phase definition

The COVID-19 pandemic disrupted TB surveillance in Brazil through facility closures, redirected diagnostic capacity, and changes in care-seeking behaviour. To prevent these disruptions from contaminating the lag-response estimates, we defined a categorical pandemic-phase variable taking five values:

**Supplementary Table 3. Pandemic-phase categories and date cutoffs.**

| Phase | Cutoff |
|---|---|
| pre_pandemic | < 2020-03-01 |
| wave1_ancestral | 2020-03-01 → 2020-10-31 |
| wave2_gamma | 2020-11-01 → 2021-10-31 |
| wave3_omicron | 2021-11-01 → 2022-04-30 |
| post_acute | ≥ 2022-05-01 |

### S2. Statistical methods

#### S2.1 Panel fixed-effects design and confounding control overview

Our identification strategy relies on within-municipality temporal variation in wildfire-related $PM_{2.5}$ over time. By comparing each municipality to itself across months, the ecological panel design reduces confounding by stable municipal characteristics, while the fixed-effects structure and time-varying covariates address major sources of temporal confounding. Conditional on this control structure and supported by the consistency of results across robustness and negative-control analyses described below, we interpret the estimated associations as population-level causal effects, while recognizing that the unit of observation is the municipality-month and that individual-level causal effects cannot be established.[21]

Three potential sources of confounding are addressed explicitly. First, time-invariant municipal characteristics, including geography, baseline health infrastructure, urbanization, and economic structure, are absorbed by municipality fixed effects. Second, state-level shocks and secular trends, including changes in TB control programmes, policy changes, and COVID-19-related disruptions to TB surveillance, are absorbed by state × year and state × pandemic-phase fixed effects. Third, seasonal patterns in both TB diagnosis and fire activity are absorbed non-parametrically by state × month fixed effects. After conditioning on these components, identification comes from deviations in a municipality's exposure in a given month relative to its own long-run average, its state's exposure pattern in that year, its state's typical seasonal pattern, and its state's pandemic-phase pattern.

The distributed lag structure, spanning 1–24 months before notification, was used to evaluate whether associations followed a biologically plausible temporal pattern for TB progression and to reduce the likelihood that short-term diagnostic or care-seeking changes explained the results.

#### S2.2 Fixed-effects specification

The model includes four blocks of fixed effects:

5) Municipality ($\alpha_i$): absorbs time-invariant differences across municipalities, including geography, baseline health infrastructure, population density, urbanization, and mining-dependent economies.
6) State × year ($\gamma_{state(i),\ year(t)}$): absorbs annual state-level shocks, including TB programme intensification, state-level policy changes, and secular trends in TB notification.
7) State × month ($\delta_{state(i),\ month(t)}$): absorbs state-specific seasonal patterns in TB notification, healthcare utilization, and diagnostic activity, which vary across Brazil's diverse climate zones.
8) State × pandemic-phase ($\zeta_{state(i),\ phase(t)}$): absorbs state-specific surveillance disruption during and after the COVID-19 pandemic. Pandemic-phase categories are defined in Section S1.17.

After conditioning on all four blocks, identification comes from the remaining within-municipality monthly deviations after removing municipality averages, annual state trends, seasonal state patterns, and pandemic-phase disruptions (Supplementary Fig. 15).

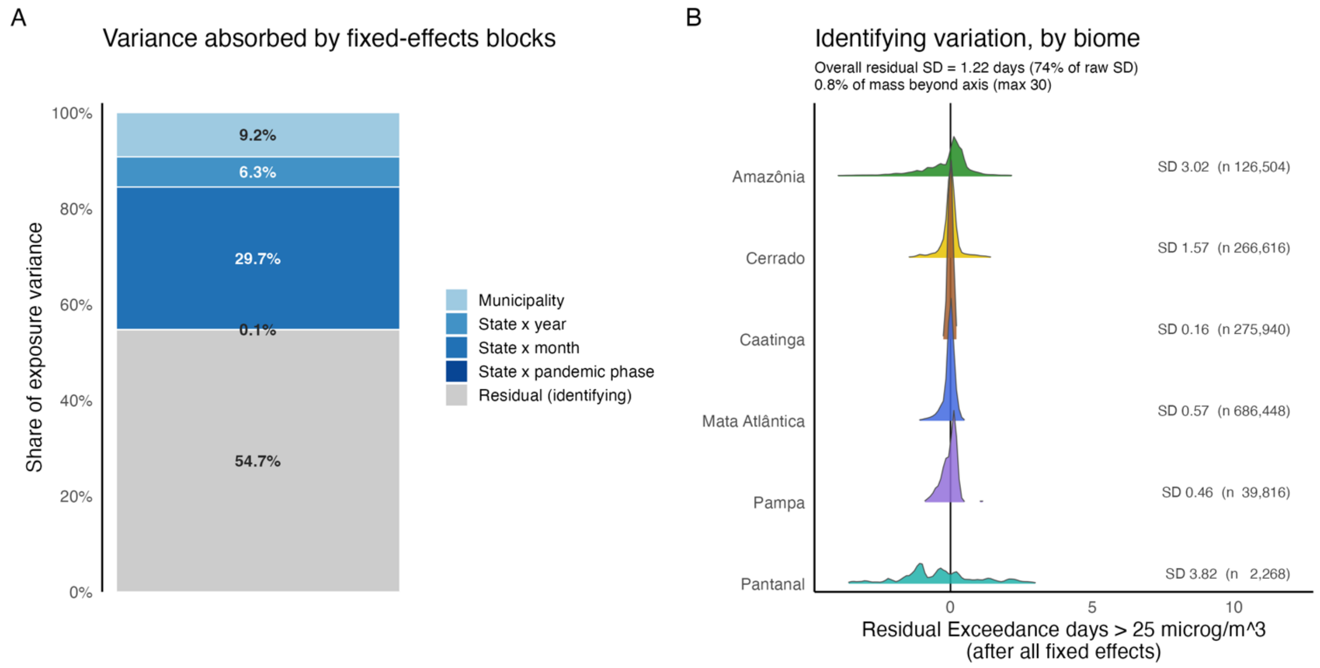


**Supplementary Fig. 15. Decomposition of the variance in monthly wildfire-related $PM_{2.5}$ exposure absorbed by the fixed-effects structure.** Panel (A) shows the cumulative $R^2$ contribution of each fixed-effects block (municipality; state × year; state × month; state × pandemic-phase). Panel (B) shows the distribution of residual within-municipality variation in exceedance days that identifies the effect, by biome.

#### S2.3 Covariates

The model includes a set of covariates whose functional form and lag structure follow from the role each plays in the identification strategy:

- **Temperature and precipitation (Section S1.5)** enter as natural-spline cross-basis terms with 3 degrees of freedom on the exposure dimension and a 24-month lag dimension, matching the primary exposure, allowing non-linear and delayed effects on TB notification while remaining parsimonious.
- **GeneXpert diagnostic coverage (Section S1.16)** enters as a contemporaneous (unlagged) linear covariate defined as the proportion in each current municipality-year, assuming that the platform affects the probability of detecting an existing case at the time of diagnosis.
- **Pandemic-phase (Section S1.17) enters only** through interaction with state identity in the state × pandemic-phase fixed-effects block (Section S2.2), absorbing differential surveillance disruption across states without constraining its functional form.
- **NO2 (Section S1.6) and the MapBiomas land-use measures (Section S1.8) are not** part of the primary specification; they enter sensitivity analyses (Sections S3.3 and S3.6 respectively) used to test the robustness of the wildfire $PM_{2.5}$ lag-response to combustion co-pollutants and to agricultural-frontier dynamics.

#### S2.4 Poisson regression

The primary model is a Poisson generalized additive panel regression[22] with a penalized distributed lag in wildfire-related $PM_{2.5}$ exposure. The primary model takes the form:

$$\log E(Y_{it}) = \alpha_i + \gamma_{state(i),year(t)} + \delta_{state(i),month(t)} + \zeta_{state(i),phase(t)} + \sum_{\ell=1}^{24} \beta_\ell \cdot \text{Exposure}_{i,t-\ell} + \sum_{\ell=1}^{24} g_T(\ell) \cdot \text{Temp}_{i,t-\ell} + \sum_{\ell=1}^{24} g_P(\ell) \cdot \text{Precip}_{i,t-\ell} + \theta \cdot \text{GeneXpert}_{i,year(t)} + \log(\text{Pop}_{i,year(t)})$$

where

- $Y_{it}$ is the count of TB notifications in municipality i and month t;
- α, γ, δ and ζ are the fixed effects for municipality, state-by-year, state-by-month, and state-by-pandemic-phase (Section S2.2), where state(i) is the state of municipality i and year(t), month(t) and phase(t) are the calendar year, calendar month and pandemic phase of month t;
- $\text{Exposure}_{i,t-\ell}$ is the number of high-exposure days (days with wildfire-related $PM_{2.5}$ above 25 μg/m³) in municipality i in month t−ℓ, for lags ℓ = 1 to 24 months;
- the coefficients $\beta_\ell$ form the lag-response, which is constrained to a penalized cubic P-spline basis of dimension k = 8 over ℓ ∈ {1,…,24} months, with the smoothness penalty supplied to bam via paraPen and selected data-adaptively by fREML.[23,24] Cross-bases are constructed with dlnm::crossbasis;[25,26]
- $\beta_\ell$ is the lag-specific coefficient of the primary $PM_{2.5}$ exposure;
- $\theta$ is the coefficient for GeneXpert diagnostic coverage, the proportion of notified TB cases in municipality i and year(t) with a GeneXpert result (Section S1.16);
- $g_{T(l)}$ and $g_{P(l)}$ are unpenalized natural-spline cross-basis lag- response functions (df = 3) for temperature and precipitation, fitted over the same 1–24 month lag window; and
- $\log(\text{Pop}_{it})$ is the log-population offset, using the annual population estimate for year(t).

The model was fitted via mgcv::bam(family=poisson(link="log"))with the smoothness parameter selected by fast restricted maximum likelihood fREML.[27] Poisson log-likelihood estimation under the conditional-mean specification is consistent for the regression coefficients without assuming equidispersion (Poisson quasi-maximum-likelihood).[28,29]

Effect estimates are reported for a fixed contrast of 14 additional high-exposure days. This value corresponds to the difference between the 95th and 25th percentiles of fire-event duration in Brazil, derived from the Global Fire Atlas of individual fire events, and represents the change from a low-exposure baseline to a high fire-activity level.[30] Because exposure enters the model as the monthly count of days exceeding 25 μg/m³ with a penalized spline over the lag dimension only (i.e., linear in the day-count), the contrast of 14 additional exceedance days is invariant to how those days are distributed within or across calendar months. The average monthly change should be read as a geometric-mean summary of the cumulative effect.

#### S2.5 Conley spatial HAC standard errors

Standard errors were computed using the *posthoc* spatial heteroskedasticity- and autocorrelation-consistent (HAC) estimator of Conley.[31] This estimator accounts for spatial correlation in regression residuals by weighting the contribution of observation pairs to the variance-covariance matrix by their geographic distance: pairs of municipalities within a specified distance cutoff receive non-zero weight, while pairs beyond the cutoff are treated as uncorrelated. The primary cutoff in our analysis was 300 km. Sensitivity analyses examined a cutoff of 500 km (Section S3.12).

#### S2.6 Penalized P-spline distributed lag model

We estimated the lag-response relationship with a penalized distributed lag model (DLM) using mgcv::bam with the paraPen argument in R.[27] The construction is:

i. A P-spline basis matrix B of dimension L × k is constructed over the lag indices $\ell = 1, 2, ..., 24$, where $L = 24$ is the number of lag positions and $k = 8$ the basis dimension.
ii. For each basis function $B_i(\ell)$, a cross-basis variable is defined as $CB_j(i, t) = \Sigma_\ell B_i(\ell) \times \text{Exposure}_{i, t-\ell}$.
iii. The k cross-basis variables enter bam as parametric terms, with the penalty matrix S from the P-spline basis applied via paraPen. The smoothing parameter is estimated via fast restricted maximum likelihood (fREML), so the data determine the degree of smoothness on the lag dimension.[27]
iv. A second-order difference penalty ($m = 2$) is used throughout, as recommended by Gasparrini and colleagues for distributed lag models.[24]
v. We retain the constant (level) component of the lag basis rather than imposing a sum-to-zero centering constraint, so the cumulative effect across the 1–24 month window remains estimable. This component is not penalized by the second-order difference penalty.

Lag-specific coefficients and pointwise 95% confidence intervals were recovered by projecting the estimated basis coefficients onto the original lag grid: $\beta(l) = B(l)'\gamma$ , where γ is the vector of estimated basis coefficients and $B(\ell)$ is the $\ell$-th row of the basis matrix. The cumulative effect across the 24-lag window is the row sum of the basis matrix multiplied by γ. We report the average per-lag-month effect (cumulative ÷ 24) as the headline summary.

The peak lag is identified as the argmax of the reconstructed lag-response curve $\beta(l) = B(l)' \cdot \hat{\gamma}$. Uncertainty in the peak was characterized by drawing 10 000 samples of the basis coefficients γ from a multivariate normal distribution centred at $\hat{\gamma}$ with covariance equal to the covariance matrix Vp returned by mgcv, reconstructing the lag-response curve for each draw and taking the argmax.[27] We report the median peak lag and the 2.5$^{th}$ and 97.5$^{th}$ percentiles of the resulting distribution as the 95% credible interval.

#### S2.7 Backward attributable-fraction g-computation

Attributable cases and attributable fractions were computed using the backward g-computation framework of Gasparrini and Leone adapted to distributed lag models.[32] For each municipality-month, we defined a counterfactual scenario in which exposure was set to zero high-exposure days across the entire 24-month lag window, with all other covariates and fixed effects held at their observed values. Attributable quantities were then computed by contrasting predictions under observed exposure against predictions under the counterfactual:

vi. Predicted cases under observed exposure: $\hat{y}_{it}$
vii. Predicted cases under zero exposure: $\hat{y}_{it}(0)$

viii. Attributable cases at municipality-month it: $AC_{it} = \hat{y}_{it} - \hat{y}_{it}(0)$

The primary estimand was the attributable fraction in the exposed (AFE) based on Cole and MacMahon's definition of attributable risk among the population exposed.[33] We defined AFE as the ratio of total attributable cases across exposed municipality-months (those with at least one high-exposure day in the 24-month lag window) to total observed cases in the same set:

$$AFE = \frac{\sum_{(i,t)\in\,exposed} AC_{it}}{\sum_{(i,t)} Y_{it}^{exposed}}$$

Biome-level AFEs, microregion-level attributable case counts, and year-level attributable case counts were computed by stratifying the numerator and denominator sums to the appropriate subset. Uncertainty was propagated by drawing 10,000 samples from the multivariate-normal posterior distribution of the basis coefficients implied by the Conley spatial HAC covariance matrix, recomputing AFE and attributable cases for each draw, and reporting 2.5$^{th}$ and 97.5$^{th}$ empirical percentiles as 95% confidence intervals. This procedure propagates both coefficient uncertainty and the spatial-correlation structure into the attributable estimates.

Microregion-level aggregation was used for the choropleth display (Figure 4 in the main text) rather than municipality-level aggregation. Municipality-level AFE ratios are noisy because the numerator and denominator can both be small in low-burden municipalities; aggregation to the microregion stabilizes the ratio for visualization without altering the underlying point estimates.

#### S2.8 Concentration-based DLNM specification

**Approach.** To complement the threshold-based primary analysis, we estimated $PM_{2.5}$ exposure-response associations were with a penalized distributed-lag non-linear model (DLNM).[25,26] The exposure-response dimension used a penalized P-spline (maximum basis dimension 8) and the lag-response dimension a natural cubic spline (4 degrees of freedom) over a 1-24 month lag window; the cross-basis penalty was built with cbPen() and its smoothing parameter selected by fast REML, yielding 12.6 effective degrees of freedom. Monthly TB notification counts were modelled with a Poisson log link with municipality, state-by-year, state-by-month, and state-by-pandemic-phase fixed effects and a log-population offset, and additional adjustment for temperature and precipitation cross-bases, and GeneXpert diagnostic-test coverage. Confidence intervals for the cross-basis terms were obtained from post-hoc Conley spatial heteroskedasticity- and autocorrelation-consistent (HAC) standard errors with a 300 km cutoff, computed on the Bayesian posterior covariance of the penalized fit.

The cumulative (overall) relative risk at a given $PM_{2.5}$ concentration is the exponential of the lag-specific log relative risks summed across lags 1-24 months, expressed relative to a reference concentration of 15 µg/m³ (at which the relative risk equals one by construction). For the consecutive-increase comparison, each increase from $x$ to $y$ µg/m³ was computed as a separate prediction referenced to its own lower bound a: although the point estimate is invariant to the reference concentration, the standard error depends on the spline difference between b and a. The 80% and 95% confidence intervals were reconstructed log-symmetrically from the Conley standard error of the cumulative log relative risk.

**Results.** The relationship between $PM_{2.5}$ concentrations and TB notifications is monotonically increasing and non-linear (Supplementary Fig. 16, Supplementary Fig. 17): it is essentially flat across the low concentrations where most municipality-months lie (shown by the rug) and increases progressively above roughly 20-25 µg/m³. The confidence band widens markedly in the upper tail, reflecting the sparse observations at high wildfire-related $PM_{2.5}$ concentrations. The effect of an identical +10 µg/m³ increment grows as the baseline concentration rises. Lowest increases fall in the data-dense part of the exposure distribution and is estimated precisely, whereas the 25-to-35 increase lies in a sparse, high-concentration range and is correspondingly uncertain. The full relative-risk surface is shown over the $PM_{2.5}$ (horizontal axis) by lag (vertical axis) plane, referenced to 15 µg/m³, so that the relative risk equals one along the vertical line at that concentration (Supplementary Fig. 18). Elevated risk is concentrated at high $PM_{2.5}$ and at lags of approximately 18 months, consistent with a delayed effect of smoke exposure on subsequent TB notification, with little departure from the null at low concentrations or short lags.

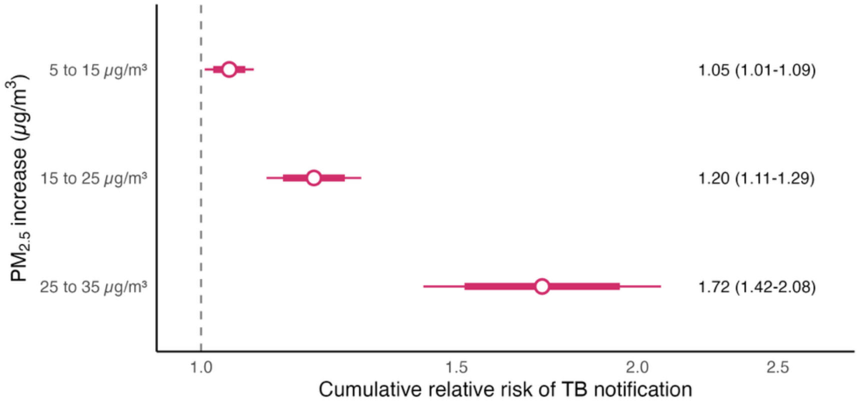


**Supplementary Fig. 16. Cumulative relative risk of municipality tuberculosis notification for successive increases in monthly wildfire-related $PM_{2.5}$ in Brazil, Jan 1, 2005 – Dec 1, 2023.** Each row shows the overall cumulative relative risk of TB notification, summed over lags 1-24 months, for a 10 µg/m³ increase in $PM_{2.5}$ referenced to the lower bound of the increase. Open points mark the central estimates; thick and thin bars show the 80% and 95% confidence intervals (Conley spatial HAC, 300 km cutoff); the vertical dashed line marks the null. $PM_{2.5}$ is the GFED4.1s wildfire-related monthly 90th-percentile metric. Estimates are from a penalized distributed-lag non-linear model of monthly TB notification counts (Poisson, log-population offset) with municipality, state-by-year, state-by-month, and state-by-pandemic-phase fixed effects, adjusted for temperature and precipitation cross-bases, and GeneXpert diagnostic-test coverage.

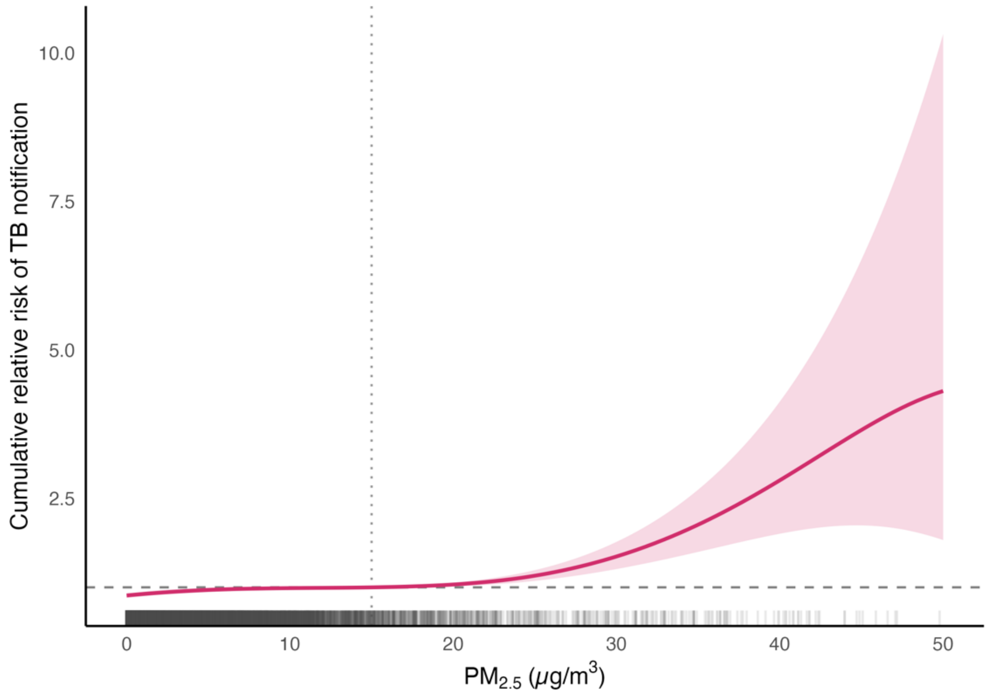


**Supplementary Fig. 17. Exposure-response relationships between $PM_{2.5}$ and TB notifications from the penalized distributed-lag nonlinear model in Brazil, Jan 1, 2005 – Dec 1, 2023.** The figure represents the overall cumulative relative risk of monthly $PM_{2.5}$ concentrations (90th-percentile) on tuberculosis notifications, with the reference exposure set to 15 µg/m³, summed over lags 1-24 months. Curves show the posterior estimate (line) with 95% confidence intervals (shaded band) derived from the Conley spatial HAC variance-covariance matrix (300 km cutoff). The vertical dotted line marks the reference exposure (15 µg/m³), at which the relative risk equals 1 by construction; the horizontal dashed line marks the null. The rug along the bottom shows the distribution of observed monthly $PM_{2.5}$ (a random sample of municipality-months). The model adjusts for temperature and precipitation cross-bases, diagnostic-test coverage and pandemic phase, with municipality, state-by-year and state-by-month fixed effects and a log-population offset.

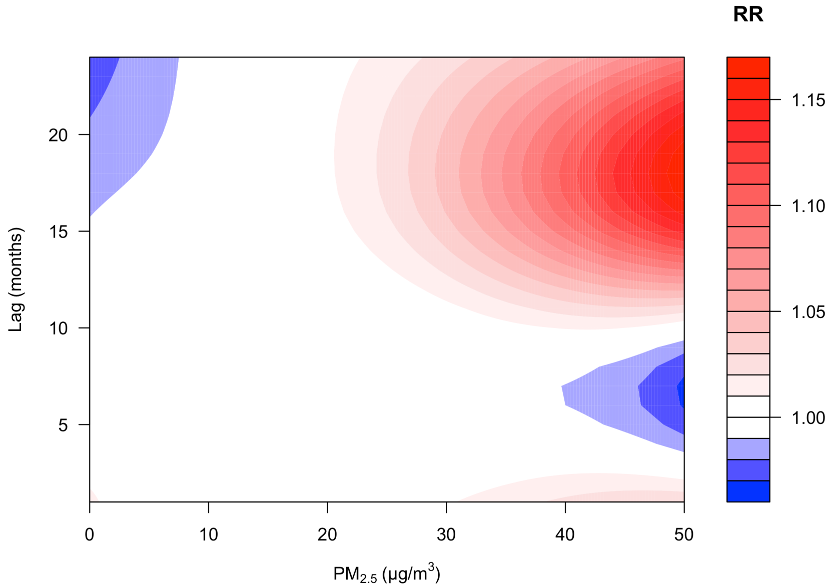


**Supplementary Fig. 18. Relative-risk surface for the relationship between monthly wildfire-related $PM_{2.5}$ and tuberculosis notification across exposure and lag.** The contour shows the relative risk of TB notification over the joint plane of monthly $PM_{2.5}$ (horizontal axis) and lag in months (vertical axis), referenced to 15 µg/m³ so that the relative risk equals one along the vertical line at that concentration; colour denotes the relative risk (key at right). $PM_{2.5}$ is the GFED4.1s wildfire-related monthly 90th-percentile metric. Estimates are from a penalized distributed-lag non-linear model of monthly TB notification counts (Poisson, log-population offset) with municipality, state-by-year and state-by-month fixed effects, adjusted for temperature and precipitation cross-bases, GeneXpert diagnostic-test coverage and pandemic phase; confidence intervals derive from Conley spatial HAC standard errors (300 km cutoff). Brazil, Jan 1, 2005 – Dec 1, 2023.

### S3. Sensitivity and robustness analyses

#### S3.1 Alternative $PM_{2.5}$ thresholds

The primary exceedance threshold of 25 µg/m³ is anchored to the Brazilian air pollution CONAMA 491/2018 *Padrão Final* 24-hour target. The choice of threshold could affect both effect magnitude (different exposure intensities) and statistical precision (different event frequencies). We re-estimated the primary penalized distributed lag model at thresholds of 15 µg/m³ (WHO interim target 4) and 35 µg/m³ (WHO interim target 1), with all other specifications held constant. Effect estimates were rescaled to the common 14-high-exposure-day contrast.

The lag-response shape was mostly preserved across all three thresholds (Supplementary Fig. 19), with a flatter curve for 15 µg/m³ and a more pronounced peak at 35 µg/m³. The median peak lag was 11 months at the 25 µg/m³ (IQR 11–12) and 35 µg/m³ (IQR 11–11) thresholds and 15 months (IQR 13–19) at the 15 µg/m³ threshold (Supplementary Fig. 20). Effect magnitude per 14-day contrast scaled with threshold (smaller at 15 µg/m³, larger at 35 µg/m³) consistent with a dose-response gradient (Figure 3A in the main text).

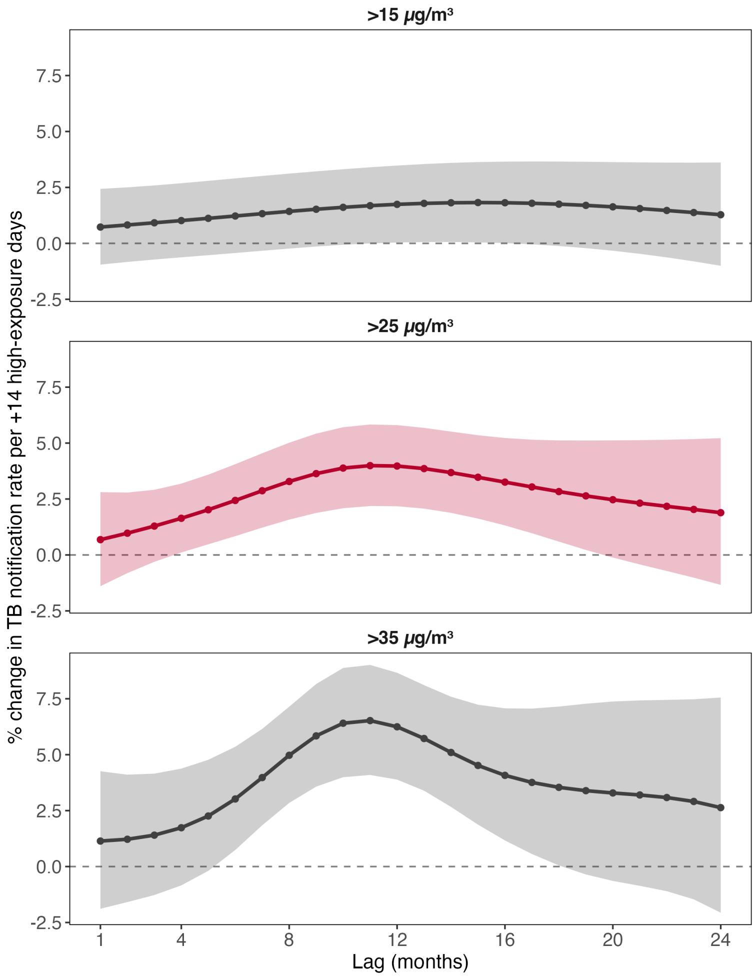


**Supplementary Fig. 19. Lag-response relationships between wildfire-related $PM_{2.5}$ and TB notification rates across thresholds in Brazil, Jan 1, 2005 – Dec 1, 2023.** Each panel shows the estimated percent change in the TB notification rate per +14 high-exposure days per month as a function of lag (in months), from the penalized-spline distributed-lag model for one $PM_{2.5}$ high-exposure threshold: (top) >15, (middle) >25, and (bottom) >35 µg/m³. Lines are posterior point estimates with points at each integer lag; shaded bands are pointwise 95% confidence intervals. The primary threshold (25 µg/m³) is drawn in dark pink; the sensitivity thresholds (15 / 35 µg/m³) in dark grey.

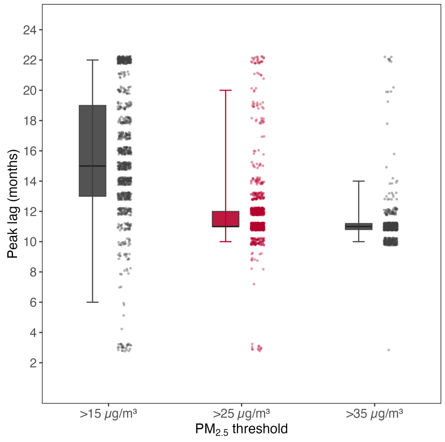


**Supplementary Fig. 20. Peak-lag distribution for the effect of wildfire-related $PM_{2.5}$ on TB notification rates across high-exposure thresholds in Brazil, Jan 1, 2005 – Dec 1, 2023.** For each of the three $PM_{2.5}$ high-exposure thresholds (>15, >25, and >35 µg/m³), 10,000 coefficient vectors were drawn from the multivariate normal posterior of the fitted spline coefficients (mean = fitted coefficients; covariance = variance matrix of fitted coefficients) and mapped to a lag-response curve via the lag basis. The peak lag of each draw is the lag (in months) at which its lag-response is maximal. Each half-boxplot shows the 25th, 50th (thick line), and 75th percentiles (box) with whiskers at the 2.5th and 97.5th percentiles of the peak-lag posterior; jittered points to the right show a subsample of the individual posterior draws. The primary threshold (25 µg/m³) is drawn in dark pink; the sensitivity thresholds (15 / 35 µg/m³) in dark grey.

#### S3.2 QFED2.5 alternative fire-emission inventory

Wildfire-related $PM_{2.5}$ concentrations depend on the fire-emission inventory, which affects burned area, fuel consumption, and emission factors. GFED4.1s (primary) derives emissions from satellite-observed burned area; QFED2.5 (secondary) scales emissions to MODIS-retrieved aerosol optical depth and tends to assign higher fire-$PM_{2.5}$. In this sensitivity analysis, we assess how our main results are affected by the choice of the fire-emission inventory.

**Approach.** We compared the two inventories at both the concentration level and the high-exposure-day level, then re-estimated the primary penalized P-spline distributed lag model with QFED2.5-derived wildfire $PM_{2.5}$ in place of GFED4.1s, holding all other specifications constant. The primary 25 µg/m³ threshold was retained for direct comparability. Agreement was summarized by Pearson and Spearman correlations across all paired municipality-months and by inspection of the bivariate distribution.

**Result.** General spatiotemporal patterns were preserved across inventories (compare Supplementary Fig. 3 and Supplementary Fig. 21). The time series of the monthly-mean concentrations tracked one another closely across 1,390,837 paired municipality-months (Pearson r 0.77; Spearman ρ 0.80; Supplementary Fig. 22B). QFED2.5 assigned higher fire-$PM_{2.5}$ than GFED4.1s across most of the distribution, with the two converging towards the 1:1 line only at the highest concentrations. Translating to the primary exposure metric, the inventories agreed more closely on high-exposure days above 25 µg/m³ (Pearson r 0.76; Spearman ρ 0.88; Supplementary Fig. 23B), reflecting convergence of the two inventories where threshold exceedances occur. The largest discrepancies in exceedance days were concentrated in biomes with lower fire activity (Mata Atlântica, Caatinga), whereas the high-burden Amazônia and Pantanal biomes, those that dominate the national exposure signal, showed close inter-inventory agreement (Supplementary Fig. 22C, Supplementary Fig. 23C).

The lag–response shape was consistent between inventories at the 25 µg/m³ threshold (Supplementary Fig. 24A). Both inventories yielded peak-lag distributions centred near 10–11 months (Supplementary Fig. 24C). The QFED2.5 estimate was lower in magnitude than the GFED4.1s primary estimate (average per-lag-month change of approximately 1% versus 3%; Supplementary Fig. 24B), and its 95% confidence interval included the null (+0.9%, 95% CI −0.2 to +1.9, versus +2.7%, 95% CI +0.9 to +4.5), although the confidence intervals of the two estimates overlapped. The preservation of the lag–response shape under the alternative fire-emission inventory indicates that the timing and shape of the association are not specific to GFED4.1s.

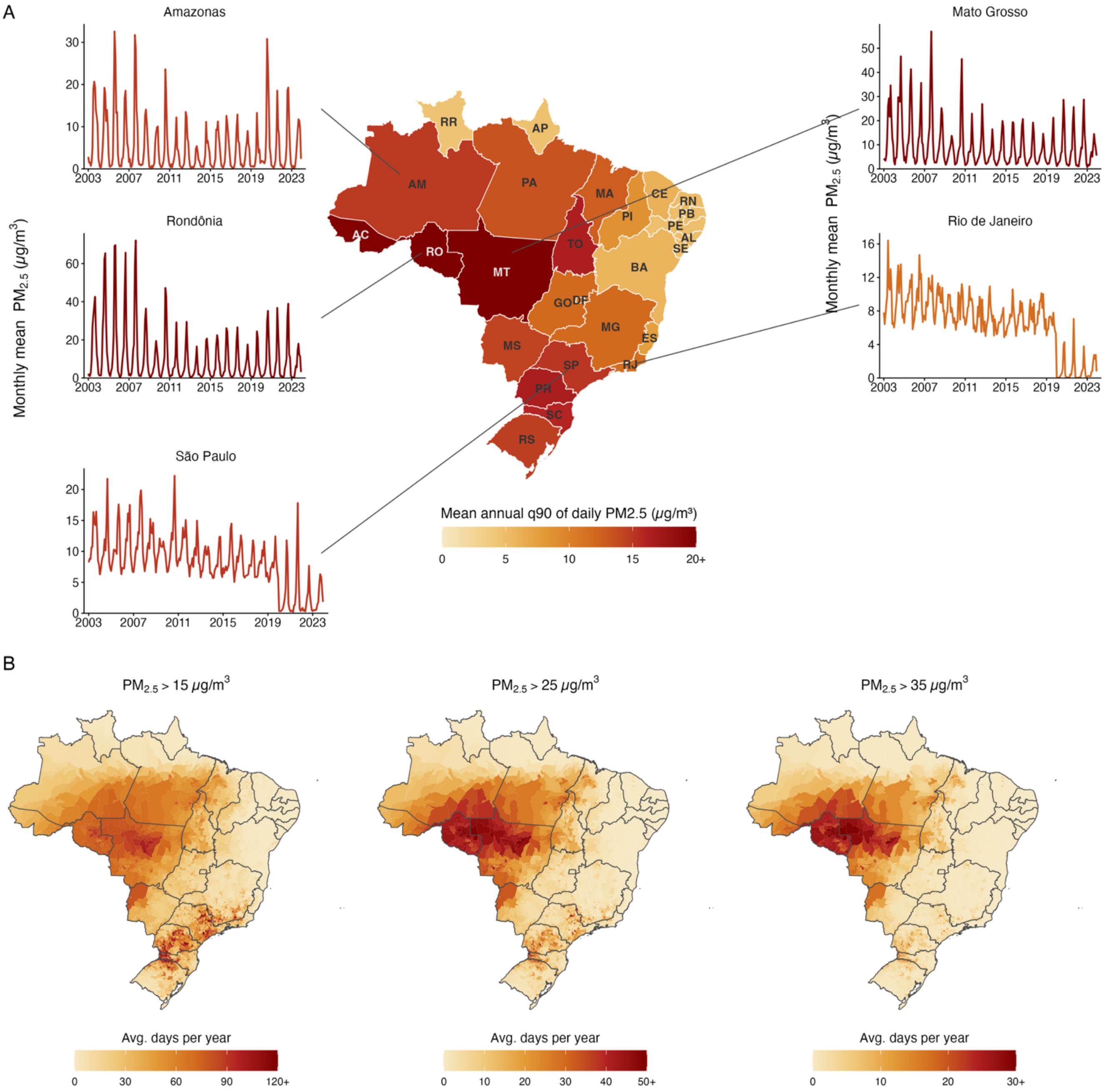


**Supplementary Fig. 21. Spatiotemporal patterns of QFED2.5 wildfire-related $PM_{2.5}$ in Brazil, Jan 1, 2003 – Dec 1, 2023.** (A) Choropleth of the mean of annual 90th-percentile daily wildfire-related $PM_{2.5}$ per state (μg/m³). For each state-year, the 90th percentile is computed over all daily municipality-level $PM_{2.5}$ observations within that state; the map shows the mean of these annual q90 values across the study period. Time-series insets show the monthly mean of QFED2.5 wildfire-related $PM_{2.5}$ for selected states (Amazonas, Rondônia, São Paulo, Mato Grosso, Rio de Janeiro), where each monthly value is the monthly mean over all municipality × day observations within that state-month. (B) Municipality-level choropleth of the average number of days per year with wildfire-related $PM_{2.5}$ concentrations exceeding 15, 25, and 35 μg/m³. Monthly high-exposure day counts per municipality were summed to annual totals and then averaged across years. Each panel uses an independently scaled colourbar; the upper limit is set to the 99th percentile of municipality values (rounded up to the nearest 10). Municipalities above this cap are rendered in the top colour of the ramp, and the top legend tick is annotated with "+" to indicate this.

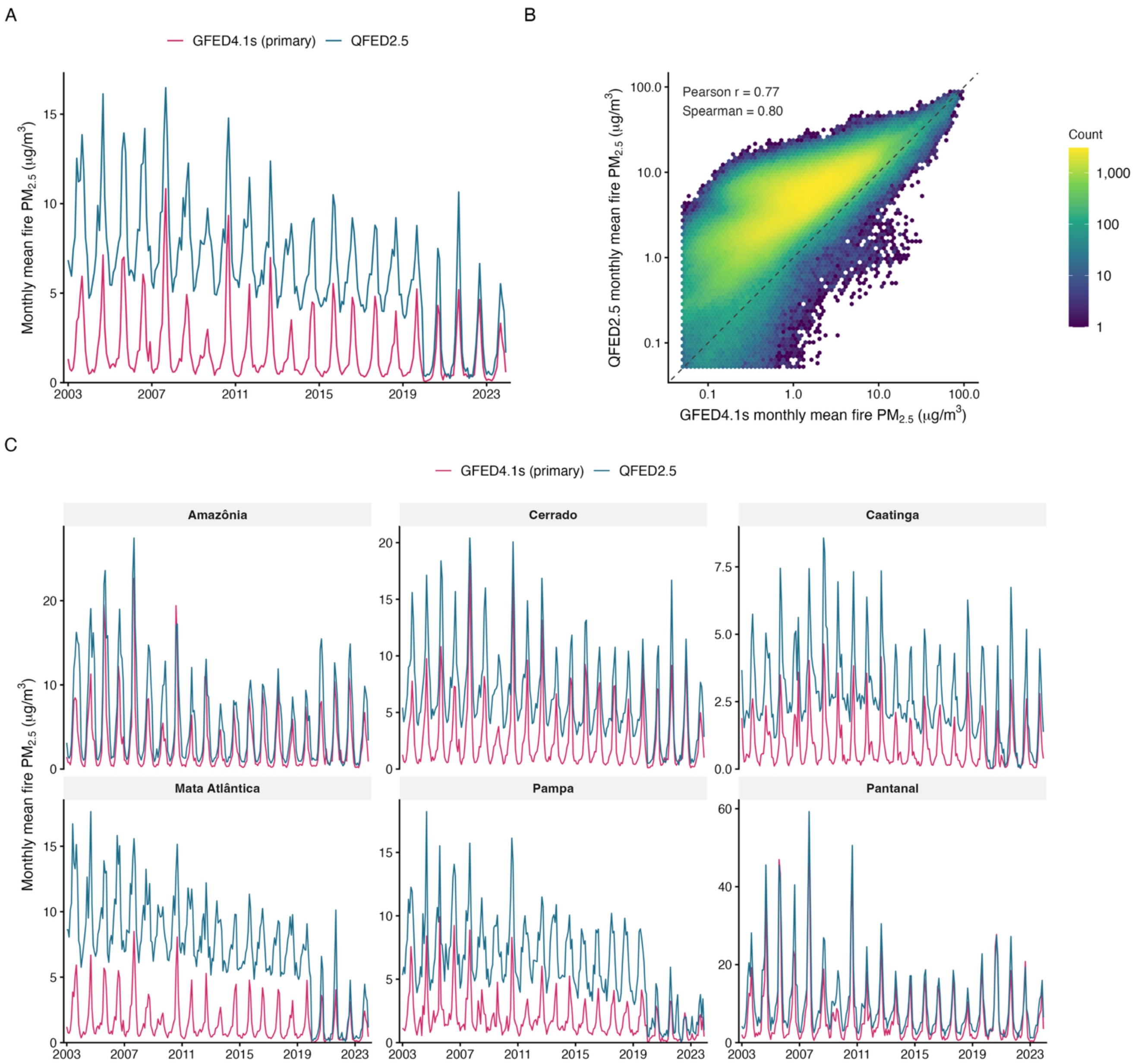


**Supplementary Fig. 22. Comparison of wildfire-related $PM_{2.5}$ concentrations from the two fire-emissions inventories GFED4.1s (primary exposure) and QFED2.5 (sensitivity analysis), across the analytic sample in Brazil**, Jan 1, 2003 – Dec 1, 2023. (A) Brazil-wide monthly mean fire-$PM_{2.5}$ (μg/m³) for each inventory, computed as the population-weighted average of municipality monthly means across the analytic sample (n = 5,544 municipalities). Weights are annual IBGE municipality population estimates and are identical for both inventories. (B) Paired agreement between the inventories at the municipality-month level: each cell shades the number of municipality-months (log scale) at a given combination of GFED4.1s (x-axis) and QFED2.5 (y-axis) monthly mean fire-$PM_{2.5}$; both axes are on a log scale. The dashed line is the 1:1 line. Correlations are computed over all municipality-months with a positive monthly mean in both inventories; for display the axes are floored at 0.05μg/m³ (38,082 of 1,390,837 municipality-months fall below this floor in one inventory and are omitted from the hexbin only). (C) Monthly mean fire-$PM_{2.5}$ by IBGE biome (Amazonia, Cerrado, Caatinga, Mata Atlantica, Pampa, Pantanal) for each inventory, computed as the population-weighted average of municipality monthly means across analytic-sample municipalities assigned to each biome. Y-axes are independent across biome panels.

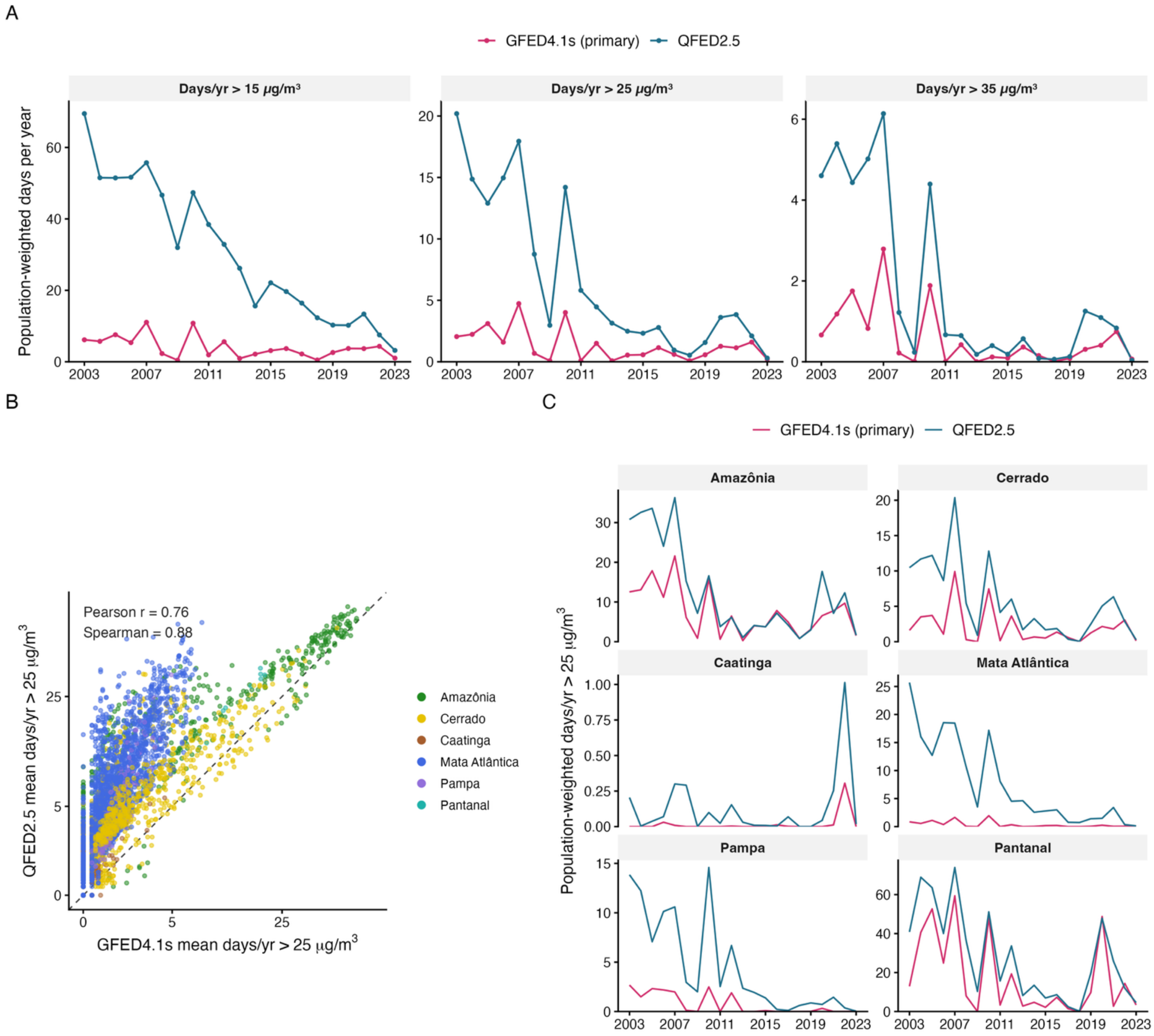


**Supplementary Fig. 23. Comparison of high-exposure days from the two fire-emissions inventories, GFED4.1s (primary exposure) and QFED2.5 (sensitivity analysis), across the analytic sample in Brazil, Jan 1, 2003 – Dec 1, 2023.** High-exposure days are days on which wildfire-related $PM_{2.5}$ exceeds a threshold; 25 µg/m³ is the study's primary exceedance threshold. (A) Brazil-wide population-weighted mean number of days per year exceeding 15, 25, and 35 µg/m³ for each inventory, by calendar year. For each municipality-year the monthly exceedance-day counts were summed to an annual total; the national value is the population-weighted average of these annual totals across the analytic sample (n = 5,544 municipalities). Weights are annual IBGE municipality population estimates, identical for both inventories. Y-axes are independent across thresholds (note scales). (B) Paired municipality-level agreement in the mean annual number of days above 25 µg/m³ (averaged over 2003-2023): GFED4.1s (x-axis) vs QFED2.5 (y-axis); both axes are on a square-root scale. Each point is one municipality, coloured by IBGE biome. The dashed line is the 1:1 line. (C) Population-weighted mean days per year above 25 µg/m³ by IBGE biome (Amazonia, Cerrado, Caatinga, Mata Atlantica, Pampa, Pantanal) for each inventory, by calendar year.

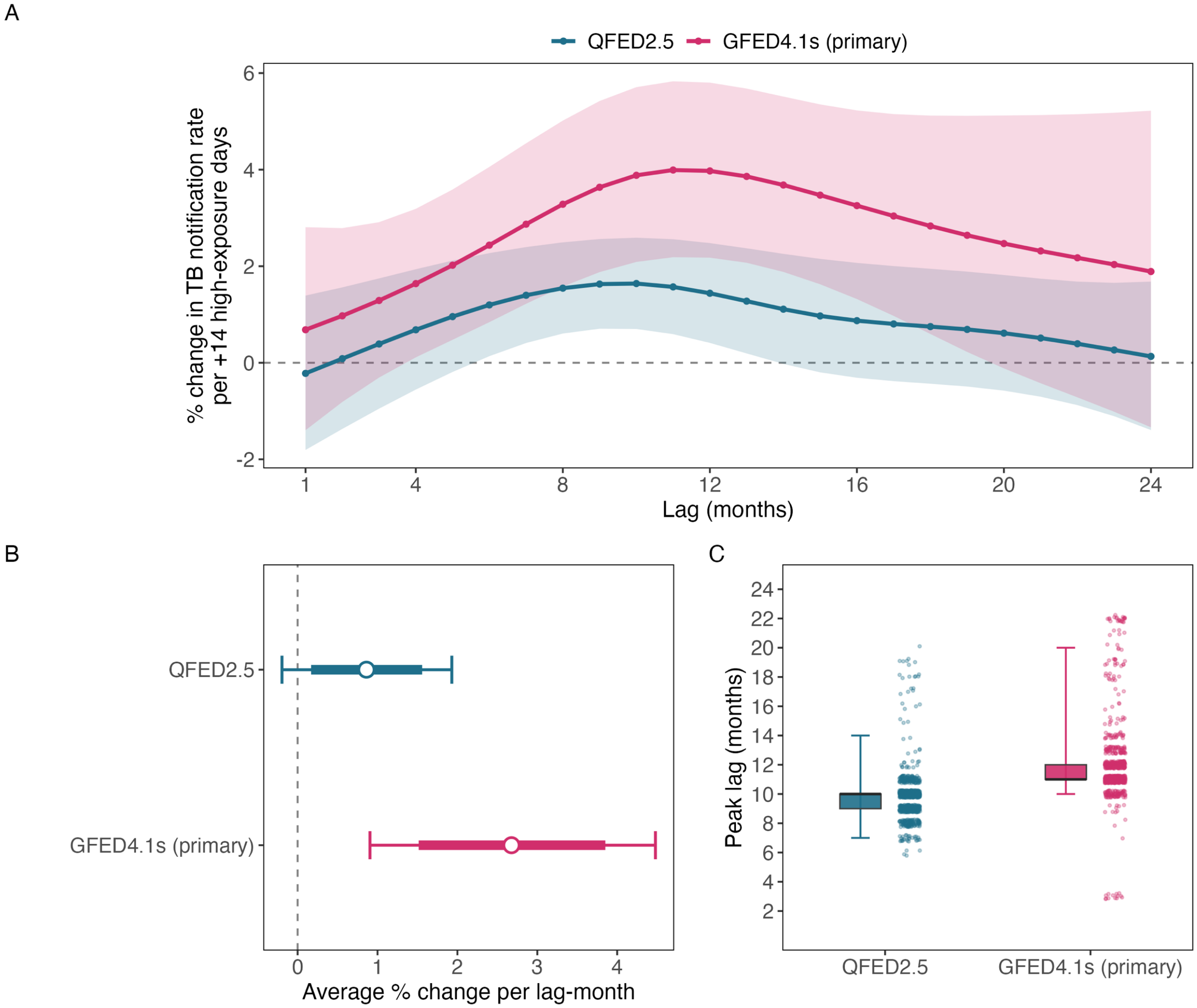


**Supplementary Fig. 24. Sensitivity of the lag-response relationship between wildfire-related $PM_{2.5}$ and TB to the choice of $PM_{2.5}$ inventory in Brazil, Jan 1, 2005 – Dec 1, 2023.** All three panels compare two penalized-spline distributed-lag models that are identical in every modelling choice (>25 µg/m³ high-exposure days, penalized lag basis k=8, lags 1-24 months, municipality + state-year + state-month + state-pandemic-phase fixed effects, log-population offset, Conley HAC standard errors at 300 km) and differ only in the wildfire-related $PM_{2.5}$ input product: QFED2.5 versus GFED4.1s (primary exposure inventory). They therefore isolate sensitivity to exposure measurement. (A) Lag-response curves: estimated percent change in the TB notification rate per +14 high-exposure days per month (the P25-to-P95 exposure contrast) as a function of lag (in months), overlaid for the two inventories. Lines are posterior point estimates with points at each integer lag; shaded bands are pointwise 95% confidence intervals. (B) Average monthly effect: the average monthly percent change in the TB notification rate across the 1–24-month lag window for each inventory, shown as the point estimate with its 80% (thick bar) and 95% (thin bar) confidence intervals (dashed line at no effect). (C) Peak-lag distribution: posterior distribution of the peak lag (argmax of the lag-response curve) for each inventory. 10,000 coefficient vectors were drawn from the Bayesian posterior of each fit and mapped to a lag-response curve via the lag basis; the peak lag of each draw is the lag (in months) at which its lag-response is maximal. Each half-boxplot shows the 25th, 50th (thick line), and 75th percentiles (box) with whiskers at the 2.5th and 97.5th percentiles; jittered points to the right show a subsample of individual posterior draws.

#### S3.3 $NO_2$ co-pollutant adjustment

Wildfire-related $PM_{2.5}$ could be acting as a marker for general urban combustion pollution, which would imply that the estimated effect reflects fossil-fuel rather than biomass-burning sources.

**Approach.** We re-estimated the primary penalized distributed lag model with $NO_2$ threshold-exceedance variable (at 25 µg/m³) entered through a matched 1–24-month distributed lag structure.

**Result.** The $PM_{2.5}$ lag-response was similar in magnitude and shape after NO2 adjustment (Supplementary Fig. 25). This suggests that the wildfire $PM_{2.5}$ signal is distinct from general urban combustion, consistent with the markedly different spatial signatures of the two pollutants (urban centres for NO2; agricultural frontier for wildfire-related $PM_{2.5}$).

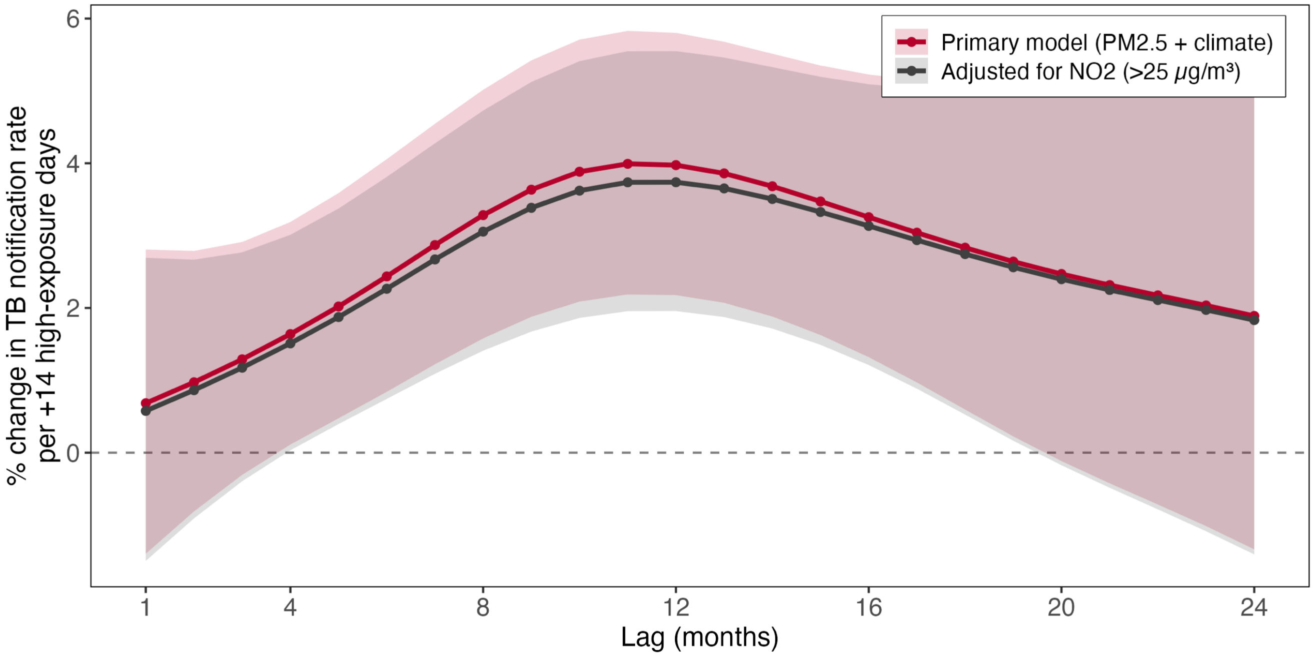


**Supplementary Fig. 25. Robustness of the wildfire-related $PM_{2.5}$ lag-response to NO2 co-pollutant adjustment, Brazil, Jan 1, 2005 – Dec 1, 2023.** Estimated percent change in the TB notification rate per +14 high-exposure days per month as a function of lag (in months), from the primary penalized-spline distributed-lag model for wildfire-related $PM_{2.5}$ high-exposure days (>25 µg/m³). The dark-pink curve is the primary model adjusted for temperature and precipitation only; the grey curve is the same $PM_{2.5}$ lag-response after additionally adjusting for NO2 high-exposure days (>25 µg/m³) entered as a second penalized distributed-lag term.

##### S3.4 $O_3$ co-pollutant adjustment

Wildfire-related smoke is a chemically complex mixture, and ozone is formed photochemically downwind of fires from co-emitted precursors (nitrogen oxides and volatile organic compounds). Because gas-phase $O_3$ shares meteorological and emission drivers with wildfire smoke, we examined whether the estimated $PM_{2.5}$-TB effect might partly reflect co-varying ozone rather than particulate matter itself.

**Approach.** We re-fitted the primary penalized-spline distributed-lag model and added a second penalized distributed-lag term for monthly gas-phase $O_3$ high-exposure days (days with $O_3$ > 60 µg/m³.

**Results.** The wildfire-$PM_{2.5}$ lag-response relationship was essentially unchanged by O3 adjustment (Supplementary Fig. 26). The unimodal curve retained its peak at a 11-month lag, at +4.0% (95% CI 2.2 to 5.8) per 14 additional high-exposure days per month in the primary model and +3.9% (95% CI 2.1 to 5.8) after O3 adjustment; the two curves differed by at most 0.12 percentage points at any lag. Adjustment for co-varying gas-phase ozone did not attenuate the association between wildfire-related-$PM_{2.5}$ and TB, suggesting that photochemical ozone is unlikely to explain the result.

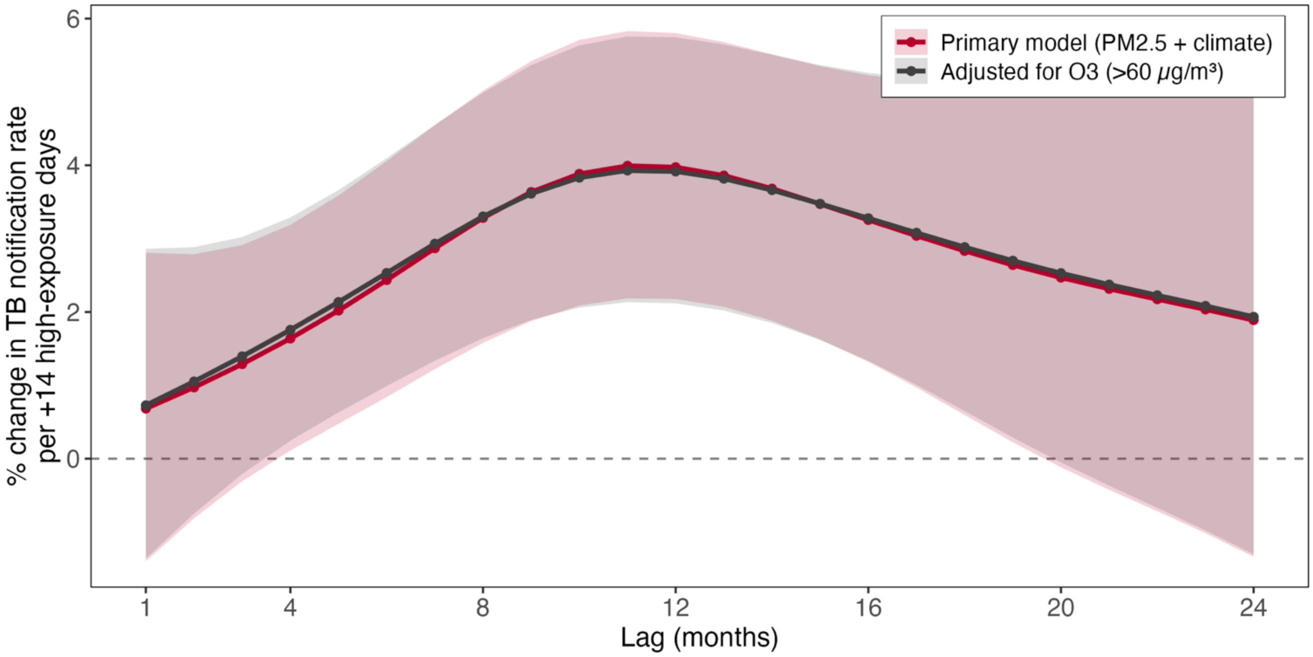


**Supplementary Fig. 26. Robustness of the wildfire-related $PM_{2.5}$ lag-response to gas-phase ozone (O3) co-pollutant adjustment, Brazil, Jan 1, 2005 – Dec 1, 2023.** Estimated percent change in the TB notification rate per 14 additional high-exposure days per month as a function of lag (in months), from the primary penalized-spline distributed-lag model for wildfire-related $PM_{2.5}$ high-exposure days (>25 μg/m³). The dark-pink curve is the primary model adjusted for temperature and precipitation only; the grey curve is the same $PM_{2.5}$ lag-response after additionally adjusting for gas-phase ozone high-exposure days (>60 μg/m³) entered as a second penalized distributed-lag term. Lines are posterior point estimates with points at each integer lag; shaded bands are pointwise 95% confidence intervals. Models are penalized-spline distributed-lag Poisson models with municipality, state-by-year, state-by-month, and state-by-pandemic-phase fixed effects, a log-population offset, and Conley spatial HAC standard errors (300 km).

#### S3.5 Drought and SPEI adjustment

Drought is a plausible shared cause of both wildfire activity and tuberculosis dynamics. Dry conditions promote fire spread and elevated smoke $PM_{2.5}$ and may independently affect TB through pathways such as food insecurity, undernutrition, and population displacement.

**Approach.** To assess sensitivity to this pathway, we re-estimated the primary model with additional adjustment for the 12-month Standardized Precipitation–Evapotranspiration Index. SPEI-12 integrates precipitation and evapotranspiration over the preceding 12 months, indexing the sustained water balance that governs both fire-fuel dryness and the slower socioeconomic and nutritional pathways through which drought may affect TB. SPEI-12 was entered as a 1–24-month natural-spline distributed-lag term, paralleling the temporal structure imposed on the exposure, with all other elements of the primary specification remained unchanged.

**Results.** The lag–response curve was essentially unchanged after SPEI-12 adjustment (Supplementary Fig. 27A), with overlapping confidence bands across the full lag window and a near-identical peak. The average monthly effect were similar after adjustment (+2.7% [95% CI 0.9 to 4.5] in the main analysis and +2.5% [95% CI 1.0 to 4.0] after adjustment; panel B). The posterior distribution of the peak lag was likewise stable (panel C): 11 months (95% interval 10–20) in the main analysis and 11 months (95% interval 9–22) after adjustment. The results suggest that the association is unlikely driven by drought-mediated confounding.

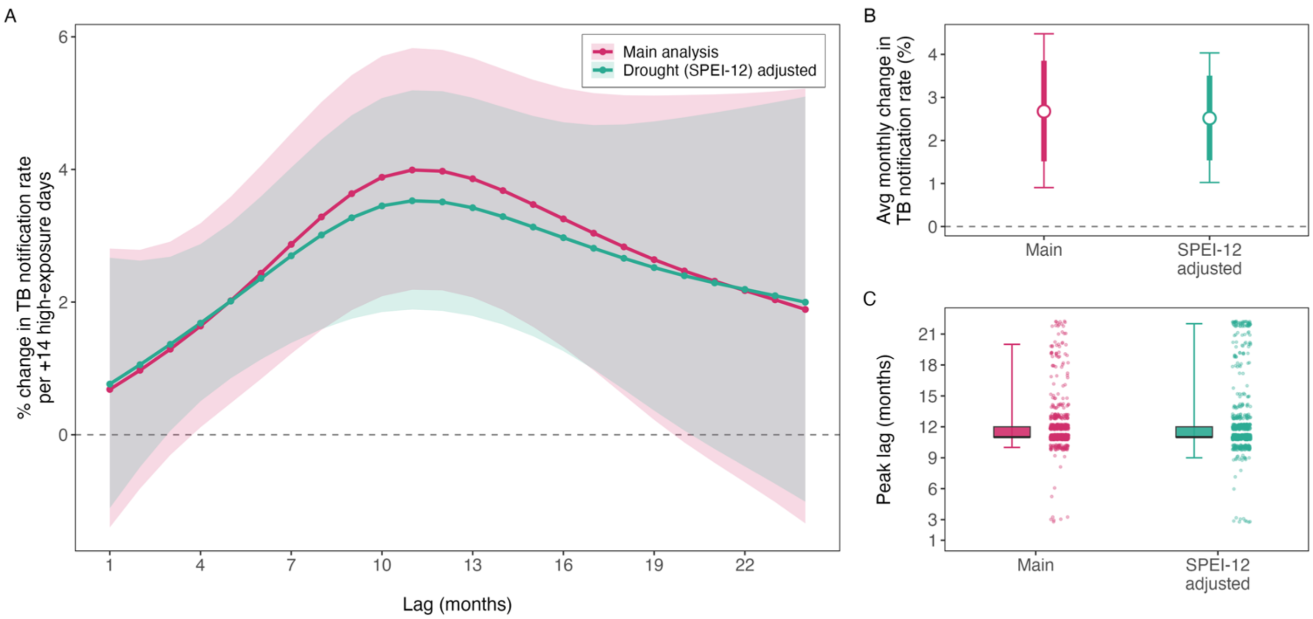


**Supplementary Fig. 27. Robustness of lag-response relationship between wildfire-related $PM_{2.5}$ and TB notification rates to adjustment for drought (SPEI-12).** Penalized-spline distributed-lag model ($PM_{2.5}$ >25 µg/m³ exceedance days, GFED4.1s), Brazil, Jan 1, 2005–Dec 1, 2023, with municipality, state×year, state×month, state×pandemic-phase fixed effects and Conley spatial HAC standard errors (300 km cutoff). (A) Lag–response curves for the main climate-adjusted analysis (temperature and precipitation) and a sensitivity analysis additionally adjusting for the 12-month Standardized Precipitation–Evapotranspiration Index as a 1–24-month natural-spline distributed-lag term. Lines show the mean percent change in TB notification rates per additional 14 high-exposure days at each lag; shaded bands are 95% Conley confidence intervals; the dashed line indicates no effect. (B) Average monthly effect for each model, defined as the geometric mean of the per-month rate ratios across the 24-month lag window, expressed as percent change per +14 high-exposure days. Thick bars show the 80% interval and thin bars the 95% interval; the dashed line indicates no effect. (C) Posterior distribution of the peak lag for each model, obtained by Monte Carlo sampling (N = 10,000) of the model coefficients, reconstructing the lag–response curve for each draw, and taking the argmax. Boxplots show the median and interquartile range, with whiskers at the 2.5th and 97.5th percentiles; points are a subsample of retained draws.

#### S3.6 Agricultural-expansion adjustment

Agricultural land use is closely linked to fire activity in Brazil and may also capture rural socioeconomic conditions, healthcare access, and population mobility that influence TB notification independently of wildfire smoke. Although municipality and state-by-year fixed effects account for time-invariant differences between municipalities and broad state-level temporal changes, they may not fully capture within-municipality changes in land use or local agricultural frontier expansion occurring within the same state-year. We therefore conducted a sensitivity analysis using municipal farming intensity, and its year-over-year change, as proxies for agricultural land-use change and frontier expansion. Because agricultural expansion may also contribute to the generation of wildfire-related $PM_{2.5}$, this analysis is interpreted as a conservative robustness check rather than as adjustment for a conventional confounder.

**Approach.** We re-estimated the primary penalized-spline distributed-lag model on the farming-restricted analytic sample, defined as municipality-months with available MapBiomas land-cover data and non-missing farming covariates. We adjusted for two land-use covariates: the municipal proportion of land classified as farming and the year-to-year change in that proportion. Farming was defined using the Level 1 Farming class from MapBiomas Collection 10. Both variables were included at annual lags 1 and 2, corresponding to the 1–24-month lag window used for wildfire-related $PM_{2.5}$. This allowed the model to account for recent changes in agricultural land use, rather than only the level of farming in a given year. The baseline and farming-adjusted models were fit to the same set of municipality-months, so any difference between estimates reflects the addition of the farming covariates rather than a change in the analytic sample. We compared the full lag-response curve, the average monthly effect, defined as the geometric mean of the lag-specific rate ratios across lags 1–24, and the posterior distribution of the peak lag estimated through Monte Carlo sampling of the model coefficients.

**Results.** The lag–response curve was essentially unchanged after adjustment (Supplementary Fig. 28). The average monthly effect showed little change (+2.7% [95% CI 0.9 to 4.5] at baseline and +2.6% [95% CI 0.9 to 4.4] after adjustment; panel B). The unimodal lag–response and its peak near 11 months were retained (panel C). The persistence of the association under this adjustment suggests that the main findings are not explained solely by land-use-related differences across municipalities.

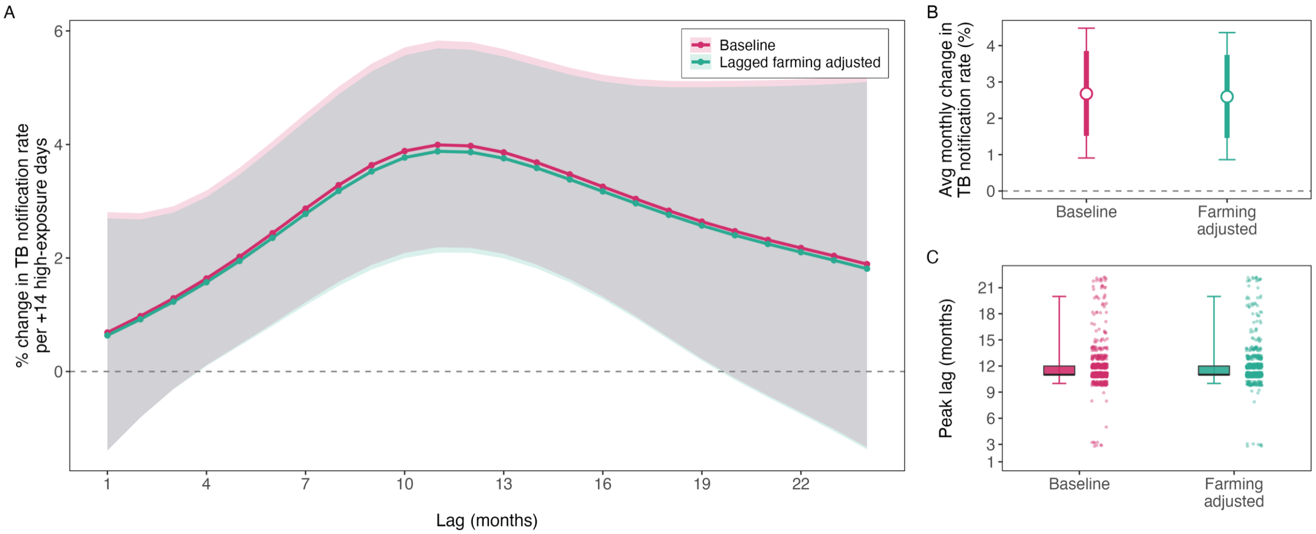


**Supplementary Fig. 28. Agricultural-expansion robustness analysis of the wildfire-related $PM_{2.5}$–tuberculosis lag–response to adjustment for the lagged farming proportion and its year-over-year change.** Penalized-spline distributed-lag model ($PM_{2.5}$ > 25 µg/m³ exceedance days, GFED4.1s), Brazil, Jan 1, 2005 – Dec 1, 2023, with municipality, state × year, and state × month fixed effects and Conley spatial HAC standard errors (cutoff 300 km). (A) Lag–response curves for the baseline model and the sensitivity analysis additionally adjusting for the municipal farming proportion (MapBiomas Collection 10) and its year-over-year change, each entered at year-lags 1–2 (spanning the 1–24-month $PM_{2.5}$ window at annual resolution). Lines are the mean percent change in TB notification rate per +14 additional high-exposure days at each lag; shaded bands are 95% Conley confidence intervals. The dashed line indicates no effect. (B) Average monthly effect for each model: the geometric mean of the per-month rate ratios across the 24-month lag window, expressed as percent change in TB notification rate per +14 high-exposure days. Thick bars show the 80% interval and thin bars the 95% interval, both from Conley spatial HAC standard errors. The dashed line indicates no effect. (C) Posterior distribution of the lag at which the effect peaks for each model. Peak lags were obtained by Monte Carlo sampling (N = 10,000) of the model coefficients, reconstructing the lag–response curve for each draw, and taking the argmax. Boxplots show the median and interquartile range, with whiskers at the 2.5th and 97.5th percentiles; points are a subsample of retained draws.

#### S3.7 Microregion aggregation

We repeated the analysis after aggregating municipalities to microregions to assess whether the lag–response curve was sensitive to the spatial unit of analysis. This addresses potential small-area surveillance noise, modifiable areal unit effects, and cross-municipality spillover in smoke transport, TB transmission, care-seeking, or notification. Although Conley HAC standard errors account for spatial correlation in inference, they do not change the point estimates.

**Approach.** To assess this, we refitted the penalized-spline distributed-lag model after collapsing observations to IBGE micro-regions (N = 558), aggregating the high-exposure-day exposure as a population-weighted mean of the already-constructed municipality-level variable and replacing within-municipality with within-microregion fixed effects.

**Results.** At the primary 25 µg/m³ threshold, the average monthly effect was +3.5% (95% CI +1.4 to +5.5) at micro-region resolution, compared with +2.7% (95% CI +0.9 to +4.5) at municipality resolution; the posterior median peak lag was 11 months under both units (Supplementary Fig. 29). The unimodal shape, peak timing, and direction and magnitude of the association were preserved after spatial aggregation across all three thresholds, suggesting that the lag–response pattern was not driven by small-area surveillance noise or by the choice of spatial unit.

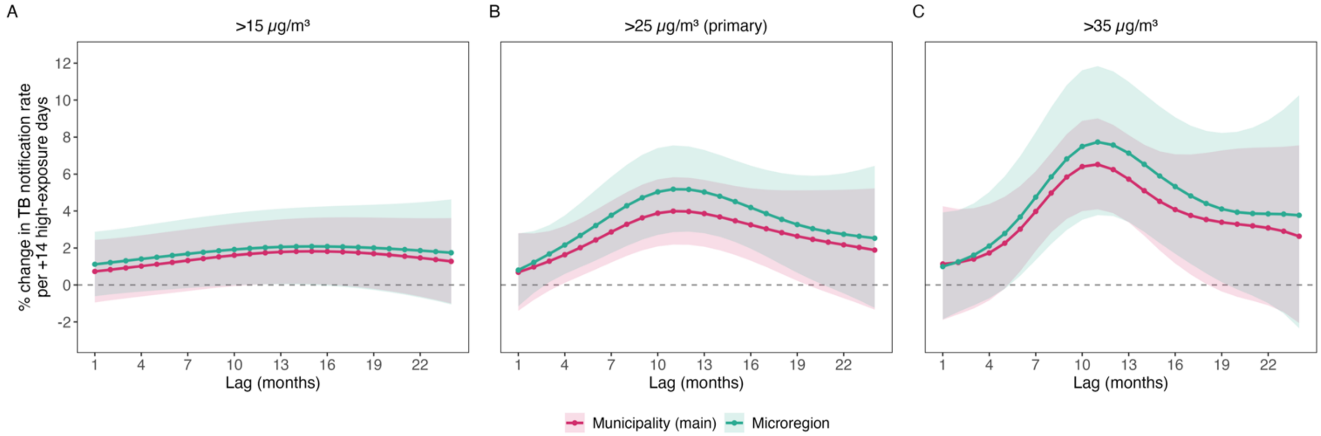


**Supplementary Fig. 29. Micro-region robustness analysis of the wildfire-related $PM_{2.5}$–tuberculosis lag–response.** Penalized-spline distributed-lag models (GFED4.1s high-exposure days, lags 1–24 months), Brazil, Jan 1, 2005–Dec 1, 2023, with state × year and state × month fixed effects, a log-population offset, and Conley spatial HAC standard errors (cutoff 300 km). The main analysis (pink) fits within-municipality fixed effects; the sensitivity analysis (teal) collapses observations to IBGE micro-regions and fits within-microregion fixed effects (N = 558 micro-regions). Each panel overlays the two spatial units at a single high-exposure threshold: (A) >15 µg/m³, (B) >25 µg/m³ (the primary analysis), and (C) >35 µg/m³. Lines are the mean percent change in TB notification rates per +14 additional high-exposure days at each lag; shaded bands are 95% Conley confidence intervals. The dashed horizontal line indicates no effect; the y-axis is shared across (A)–(C). Peak lags are obtained by Monte Carlo sampling (N = 10,000) of the model coefficients, reconstructing the lag–response for each draw and taking the argmax.

#### S3.8 Bacteriologically confirmed cases only

Our primary outcome is all notified tuberculosis cases (incident and recurrent). In Brazil's SINAN notification system, a substantial share of TB is diagnosed clinically or radiographically rather than confirmed microbiologically, on the basis of symptoms, chest imaging, contact history, or response to empirical therapy. Clinically diagnosed cases are more sensitive than laboratory-confirmed cases to health-system factors, e.g., diagnostic suspicion and care-seeking. Smoke-related respiratory symptoms could increase clinical investigations and presumptive diagnoses. If so, part of the estimated $PM_{2.5}$–TB association could reflect smoke-driven case ascertainment rather than a genuine increase in active TB. Restricting to bacteriologically confirmed cases tests this possibility.

**Approach.** We re-fit the primary model on bacteriologically confirmed cases only, using two definitions. Under the first definition, a case was classified as confirmed if it was positive on any of sputum culture, culture of other (extra-pulmonary) material, or the rapid molecular Xpert MTB/RIF assay. This definition prioritized specificity, because these methods identify *M. tuberculosis* directly, whereas smear microscopy cannot distinguish it from non-tuberculous mycobacteria and its quality may vary between laboratories. The second definition, which reflects the broader WHO case definition, additionally counted a positive smear on the first sputum sample or on other material. Missing values on a confirmation field were treated as not confirmed. The confirmed-case counts were aggregated to the same municipality-month panel. Every other part of the model was kept identical to the primary analysis.

**Results.** Culture- or Xpert-confirmed cases made up 24.5% of notified cases in the analysis sample, and cases confirmed by culture, Xpert or smear made up 63.9%; the counts are correspondingly sparser (92.3% and 80.4% of municipality-months, respectively, had zero confirmed cases). The culture or Xpert share rose from 7.8% in 2003 to 49.0% in 2023, whereas the share that also included smear rose from 59.0% to 70.5% (Supplementary Fig. 30).

The association was robust to both case definitions (Supplementary Figs. 31 and 32). The lag-response under the culture, Xpert or smear definition remained unimodal-shaped and positive throughout, with confidence intervals excluding the null around the peak. The timing was essentially unchanged: the effect peaked at a lag of 12 months for confirmed cases (posterior median 12 months, 95% interval 3–19) versus 11 months for all notified cases (median 11, 95% interval 10–20; panel C). Under the culture or Xpert definition, the lag-response was flatter and the timing of the peak less well identified (posterior median 16 months, 95% interval 3–22). The average monthly effect was +2.5% (95% CI +0.1 to +5.0) under the culture, Xpert or smear definition and +5.6% (+1.6 to +9.7) under the culture or Xpert definition per additional 14 high-exposure days, versus +2.7% (+0.9 to +4.5) in the primary analysis (Supplementary Figs. 31 and 32, panel B). The estimate did not attenuate when restricted to microbiologically confirmed disease under either definition, arguing against differential clinical ascertainment as the main explanation.

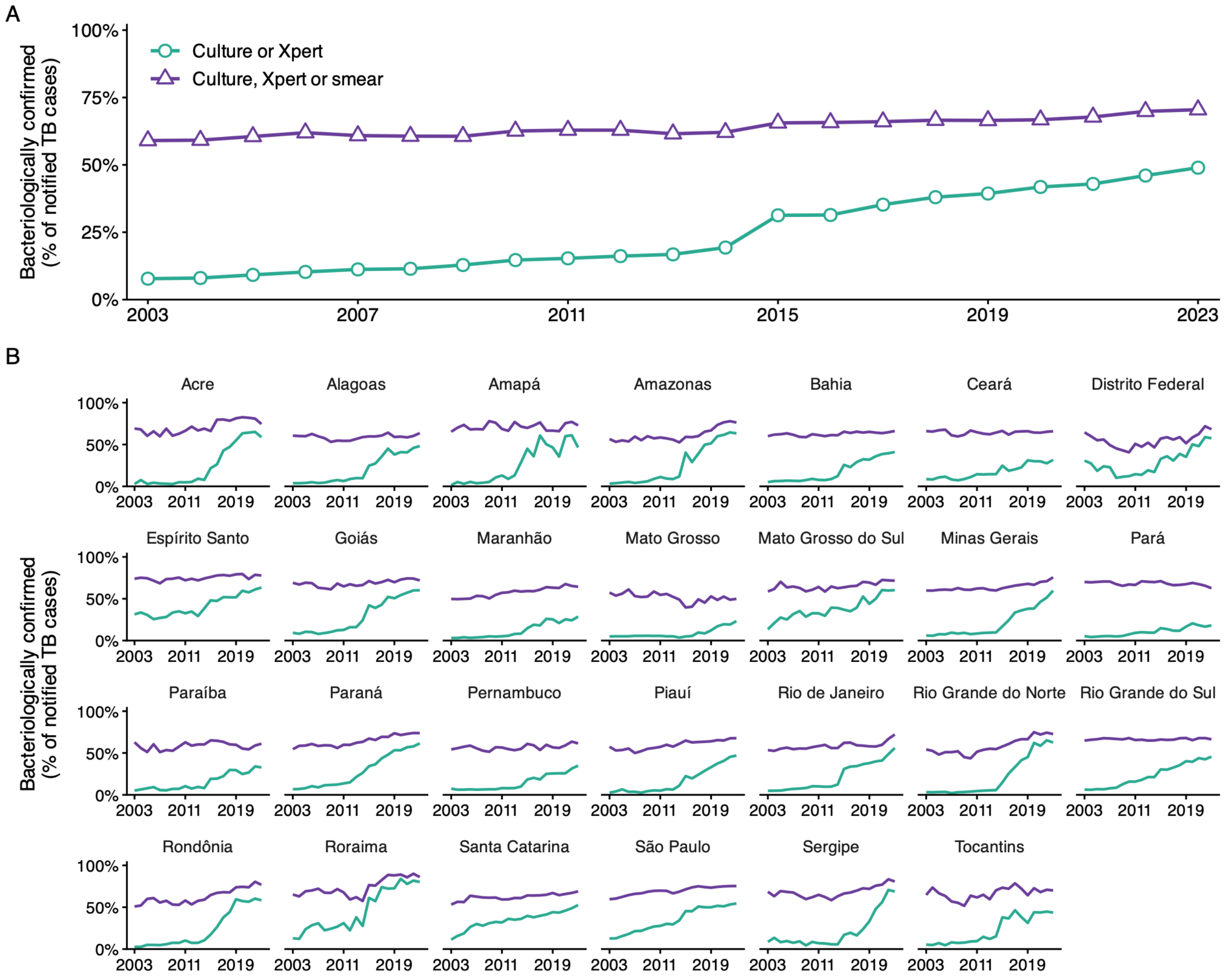


**Supplementary Fig. 30. Annual proportion of bacteriologically confirmed cases among notified TB cases across the analytic sample of Brazilian municipalities, Jan 1, 2003 – Dec 1, 2023, under two definitions of bacteriological confirmation.** Under the culture or Xpert definition (teal), a notified case is classified as bacteriologically confirmed if it is positive on any of: sputum culture (CULTURA_ES = 1), culture of other (extra-pulmonary) material (CULTURA_OU = 1), or the rapid molecular Xpert MTB/RIF assay (TEST_MOLEC = detectable, rifampicin-sensitive or -resistant). The culture, Xpert or smear definition (purple) additionally counts a positive smear on the first sputum sample (BACILOSC_E = 1) or on other material (BACILOSC_O = 1). Missing values on a confirmation field are counted as not confirmed. Aggregation is case-weighted (sum of confirmed cases divided by sum of notified cases within each year and scope). (A) Brazil overall: national case-weighted proportion by calendar year. (B) Per state results, showing the case-weighted proportion across municipalities in that state.

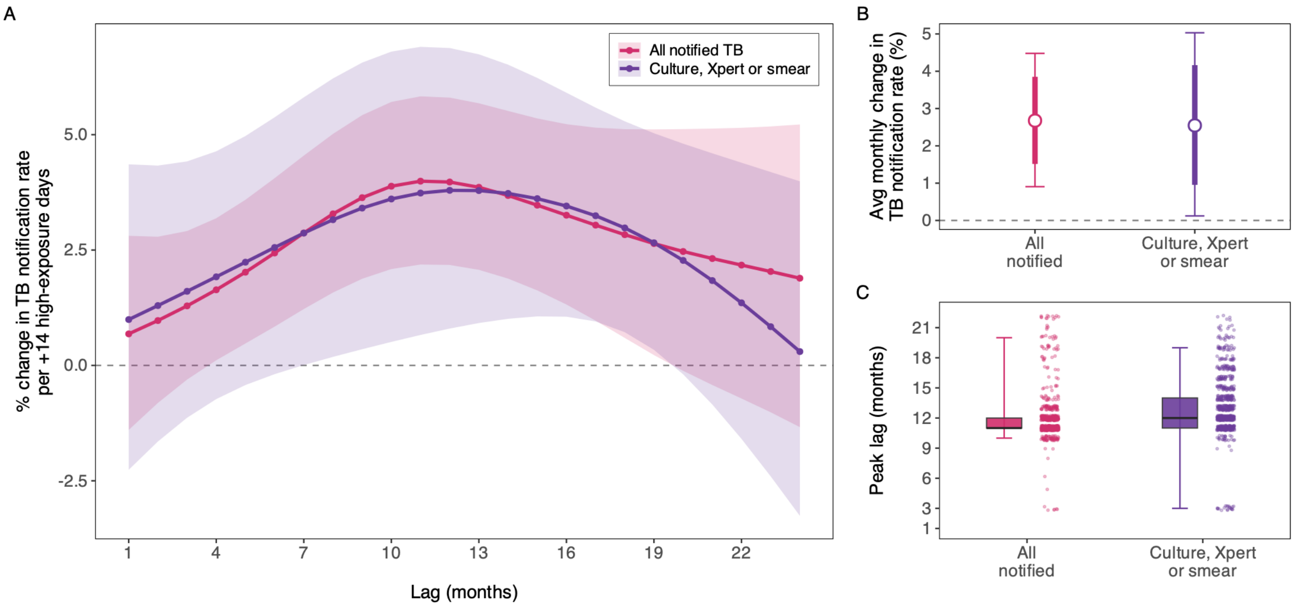


**Supplementary Fig. 31. Bacteriologically confirmed robustness analysis of the relationship between wildfire-related $PM_{2.5}$ and TB notification rates, using the culture, Xpert or smear definition.** Penalized-spline distributed-lag model ($PM_{2.5}$ > 25 µg/m³ exceedance days, GFED4.1s wildfire-related $PM_{2.5}$), Brazil, Jan 1, 2005 – Dec 1, 2023. The main analysis uses all notified TB cases (the primary outcome); the sensitivity analysis re-fits the identical model on bacteriologically confirmed cases only, i.e., those positive on smear microscopy (first sputum sample or other material), on sputum or other-material culture, or on the rapid molecular Xpert MTB/RIF assay. The two analyses share the same fixed effects, offset, lag basis, exposure definition, and Conley standard errors; only the outcome count differs. (A) Lag-response curves for the main analysis (all notified TB) and the bacteriologically confirmed sensitivity analysis. Lines are the mean percent change in the TB notification rate per 14 additional high-exposure days (days above 25 µg/m³) at each lag from 1 to 24 months; shaded bands are 95% Conley confidence intervals. The dashed line indicates no effect. (B) Average monthly effect for each model: the geometric mean of the per-month rate ratios across the 1–24-month lag window, expressed as percent change in the TB notification rate per 14 additional high-exposure days. The white point is the central estimate; thick bars show the 80% interval and thin bars the 95% interval, both from Conley spatial HAC standard errors. The dashed line indicates no effect. (C) Posterior distribution of the lag at which the effect peaks, for each model. Peak lags are obtained by Monte Carlo sampling (N = 10,000) of the model coefficients, reconstructing the lag-response curve for each draw, and taking the argmax. Boxplots show the median and interquartile range with whiskers at the 2.5th and 97.5th percentiles; points are a subsample of retained draws.

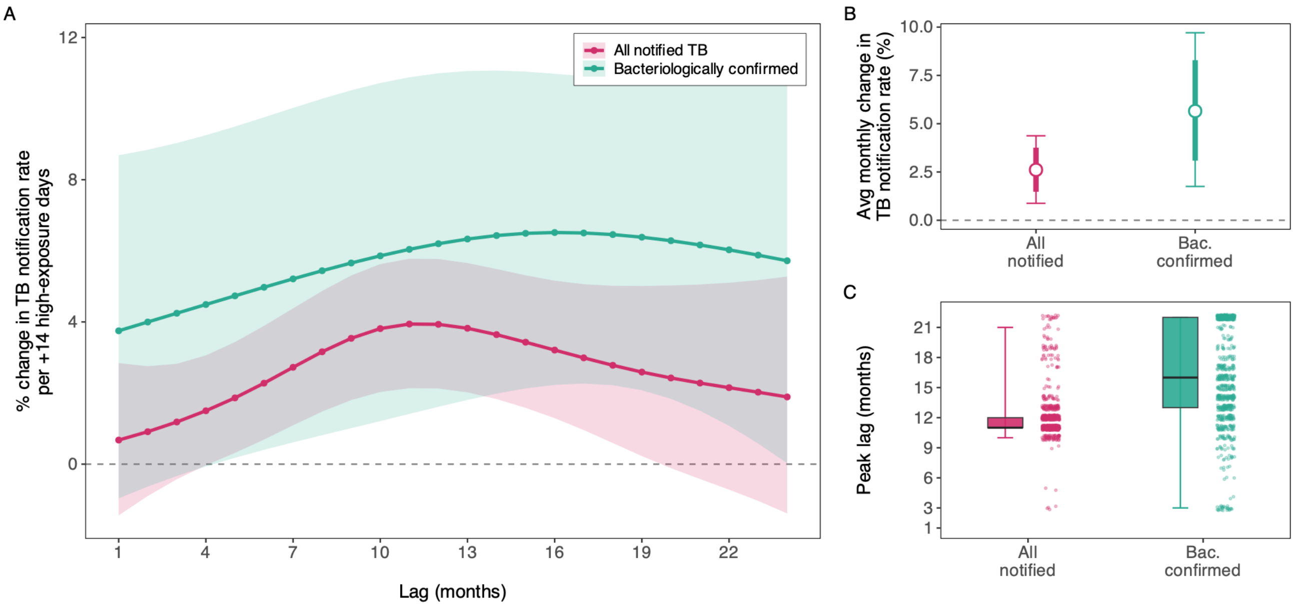


**Supplementary Fig. 32. Bacteriologically confirmed robustness analysis of the relationship between wildfire-related $PM_{2.5}$ and TB notification rates, using the culture or Xpert definition.** Penalized-spline distributed-lag model ($PM_{2.5}$ > 25 µg/m³ exceedance days, GFED4.1s wildfire-related $PM_{2.5}$), Brazil, Jan 1, 2005 – Dec 1, 2023. The main analysis uses all notified TB cases (the primary outcome); the sensitivity analysis re-fits the identical model on bacteriologically confirmed cases only, i.e., those positive on sputum or other-material culture, or on the rapid molecular Xpert MTB/RIF assay. The two analyses share the same fixed effects, offset, lag basis, exposure definition, and Conley standard errors; only the outcome count differs. (A) Lag-response curves for the main analysis (all notified TB) and the bacteriologically confirmed sensitivity analysis. Lines are the mean percent change in the TB notification rate per 14 additional high-exposure days (days above 25 µg/m³) at each lag from 1 to 24 months; shaded bands are 95% Conley confidence intervals. The dashed line indicates no effect. (B) Average monthly effect for each model: the geometric mean of the per-month rate ratios across the 1–24-month lag window, expressed as percent change in the TB notification rate per 14 additional high-exposure days. The white point is the central estimate; thick bars show the 80% interval and thin bars the 95% interval, both from Conley spatial HAC standard errors. The dashed line indicates no effect. (C) Posterior distribution of the lag at which the effect peaks, for each model. Peak lags are obtained by Monte Carlo sampling (N = 10,000) of the model coefficients, reconstructing the lag-response curve for each draw, and taking the argmax. Boxplots show the median and interquartile range with whiskers at the 2.5th and 97.5th percentiles; points are a subsample of retained draws.

#### S3.9 Sensitivity to adjustment for GeneXpert diagnostic

Because rapid molecular testing (GeneXpert / Xpert MTB/RIF) was rolled out from 2014 and raised TB case detection over time, the primary model adjusts for the municipality-year proportion of notifications with a rapid molecular result. We re-fitted the identical penalized distributed-lag model (lags 1-24 months; municipality, state-by-year and state-by-month fixed effects; Conley SEs at 300 km) with this covariate *excluded* from the model.

**Results.** The lag-response was essentially unchanged (Supplementary Fig. 33). The average monthly effect (+2.7%; 95% CI +0.9 to +4.5 vs +2.8%; 95% CI: +1.1 to +4.6) and the median peak lag were likewise concordant. The association is therefore not affected by controlling for the GeneXpert diagnostic scale-up.

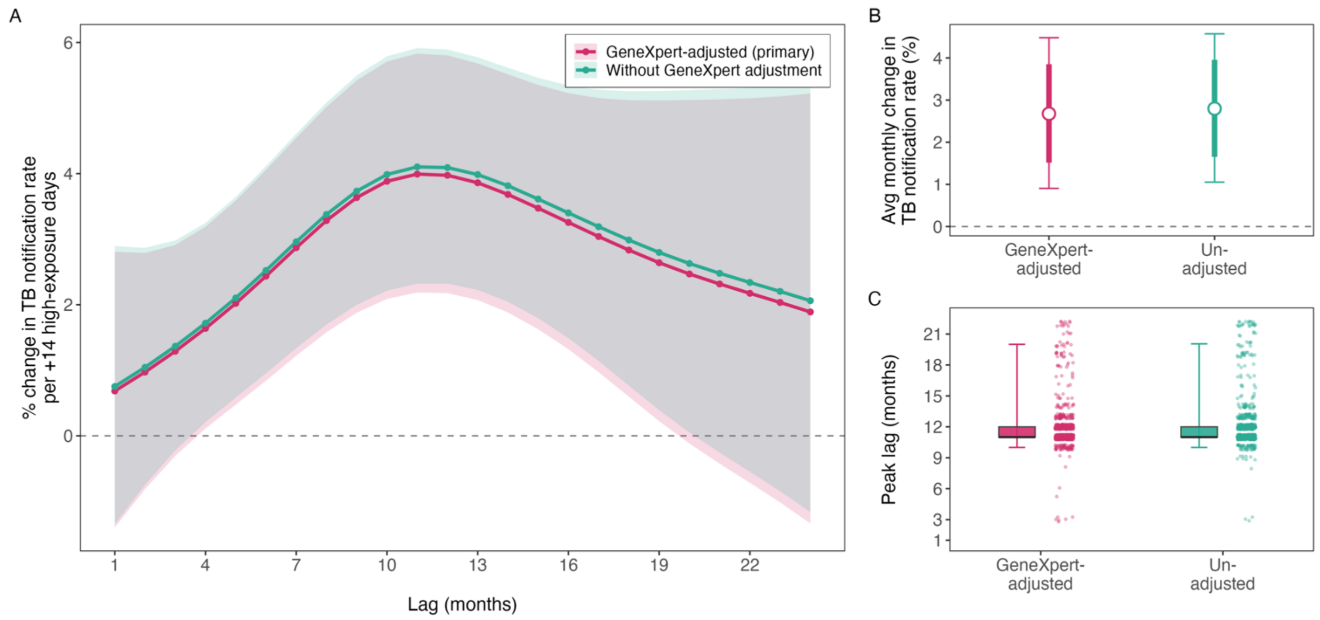


**Supplementary Fig. 33. Robustness of the wildfire-related $PM_{2.5}$ to tuberculosis lag-response to adjustment for the GeneXpert (rapid molecular test) diagnostic scale-up.** The primary analysis adjusts for the municipality-year proportion of notified TB cases with a rapid molecular (GeneXpert / Xpert MTB/RIF) result, a time-varying control for the diagnostic scale-up rolled out across the Brazilian public TB network from 2014; the sensitivity analysis re-fits the identical model with that covariate dropped. (A) Lag-response curves for the primary (GeneXpert-adjusted) analysis and the unadjusted sensitivity analysis. Lines are the mean percent change in the TB notification rate per +14 additional high-exposure days (days above 25 $\mu g/m^3$) at each lag from 1 to 24 months; shaded bands are 95% Conley confidence intervals. The dashed line indicates no effect. (B) Average monthly effect for each model: the geometric mean of the per-month rate ratios across the 1-24 month lag window, expressed as percent change in the TB notification rate per 14 additional high-exposure days. The white point is the central estimate; thick bars show the 80% interval and thin bars the 95% interval, both from Conley spatial HAC standard errors. The dashed line indicates no effect. (C) Posterior distribution of the lag at which the effect peaks, for each model. Peak lags are obtained by Monte Carlo sampling (N = 10,000) of the model coefficients, reconstructing the lag-response curve for each draw, and taking the argmax. Boxplots show the median and interquartile range with whiskers at the 2.5th and 97.5th percentiles; points are a subsample of retained draws.

#### S3.10 Testing for residual seasonality by extending the lag window

Wildfire-related $PM_{2.5}$ shows strong patterns of annual seasonality (wildfire-related smoke peaks in the dry season). Although state-by-month fixed effects absorb the average within-state seasonal pattern, a residual concern is that the 11-month peak in the lag-response could partly reflect exposure and notifications co-peaking in the same calendar months rather than a genuine lagged biological effect. Such an artefact has a signature: it would recur at the annual harmonic, producing a second mode in the lag-response near 24 months, whereas a true latency should decay to the null after its peak and stay there. To look for this signature we extended the penalized distributed-lag model from 1-24 to 1-36 months, leaving the model otherwise identical and fitted over the same municipalities as the primary analysis. Because the last four lags of a 36-month lag basis sit at its boundary, where the fit is least stable, the extended lag-response is shown and summarized over lags 1-32.

**Results.** No annual recurrence appeared (Supplementary Fig. 34). The lag-response peaked at the same lag under both windows (posterior median 11 months, 95% interval 10-20, over 1-24 and 11 months, 8-18, over 1-32) and then decayed monotonically to the null, with no second hump near 24 months. The average monthly effect per 14 additional high-exposure days was 2.7% (95% CI 0.9 to 4.5) over the 1-24 month window and 1.6% (95% CI -0.7 to 4.0) over 1-32, and over the extended tail (lags 25-32) it was -0.4% (95% CI -3.4 to 2.7), an interval that includes the null. The absence of a recurring, roughly annual hump indicates that the 11-month peak reflects a genuine lagged effect on the latent-reactivation timescale rather than residual seasonality.

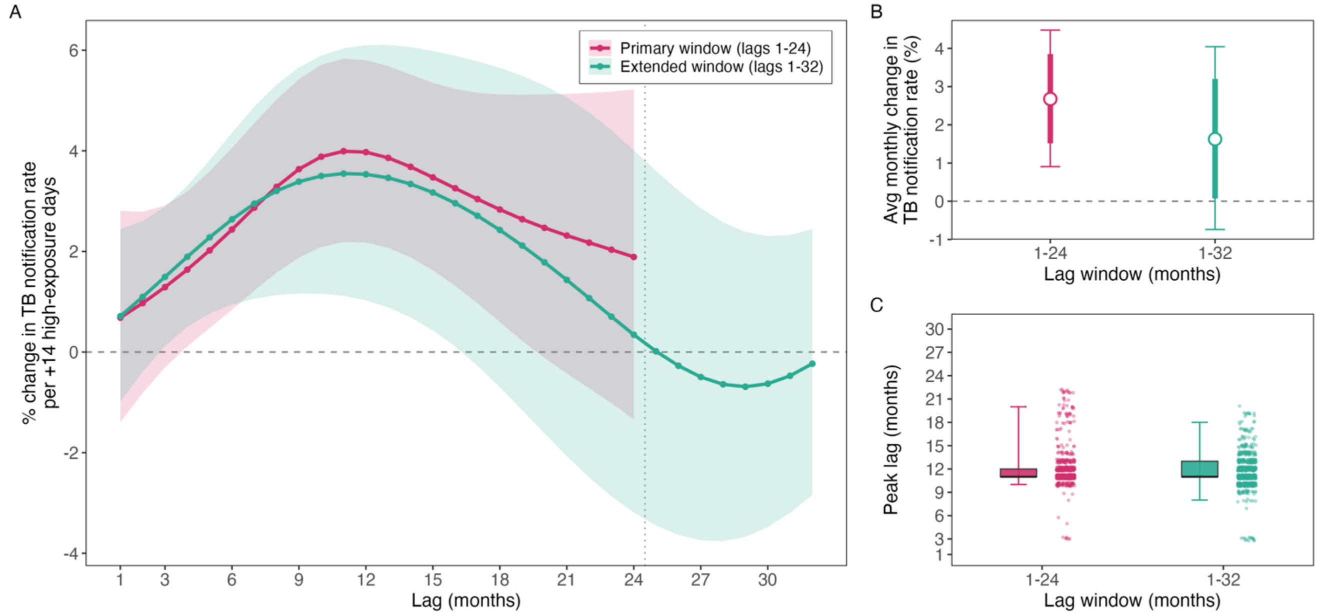


**Supplementary Fig. 34. Sensitivity of the wildfire-related $PM_{2.5}$ to tuberculosis lag-response to the length of the distributed-lag window.** Penalized-spline distributed-lag models ($PM_{2.5}$ > 25 µg/m³ exceedance days, GFED4.1s), Brazil, Jan 1, 2005 – Dec 1, 2023 (from Jan 1, 2006 for the 36-month window), fitted across municipalities with municipality, state × year, state × month, and state × pandemic-phase fixed effects, a log-population offset, and Conley spatial HAC standard errors (cutoff 300 km). The primary 1–24-month window (pink) is compared with the same model refit over an extended 1–36-month window (teal), shown and summarized over lags 1–32; the two share an identical penalized P-spline lag basis (k = 8) and differ only in window length. (A) Lag-response curves for the two windows. Lines are the mean percent change in TB notification rates per 14 additional high-exposure days at each monthly lag; shaded bands are 95% Conley confidence intervals. The dashed horizontal line indicates no effect; the dotted vertical line marks the 24-month boundary of the primary window. (B) Average monthly effect for each window: the geometric mean of the per-month rate ratios across the lag window, expressed as percent change in TB notification rates per 14 additional high-exposure days. Thick bars show the 80% interval and thin bars the 95% interval, both from Conley spatial HAC standard errors. The dashed line indicates no effect. (C) Posterior distribution of the lag at which the effect peaks for each window. Peak lags are obtained by Monte Carlo sampling (N = 10,000) of the model coefficients, reconstructing the lag-response curve for each draw, and taking the argmax. Boxplots show the median and interquartile range with whiskers at the 2.5th and 97.5th percentiles; points are a subsample of retained draws.

#### S3.11 Testing for residual seasonality with a de-seasonalized exposure variable

Wildfire-related $PM_{2.5}$ follows a strong annual cycle, with smoke concentrated in the dry season (Supplementary Fig. **9**B). State-by-month fixed effects absorb the average seasonal pattern within each state, but municipalities within a state may differ in the timing and intensity of both their smoke season and their TB notification seasonality. If such municipality-specific seasonal patterns were shared by exposure and outcome, they would confound the relationship between exposure and TB notifications.

**Approach.** We constructed a municipality-specific de-seasonalised exposure variable: For each municipality and calendar month, we computed the mean number of high-exposure days ($PM_{2.5}$ > 25 µg/m³) over Jan 1, 2003 – Dec, 1 2023, and replaced each lagged exposure value by its deviation from the mean for its own calendar month. The model was otherwise identical to the one used in the primary analysis. Under residual seasonality, the de-seasonalised lag-response would be expected to flatten, or its peak to shift.

**Results.** The de-seasonalized exposure variable retained 53% of the variance of the original high-exposure-day series (SD 1.21 vs 1.67 days). The re-estimated association was unchanged after removing municipality-specific seasonality (Supplementary Fig. **35**). The average monthly effect was +2.7% (95% CI 0.8–4.6) per 14 additional high-exposure days, compared with +2.7% (0.9–4.5) in the primary analysis. The lag-response retained its unimodal shape and peaked at 11 months in both analyses (posterior median 11 months; IQR 11–11 after de-seasonalisation vs IQR 11–12 in the primary analysis), with confidence intervals excluding the null from lags 7 to 18. If anything, the peak more pronounced (+4.7% vs +4.0% at lag 11). These results indicate that the delayed peak reflects year-to-year variation in smoke exposure rather than a seasonal pattern shared by wildfire smoke and TB notifications.

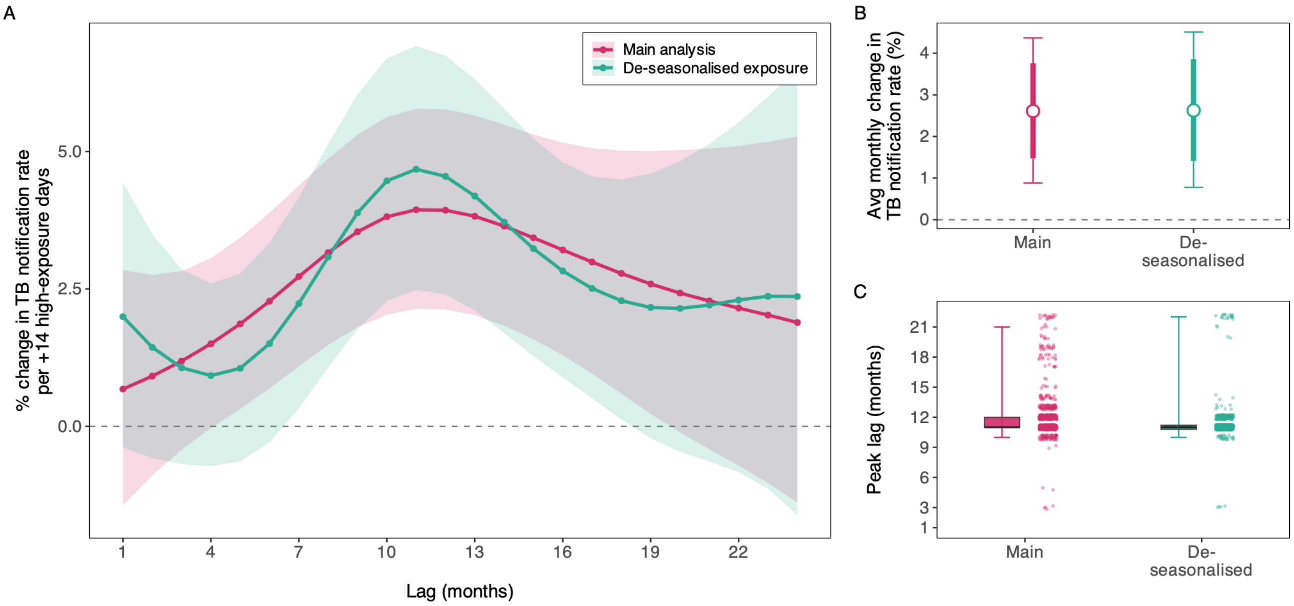


**Supplementary Fig. 35. Robustness of the lag-response relationship between wildfire-related $PM_{2.5}$ and TB notification rates to removal of municipality-specific seasonality, Brazil, Jan 1, 2005 – Dec 1, 2023.** Penalized-spline distributed-lag model (PM2.5 > 25 µg/m³ high-exposure days, GFED4.1s; k = 8, lags 1–24 months) with municipality, state × year, state × month and state × pandemic-phase fixed effects, a log-population offset, and Conley spatial HAC standard errors (300 km cutoff). This sensitivity analysis replaces the monthly count of high-exposure days with its seasonal anomaly (monthly count minus the municipality's 2003–2023 mean for that calendar month). The two analyses differ only in the exposure. (A) Lag-response curves: mean percent change in the TB notification rate per 14 additional high-exposure days at each lag. Shaded bands are 95% confidence intervals; the dashed line indicates no effect. (B) Average monthly effect: geometric mean of the per-month rate ratios across lags 1–24, with 80% (thick) and 95% (thin) intervals. (C) Posterior distribution of the peak lag, from 10,000 draws of the basis coefficients (Conley covariance). Boxplots show the median and interquartile range with whiskers at the 2.5th and 97.5th percentiles.

#### S3.12 Conley distance-cutoff sensitivity

As expected, the point estimates are unchanged by the cutoff. However, the precision of the estimates change. At the primary 300 km cutoff the association is statistically significant: the average monthly effect is +2.7% (95% CI +0.9 to +4.5), excluding the null. Widening the Conley cutoff to 500 km inflates the standard errors - the variance estimator now accumulates additional, predominantly positive, long-range spatial covariance - and the interval widens accordingly: the average monthly effect becomes +2.7% (95% CI -0.5 to +6.0), so that the 95% interval at 500 km just includes zero (Supplementary Fig. 36, A-B).

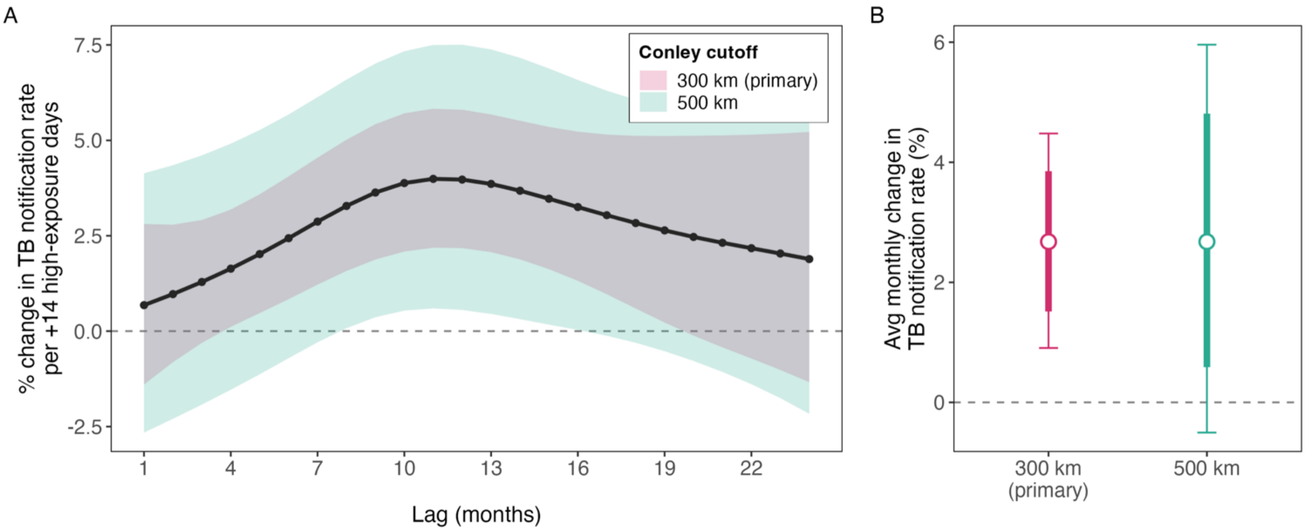


**Supplementary Fig. 36. Conley spatial heteroskedasticity- and autocorrelation-consistent (HAC) sensitivity analysis to the distance cutoff (300 km primary vs 500 km) for the wildfire-related $PM_{2.5}$ to tuberculosis lag-response.** Penalized-spline distributed-lag model ($PM_{2.5}$ > 25 µg/m³ exceedance days, GFED4.1s) of monthly TB notification rates across Brazilian municipalities, Jan 1, 2005 – Dec 1, 2023, with municipality, state × year, state × month, and state-by-pandemic-phase fixed effects and a log-population offset. The Conley cutoff is the distance over which spatially correlated scores are accumulated in the variance estimator; it enters the standard errors only, so the model coefficients are identical across cutoffs and only the confidence intervals differ. (A) Lag-response curve (single shared point estimate, dark line and points) with the 95% Conley confidence band evaluated at the 300 km primary cutoff and at the 500 km cutoff overlaid. The y-axis is the mean percent change in TB notification rates per 14 additional high-exposure days at each lag. The dashed line indicates no effect. (B) Average monthly effect at each cutoff: the geometric mean of the per-month rate ratios across the 24-month lag window, expressed as percent change in TB notification rates per 14 additional high-exposure days. The point estimate is identical at the two cutoffs; thick bars show the 80% interval and thin bars the 95% interval. The dashed line indicates no effect.

#### S3.13 Negative control exposure

**Approach.** To assess whether the estimated wildfire-related $PM_{2.5}$–tuberculosis association could reflect spatially structured confounding or shared seasonality rather than local smoke exposure, we used a *distant-donor negative-control exposure*. Each index municipality was assigned a donor municipality drawn uniformly at random from all municipalities at least 1,500 km away whose exposure series had at least 90% monthly coverage, and the donor's monthly count of wildfire-related $PM_{2.5}$ exceedance days (>25 µg/m³) was carried onto the index municipality at matching calendar months. Because 1,500 km greatly exceeds the transport distance of wildfire plumes in tropical South America, the donor series shares no physical smoke-transport pathway with the index municipality while retaining broadly similar seasonal and climatic structure; under the null of no residual spatial confounding, the donor (distant) exposure should be unrelated to local tuberculosis notifications. We implemented this falsification in two complementary ways.

<u>Distributed-lag picture.</u> We refitted the primary penalized distributed-lag model with the distant-donor exceedance history substituted for the municipality's own (local) exceedance history, entered as a penalized P-spline over lags 1–24 months, retaining the municipality, state×year, state×month, state-by-pandemic-phase fixed effects, the temperature and precipitation lag structure, and the log-population offset, with Conley spatial standard errors (300 km cutoff). A single donor assignment (fixed random seed) was used because each penalized fit is computationally intensive. We compare the lag-response curve and average monthly effect of the

distant placebo exposure with those of the primary analysis: a positive local curve peaking at 10 to 12 months alongside a null placebo curve is consistent with a local interpretation.

Permutation null. To calibrate the placebo against chance, we repeated the random donor assignment 500 times and, for each, refitted a collapsed-lag exposure model (high-exposure days summarized over a 12-month window) by Poisson fixed-effects regression with the donor exposure substituted for the real one, retaining the same fixed-effect structure, climate adjustment, and log-population offset. This produced the empirical (permutation) distribution of the placebo coefficient across the 500 random donor assignments. The observed real-exposure coefficient, estimated with Conley spatial standard errors (300 km cutoff), was referenced against this distribution; we report a two-sided permutation p-value (the proportion of placebo draws with a coefficient at least as large in magnitude as the real effect) and the corresponding one-sided value. The collapsed-lag specification was used for this resampling step because its low per-fit cost makes 500 refits tractable, whereas the penalized fit is run once. All effects are reported per 14 additional high-exposure days, consistent with the main analysis.

**Result.** In the penalized model, the average monthly effect per 14 additional high-exposure days was +2.7% (95% CI 0.9 to 4.5) for the genuine local exposure and −0.7% (95% CI −1.6 to 0.1) for the distant placebo, and the placebo lag-response showed no peak at the lag where the local effect peaked (11 months). In the collapsed-lag permutation, the genuine local exposure was associated with a +2.2% change in the TB notification rate per additional 14 high-exposure days (95% CI: 0.3–4.0%), whereas the 500 placebo coefficients were centred on zero (mean +0.21%; 2.5th–97.5th percentile −0.81% to +1.25%; largest absolute placebo effect 1.67%), giving a two-sided permutation $p < 0.002$. These results suggest that the primary association depends on the municipality's own smoke exposure rather than on Brazil-wide seasonal or temporal patterns (Supplementary Fig. 37).

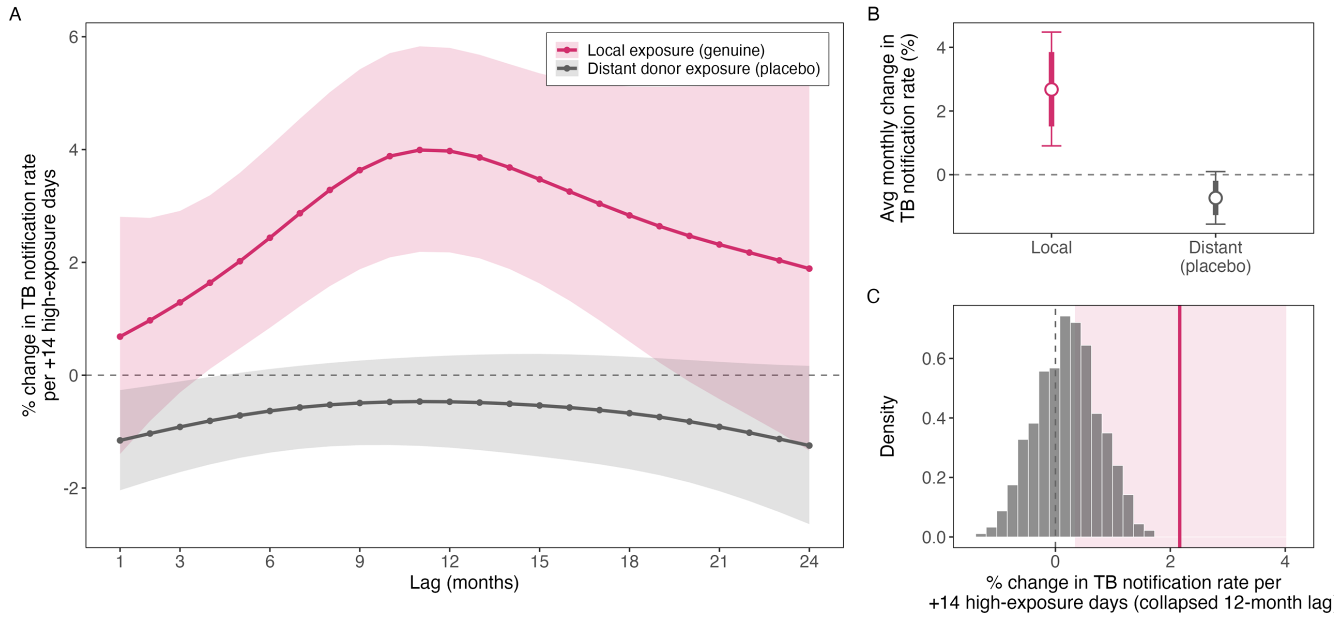


**Supplementary Fig. 37. Distant-exposure negative-control (placebo) test for the relationship between wildfire-related $PM_{2.5}$ and TB notification rates in Brazil, Jan 1, 2005 – Dec 1, 2023. Each municipality's exposure (days per month with wildfire-related $PM_{2.5}$ > 25 µg/m³, GFED4.1s) is replaced by that of a randomly drawn donor municipality at least 1,500 km away, so the placebo exposure follows the national fire-season calendar but carries no local smoke; the genuine local exposure is not retained in the placebo models. (A) Lag-response curves from the**

**penalized-spline distributed-lag model (lags 1–24 months, k = 8; municipality, state-by-year, and state-by-month fixed effects; log-population offset; Conley spatial HAC standard errors, 300 km cutoff) fitted with the genuine local exposure (pink) and, for one random donor pairing, with the distant placebo exposure (grey). Lines are the percent change in TB notification rates per 14 additional high-exposure days at each lag; shaded bands are 95% Conley confidence intervals; the dashed line indicates no effect. (B) Average monthly effect over lags 1–24 for the two fits in (A): the geometric mean of the per-month rate ratios, expressed as percent change per 14 additional high-exposure days. Thick bars show the 80% interval and thin bars the 95% interval. (C) Permutation null for the collapsed-lag model. The Poisson fixed-effects model with the same fixed effects and offset and a single 12-month collapsed exposure term was refitted over 500 random donor pairings; the histogram shows the placebo coefficients (percent change per 14 additional high-exposure days). The vertical pink line is the coefficient from the genuine local fit and the shaded band its Conley 95% confidence interval; the dashed line indicates no effect. The permutation p-value is the fraction of pairings whose placebo coefficient is at least as large in absolute value as the genuine one.**

#### S3.14 Negative control outcomes

A key concern is that the association between wildfire-related $PM_{2.5}$ and TB notifications could still reflect residual time-varying confounding. Seasonal weather patterns, changes in healthcare access or surveillance intensity, or other municipality-level environmental exposures could vary with wildfire-related smoke and also affect disease notifications. If so, an apparent lag-response could arise even without a true biological effect of wildfire-related $PM_{2.5}$ on TB progression. We therefore conducted negative-control outcome analyses to assess whether similar associations were present for outcomes not expected to be affected by wildfire smoke through the proposed TB-specific mechanism. A valid negative-control outcome should share some of the same potential sources of confounding as the primary outcome but should not be causally affected by the exposure through the hypothesized pathway.[34]

##### *S3.14.1 Leprosy*

Leprosy is useful for this purpose because it is captured in the same national surveillance system as TB, but differs in ways that make a short-lag effect of wildfire-related $PM_{2.5}$ unlikely. Its natural latency period is much longer than TB, often spanning several years (5-20 years from infection to clinically apparent disease), and its diagnosis is generally distinct from acute pulmonary TB care. Therefore, an association between wildfire-related $PM_{2.5}$ and leprosy notifications at the same lag where the TB effect peaks would raise concern about shared time-varying confounding.

**Approach.** We re-fit the primary model using monthly leprosy notifications instead of TB notifications as the outcome. Leprosy counts were obtained from the SINAN-HANS module, aggregated by municipality of residence and month (Supplementary Fig. 11). The event date was based on date of diagnosis, with date of notification used when diagnosis date was unavailable, matching the approach used for TB. Municipality-months with no leprosy notifications were treated as zero counts rather than missing values. The model specification was otherwise kept the same as in the primary analysis. The only outcome-specific change was that we removed the TB-specific GeneXpert diagnostic-coverage covariate, which has no equivalent for leprosy. The pandemic-phase adjustment was retained because leprosy case detection was also disrupted during the COVID-19 period.

**Results.** The analysis included 689,055 leprosy notifications across 3,434 municipalities and 243,777 municipality-months from January 2003 to December 2023. At least one leprosy notification was recorded in approximately 38% of municipality-months. The leprosy lag-response was flat and centered near the null throughout the 24-month lag window (Supplementary Fig. 38). The curve did not show a unimodal pattern or a peak near the TB lag-response. The average monthly effect was -0.72% (95% CI -7.74 to 6.83) per additional 14 high-exposure days. However, because leprosy notifications were much less frequent than TB notifications, the wider confidence intervals mean that this negative-control result should be interpreted as supportive rather than definitive evidence.

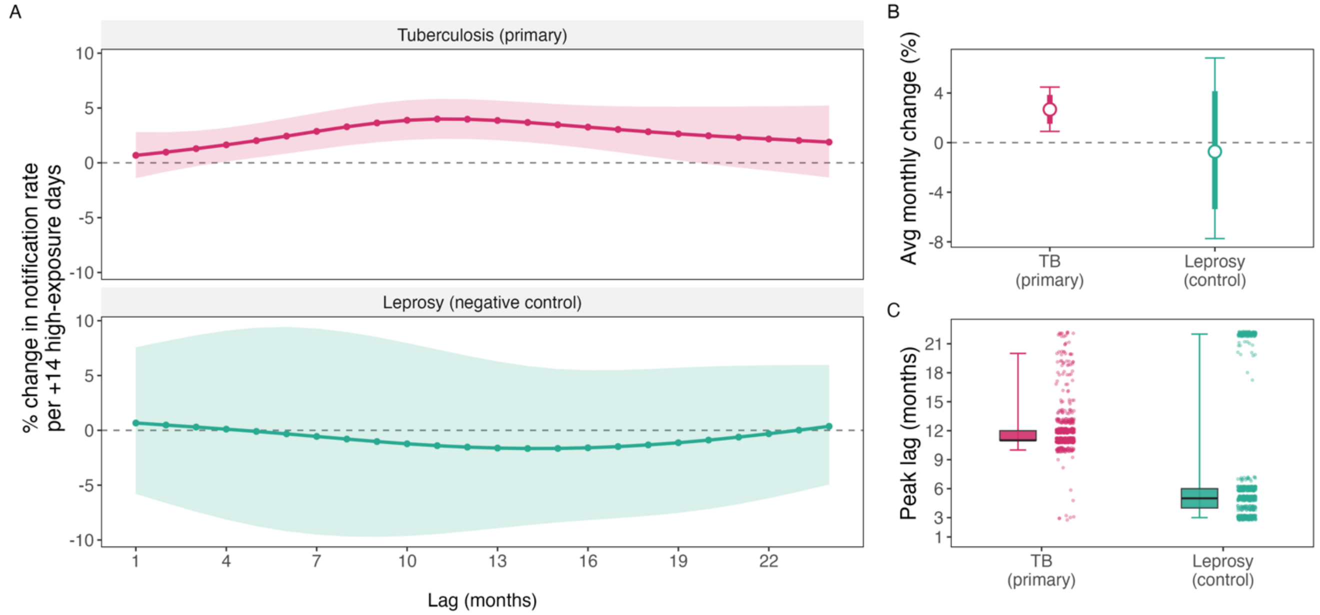


**Supplementary Fig. 38. Leprosy as a negative-control outcome for the wildfire-related $PM_{2.5}$ to tuberculosis lag-response.** Penalized-spline distributed-lag model ($PM_{2.5}$ > 25 μg/m³ exceedance days, GFED4.1s wildfire-related $PM_{2.5}$), Brazil, Jan 1, 2005 – Dec 1, 2023, with municipality, state × year, and state × month fixed effects, a log-population offset, and Conley spatial HAC standard errors (cutoff 300 km). The primary analysis uses all notified TB cases; the negative-control analysis re-fits the identical model on municipality-monthly counts of leprosy (hanseniase) notifications from SINAN-HANS. The two analyses share the same fixed effects, offset, lag basis, exposure definition, and Conley standard errors; only the outcome count differs (the TB-specific diagnostic-coverage covariate is dropped for leprosy). (A) Lag-response curves for tuberculosis (primary outcome, upper panel) and leprosy (negative control, lower panel). Each line is the mean percent change in that disease's own notification rate per 14 additional high-exposure days (days above 25 μg/m³) at each lag from 1 to 24 months; shaded bands are 95% Conley confidence intervals. The dashed line indicates no effect. (B) Average monthly effect for each outcome: the geometric mean of the per-month rate ratios across the 1-24 month lag window, expressed as percent change in the notification rate per +14 high-exposure days. The white point is the central estimate; thick bars show the 80% interval and thin bars the 95% interval, both from Conley spatial HAC standard errors. The dashed line indicates no effect. (C) Posterior distribution of the lag at which the effect peaks, for each outcome. Peak lags are obtained by Monte Carlo sampling (N = 10,000) of the model coefficients, reconstructing the lag-response curve for each draw, and taking the argmax. Boxplots show the median and interquartile range with whiskers at the 2.5th and 97.5th percentiles; points are a subsample of retained draws.

##### *S3.14.2 Appendicitis*

**Approach.** We re-fitted the primary penalized distributed-lag model on monthly counts of acute appendicitis admissions in place of TB notifications. Appendicitis was selected as a negative control outcome because it is an acute surgical condition with no clear biological link to chronic $PM_{2.5}$ exposure or to the cell-mediated immunity that contains mycobacteria, while sharing the same source population, calendar-time structure, and hospital-ascertainment pathway as the TB outcome.

**Results.** Whereas the primary analysis showed a hump-shaped lag-response peaking at 11 months (IQR: 11-12) with an average monthly effect of +2.7% per 14 additional high-exposure days (95% CI 0.9 to 4.5%), the appendicitis analysis was null at every lag (Supplementary Fig. 39): the average monthly effect was -0.3% (95% CI -3.3% to +2.8%), and the posterior distribution of the peak lag was essentially uninformative, with 57% of draws showing no interior maximum. Although no negative control outcome can rule out all forms of residual confounding, the absence of any $PM_{2.5}$ to appendicitis association argues against shared hospital-access or diagnostic changes as the main driver of the TB association.

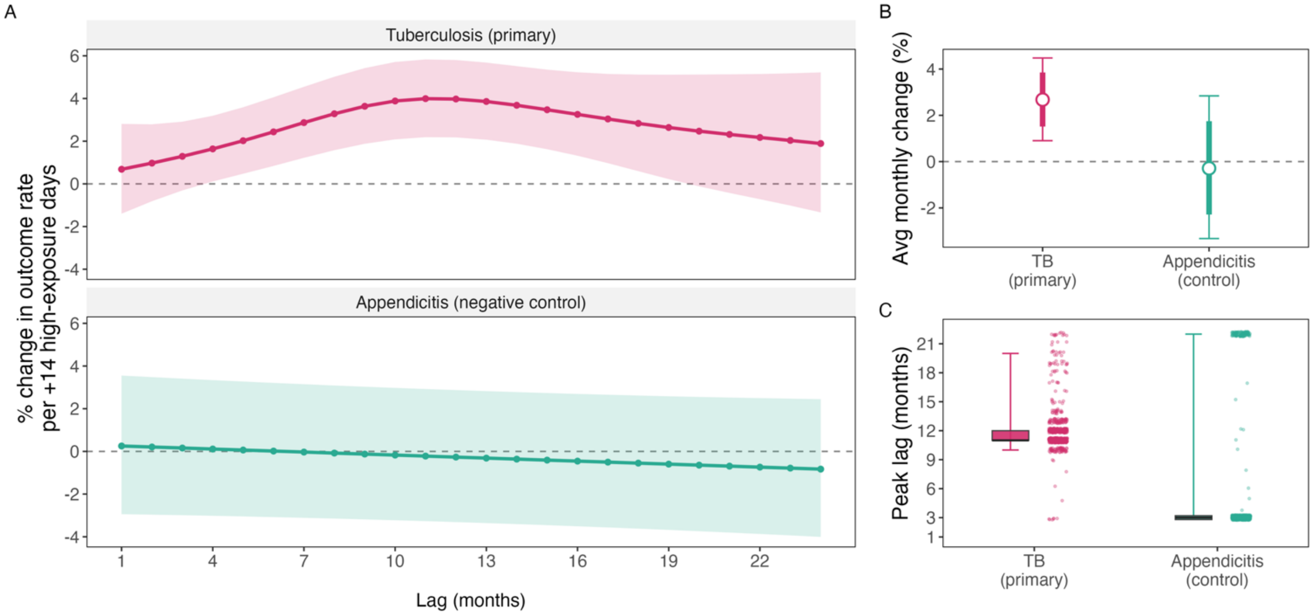


**Supplementary Fig. 39. Appendicitis as a negative-control outcome for the wildfire-related $PM_{2.5}$ to tuberculosis lag-response.** Penalized-spline distributed-lag model ($PM_{2.5}$ > 25 µg/m³ exceedance days, GFED4.1s $PM_{2.5}$), Brazil, with municipality, state × year, and state × month fixed effects, a log-population offset, and Conley spatial HAC standard errors (cutoff 300 km). The primary analysis uses all notified TB cases; the negative-control analysis re-fits the identical model on municipality-monthly counts of acute appendicitis hospital admissions (SIH-RD, ICD-10 K35). The two analyses share the same fixed effects, offset, lag basis, exposure definition, and Conley standard errors; only the outcome count differs (the TB-specific diagnostic-coverage covariate is dropped for appendicitis). (A) Lag-response curves for tuberculosis (primary outcome, upper panel) and appendicitis (negative control, lower panel), shown as stacked facets on a shared y-axis so their magnitudes are directly comparable. Each line is the mean percent change in that outcome's own rate per 14 additional high-exposure days (days above 25 µg/m³) at each lag from 1 to 24 months; shaded bands are 95% Conley confidence intervals. The dashed line indicates no effect. (B) Average monthly effect for each outcome: the geometric mean of the per-month rate ratios across the 1-24 month lag window, expressed as percent change in the outcome's rate per 14 additional high-exposure days. The white point is the central estimate; thick bars show the 80% interval and thin bars the 95% interval, both from Conley spatial HAC standard errors. The dashed line indicates no effect. (C) Posterior distribution of the lag at which the effect peaks, for each outcome. Peak lags are obtained by Monte Carlo sampling (N = 10,000) of the model coefficients, reconstructing the lag-response curve for each draw. Boxplots show the median and interquartile range with whiskers at the 2.5th and 97.5th percentiles; points are a subsample of retained draws.

#### S3.15 Sensitivity analysis of other wildfire-related effects

##### *S3.15.1 Local fire activity vs transported smoke*

Fires can affect health through pathways other than smoke inhalation, including the loss of crops, pasture, income and food supply in the area that burns. If these local consequences of fire explained the association, wildfire-related $PM_{2.5}$ would partly act as a marker of nearby burning rather than of smoke exposure. However, smoke travels well beyond the area that burns: 34% of high-exposure days occurred in months in which no fire was detected in the municipality (see also Supplementary Fig. **9**C). We used this separation between local burning and smoke in two analyses.

**Approach.** First, we re-fitted the primary penalized distributed-lag model with fire activity in the municipality itself added as a covariate. Local fire was measured as monthly burned area from the Global Fire Atlas (Section S1.9), expressed as the log of one plus the percentage of municipality area burned, with each fire assigned to the municipality and month of its ignition. It entered the model as a natural-spline distributed-lag term over lags 1–24 months (3 degrees of freedom), the same form used for temperature and precipitation. Second, we split the exposure according to whether any burned area was recorded in the municipality in the exposure month. High-exposure days in months without local fire, which reflect smoke originating outside the municipality, and high-exposure days in months with local fire entered the same model as two separate penalized lag terms, each with the lag basis of the primary analysis. We compared their average monthly effects using the joint Conley covariance of the two terms. We repeated the split with local fire defined as any burned area in the municipality's IBGE micro-region. All other model components were unchanged.

**Results.** The analysis included 1,263,996 municipality-months across 5,544 municipalities. Local fire was only weakly correlated with the $PM_{2.5}$ exposure (r = 0.20 over lags 1–24 months). Adjustment for local fire did not change the association (Supplementary Fig. 40). The average monthly effect per 14 additional high-exposure days was +2.8% (95% CI 1.0 to 4.6) after adjustment, compared with +2.7% (95% CI 0.9 to 4.5) in the main analysis. The two lag-response curves differed by at most 0.6 percentage points at any lag, and the median peak lag was 11 months in both analyses (95% interval 10–20 in the main analysis and 8–18 after adjustment). Local fire itself was not associated with higher TB notification rates (average monthly effect −0.3% [95% CI −0.5 to −0.1] per unit increase in the log of one plus the percentage of area burned). In the split analysis, the average monthly effect was +1.4% (95% CI −2.0 to 5.0) for high-exposure days without local fire and +2.9% (95% CI 1.2 to 4.6) for high-exposure days with local fire, with no evidence of a difference between the two (ratio of average monthly rate ratios 0.99, 95% CI 0.96 to 1.01; p = 0.30). When local fire was defined at the micro-region level, only 13% of high-exposure days occurred without local fire, and the corresponding estimate was imprecise (−3.5%, 95% CI −12.7 to 6.7, vs +3.0%, 95% CI 1.1 to 4.9, with local fire; p = 0.22). These results indicate that the association is not explained by the non-smoke consequences of fire within the municipality. The estimates for smoke without local fire were imprecise, so the split analysis alone cannot show that smoke from fires elsewhere has the same effect as smoke accompanied by local burning. In addition, fires smaller than one MODIS pixel (about 0.21 km²) are not detected, so some months classified as without local fire may have included small agricultural burns.

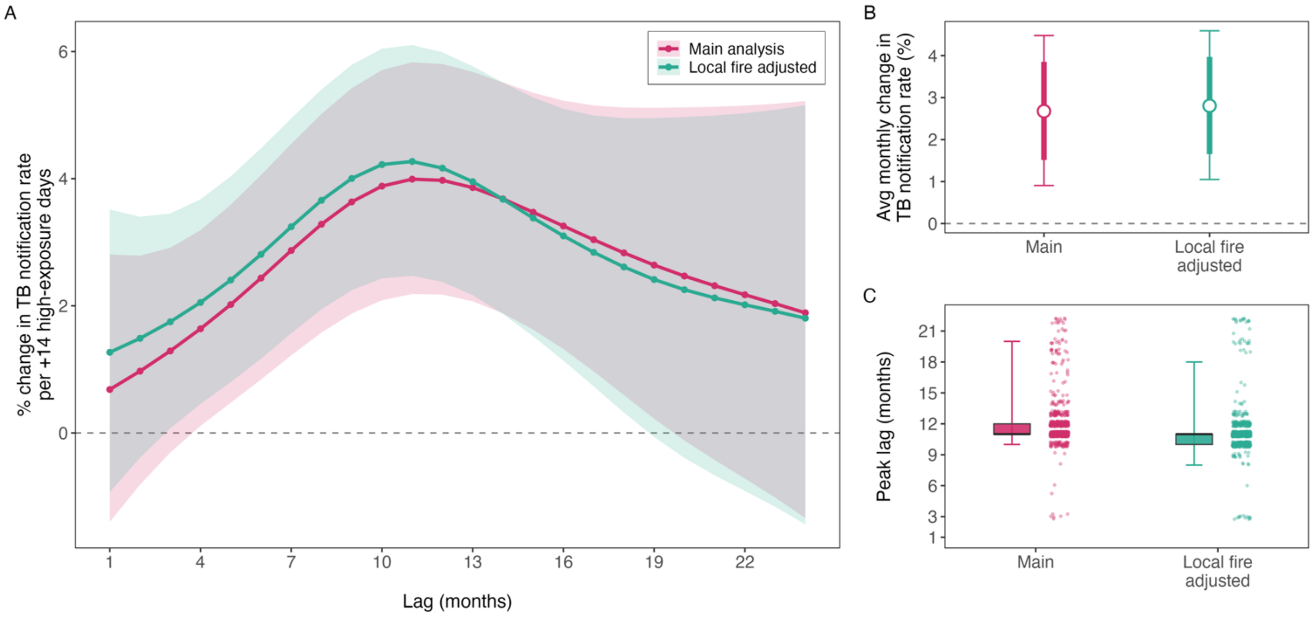


**Supplementary Fig. 40. Robustness of the lag-response relationship between wildfire-related $PM_{2.5}$ and TB notification rates to adjustment for local fire activity.** Penalized-spline distributed-lag model ($PM_{2.5}$ > 25 µg/m³ high-exposure days, GFED4.1s), Brazil, Jan 1, 2005 – Dec 1, 2023, with municipality, state × year, state × month, and state × pandemic-phase fixed effects, a log-population offset, GeneXpert diagnostic coverage, and Conley spatial HAC standard errors (300 km cutoff). The main analysis adjusts for temperature and precipitation, each as a 1–24-month natural-spline distributed-lag term; the sensitivity analysis additionally adjusts for fire activity within the municipality, measured as monthly burned area (Global Fire Atlas; log of one plus the percentage of municipality area burned) and entered in the same form. (A) Lag-response curves for the main analysis and the local-fire-adjusted sensitivity analysis. Lines are the mean percent change in the TB notification rate per 14 additional high-exposure days at each lag from 1 to 24 months; shaded bands are 95% Conley confidence intervals. The dashed line indicates no effect. (B) Average monthly effect for each model: the geometric mean of the per-month rate ratios across the 1–24-month lag window, expressed as percent change in the TB notification rate per 14 additional high-exposure days. The white point is the central estimate; thick bars show the 80% interval and thin bars the 95% interval, both from Conley spatial HAC standard errors. The dashed line indicates no effect. (C) Posterior distribution of the peak lag for each model, obtained by Monte Carlo sampling (N = 10,000) of the model coefficients, reconstructing the lag-response curve for each draw, and taking the argmax. Draws without an interior peak (non-positive mean over the window) were dropped. Boxplots show the median and interquartile range, with whiskers at the 2.5th and 97.5th percentiles; points are a subsample of retained draws.

##### *S3.15.2 Malnutrition hospitalisations as an alternative-pathway outcome*

Wildfires can also lead to damage to crops, pasture and livestock, interruptions of agricultural work and raise of local food prices. These effects could increase undernutrition, which is an established risk factor for progression from latent infection to active TB.[35] This kind of disruption could produce a lagged association between wildfire-related $PM_{2.5}$ and TB that does not reflect smoke inhalation. We examined whether wildfire-related $PM_{2.5}$ was associated with hospital admissions for malnutrition. This analysis tests one specific alternative pathway (rather than residual confounding in general). An association with a lag structure similar to that of TB would suggest that part of the TB signal operates through nutrition. The absence of an association would argue against this pathway.

**Approach.** We re-fitted the primary penalized distributed-lag model using monthly counts of hospital admissions with a principal diagnosis of protein-energy malnutrition (ICD-10 E40–E46; Section S1.13)[17] as the outcome. The model was fitted to the same municipality-months and covariates as the primary analysis.

**Results.** The analysis included 801,541 malnutrition admissions across 5,544 municipalities and 1,263,996 municipality-months. The malnutrition lag-response remained close to the null throughout the 24-month lag window, and the confidence intervals included zero at every lag (Supplementary Fig. **41**, panel A). Point estimates were slightly below zero from lag 3 onwards, including the lags at which the TB effect peaked. The average monthly effect was −1.0% (95% CI −4.6 to 2.8) per additional 14 high-exposure days. The timing of a malnutrition peak could not be identified: 70% of posterior draws had no interior maximum, and the remaining draws were spread across the whole lag window. These results do not support food-system disruption, as reflected in malnutrition admissions, as the explanation for the TB association. However, admissions capture only malnutrition severe enough to require hospital care funded by the Unified Health System (SUS). Milder undernutrition, which may already impair immunity to *M. tuberculosis*, is not captured. The result should therefore be read as supportive rather than definitive, and nutritional pathways remain a possible source of residual bias (Section S3.17).

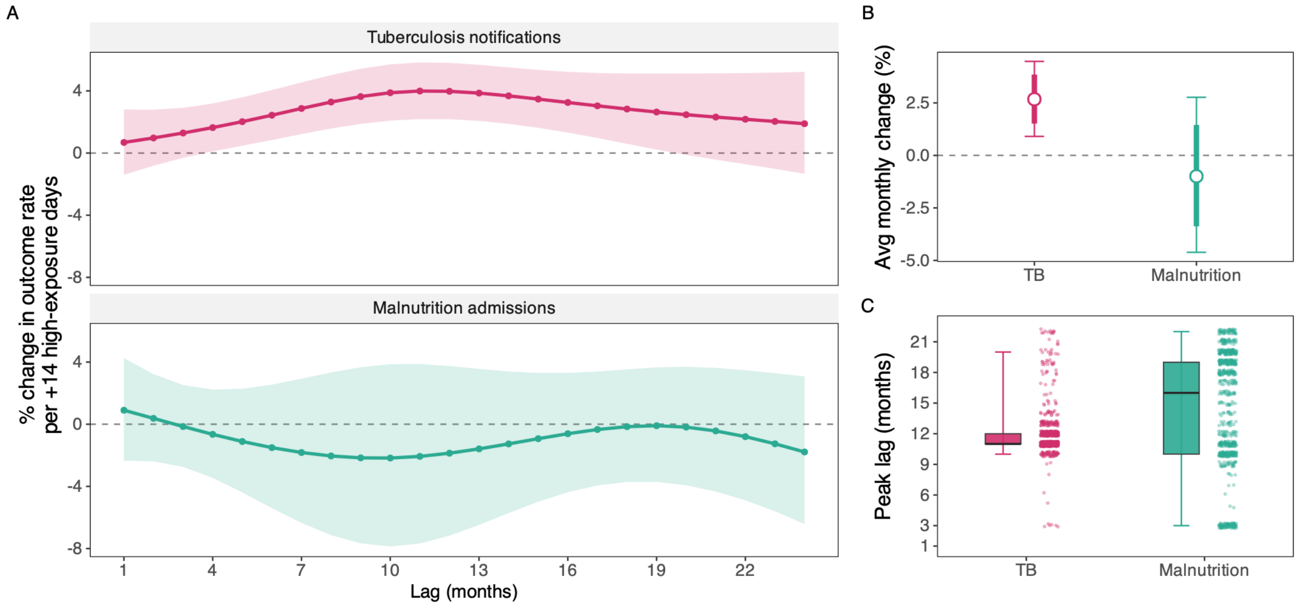


**Supplementary Fig. 41. Malnutrition hospital admissions as an alternative-pathway outcome for the wildfire-related PM2.5 to tuberculosis lag-response.** Penalized-spline distributed-lag model ($PM_{2.5} > 25$ µg/m$^3$ exceedance days, GFED4.1s), Brazil, Jan 1, 2005 – Dec 1, 2023, with municipality, state × year, and state × month fixed effects, a log-population offset, GeneXpert diagnostic coverage, state pandemic phase, temperature and precipitation distributed-lag terms, and Conley spatial HAC standard errors (cutoff 300 km). This sensitivity analysis uses hospital admissions with a principal diagnosis of protein-energy malnutrition (SIH-RD, ICD-10 E40-E46) as the outcome. (A) Lag-response curves for tuberculosis (upper panel) and malnutrition admissions (lower panel). Each line is the mean percent change in that outcome's own rate per 14 additional high-exposure days (days above 25 $\mu g/m^3$) at each lag from 1 to 24 months; shaded bands are 95% Conley confidence intervals. The dashed line indicates no effect. (B) Average monthly effect for each outcome: the geometric mean of the per-month rate ratios across the 1-24-month lag window, expressed as the percent change in the outcome's rate per 14 additional high-exposure days. The white point is the central estimate; thick bars show the 80% interval and thin bars the 95% interval, both from Conley spatial HAC standard errors. The dashed line indicates no effect. (C) Posterior distribution of the peak lag for each outcome, obtained by Monte Carlo sampling (N = 10,000) of the model coefficients, reconstructing the lag-response curve for each draw, and taking the argmax. Draws without an interior peak (non-positive mean over the window) were dropped. Boxplots show the median and interquartile range, with whiskers at the 2.5th and 97.5th percentiles; points are a subsample of retained draws.

#### S3.16 Exclusion of migrant TB cases and border municipalities

Many municipalities with high wildfire-related $PM_{2.5}$ exposure lie along Brazil's international land border. Since 2015, these areas have also received substantial migration, particularly from Venezuela into Roraima.[36] Migration could bias the effect in two ways. First, immigrants may present for diagnosis in Brazil when already symptomatic, after infection and progression elsewhere. Second, people migrating from settings with a higher TB burden could raise local notification rates in years that coincide with severe fire seasons. We conducted three sensitivity analyses that remove migrant cases or high-migration areas directly.

**Approach.** We re-fitted the primary penalized distributed-lag model three times. First, we excluded TB cases recorded as immigrants in SINAN from the outcome, keeping everything else in the analysis unchanged (Section S1.1). Second, we excluded all 15 municipalities of Roraima, the state most affected by the Venezuelan inflow, where immigrants accounted for 18.6% of TB notifications between 2015 and 2023. Third, we excluded the 33 twin cities on the international land border (Section S1.3), where cross-border movement is greatest. The exposure definition, lag basis, fixed effects, covariates, and Conley standard errors were otherwise unchanged.

**Results.** Excluding immigrant cases removed 4,879 notifications (0.29% of the total). Excluding Roraima removed 4,334 notifications (0.26%), and excluding twin cities removed 12,667 (0.76%). None of the exclusions materially changed the lag-response (Supplementary Fig. **43**, panel A). After excluding immigrant cases or Roraima, the curves differed from the main analysis by at most 0.06 percentage points at any lag. The average monthly effect was +2.7% (95% CI 1.0 to 4.5) and +2.7% (95% CI 0.9 to 4.5) per additional 14 high-exposure days, respectively, compared with +2.7% (95% CI 0.9 to 4.5) in the main analysis (panel B). Excluding twin cities slightly attenuated the estimate to +2.4% (95% CI 0.2 to 4.7) and widened the confidence interval. The peak lag remained at 11 months (IQR 11–12) after excluding immigrant cases or Roraima, and was 12 months (IQR 11–13) after excluding twin cities (panel C). Immigrant status is only recorded consistently from 2015, so immigrant cases notified before then could not be removed. However, migration from Venezuela also began around this time. These results suggest that the association is not explained by migrant cases or by migration into border municipalities.

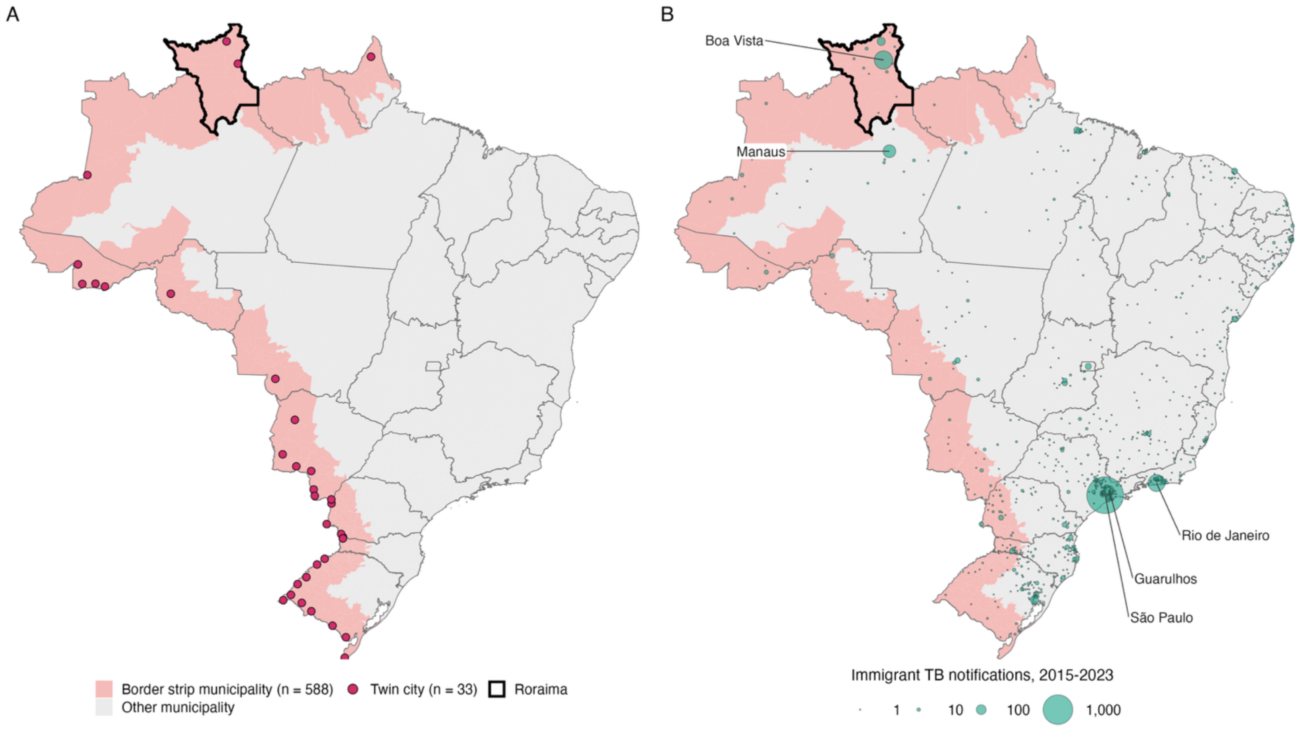


Supplementary Fig. 42. **Border municipalities and immigrant TB notifications in Brazil.** (A) Municipalities in the border strip (faixa de fronteira), the 150 km zone along Brazil's international land border, as listed by IBGE (2022). Points mark the twin cities (cidades gemeas), municipalities on the border that are paired with a town in a neighbouring country; points are placed inside each municipality so that small municipalities remain visible. The thick outline marks the state of Roraima. These are the areas excluded in the migration sensitivity analyses (Section S3.16). (B) TB notifications recorded as immigrant cases in SINAN between Jan 1, 2015 and Dec 1, 2023, by municipality of residence. The size of the bubble is proportional to the number of notifications; municipalities with none are not shown. The 5 municipalities with the most immigrant notifications are labelled. The border strip is shaded as in panel A. State boundaries are shown in grey.

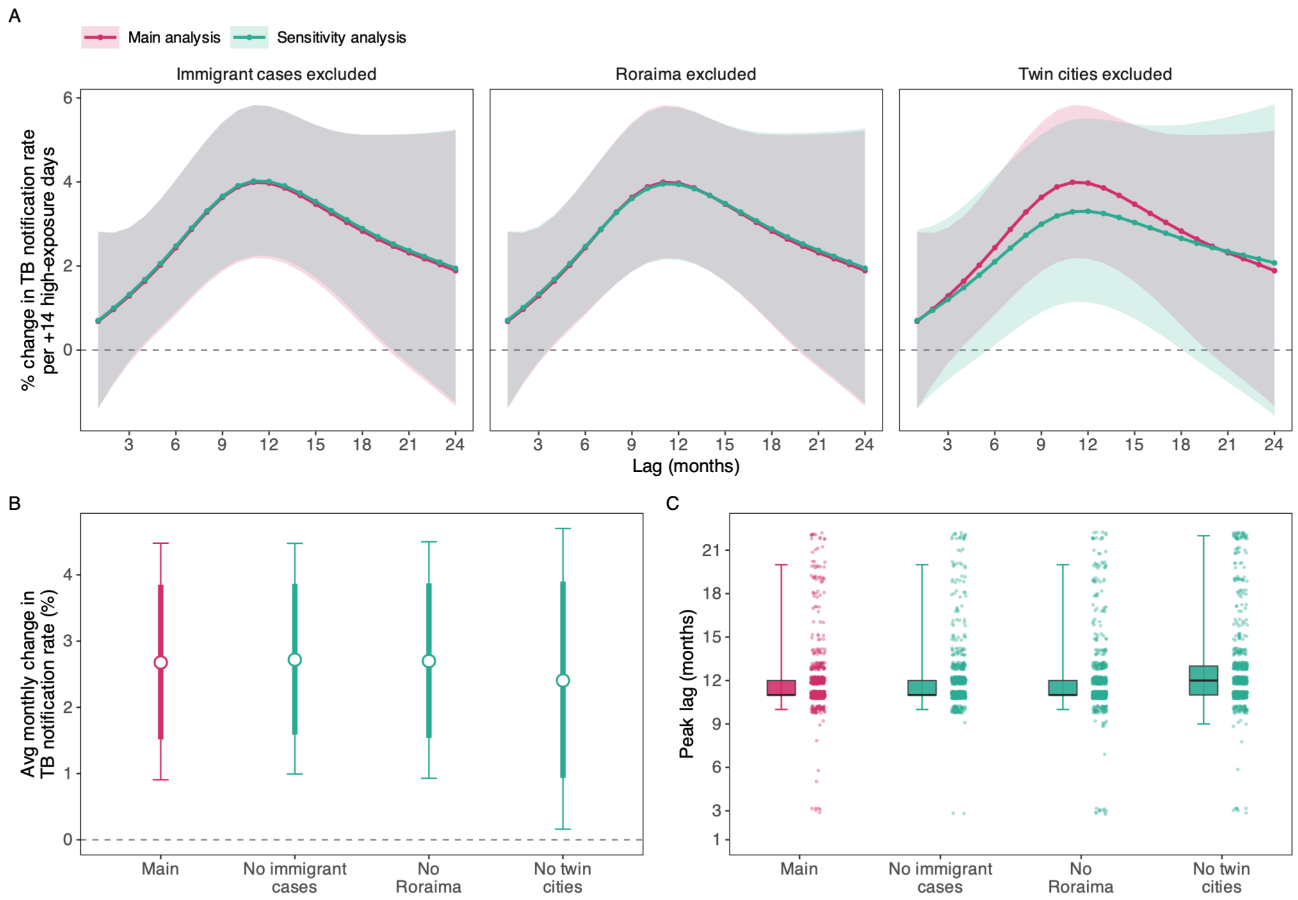


**Supplementary Fig. 43. Robustness of the wildfire-related PM2.5 to tuberculosis lag-response to migration.** Penalized-spline distributed-lag model ($PM_{2.5}$ > 25 µg/$m^3$ exceedance days, GFED4.1s), Brazil, Jan 1, 2005 – Dec 1, 2023, with municipality, state × year, and state × month fixed effects, a log-population offset, GeneXpert diagnostic coverage, state pandemic phase, temperature and precipitation distributed-lag terms, and Conley spatial HAC standard errors (cutoff 300 km). Each sensitivity analysis re-fits the main model with TB cases recorded as immigrants in SINAN removed from the outcome; all municipalities of Roraima removed; the IBGE twin cities on the international land border removed. (A) Lag-response curves for each sensitivity analysis, overlaid on the main analysis. Lines are the mean percent change in the TB notification rate per 14 additional high-exposure days (days above 25 µg/$m^3$) at each lag from 1 to 24 months; shaded bands are 95% Conley confidence intervals. The dashed line indicates no effect. (B) Average monthly effect for each model: the geometric mean of the per-month rate ratios across the 1–24-month lag window, expressed as the percent change in the TB notification rate per 14 additional high-exposure days. The white point is the central estimate; thick bars show the 80% interval and thin bars the 95% interval, both from Conley spatial HAC standard errors. The dashed line indicates no effect. (C) Posterior distribution of the peak lag for each model, obtained by Monte Carlo sampling (N = 10,000) of the model coefficients, reconstructing the lag-response curve for each draw, and taking the argmax. Draws without an interior peak (non-positive mean over the window) were dropped. Boxplots show the median and interquartile range, with whiskers at the 2.5th and 97.5th percentiles; points are a subsample of retained draws.

#### S3.17 Consideration of additional time-varying confounders

Several other time-varying factors could, in principle, confound the effect of wildfire-related $PM_{2.5}$ on TB notifications. Seasonal labour migration along the arc of deforestation may coincide with the dry-season fire period and alter the local distribution of latent TB infection risk; household biomass-fuel use may be higher in rural fire-prone municipalities and could independently increase TB risk; and drought- or fire-related food insecurity could promote progression from latent infection to active disease. For these pathways to confound the relationship each would need to vary within municipalities over time, align with wildfire-related $PM_{2.5}$ at the relevant lag structure, and affect TB notifications independently of the fixed effects and covariates already included. Municipality fixed effects absorb stable rural-urban and household-energy differences, state-by-month and state-by-year fixed effects control broad seasonal and secular patterns, and additional adjustment for drought and agricultural expansion addressed two major time-varying correlates of fire activity. Although residual confounding from these mechanisms cannot be excluded, it would need to be highly structured in space, season, and lag to reproduce the observed delayed lag-response pattern.

#### S4. Supplementary heterogeneity results

This section reports pre-specified secondary analyses of effect modification by age, HIV co-infection status, and urbanization, as well as disaggregated biome and microregion results that complement main-text Figures 3-4.

##### S4.1 Effect modification by age group

**Approach.** To assess effect modification by age, we estimated the wildfire-related $PM_{2.5}$–TB lag-response separately for three age groups: paediatric cases aged 0–14 years, working-age adults aged 15–59 years, and older adults aged 60 years and above. We used a single interaction model fitted across 558 IBGE microregions. Each microregion-month contributed three rows, one for each age group. The model included a separate penalized P-spline lag block for each age group, with $k = 8$ over lags 1–24 months, as well as an age-group main effect. Each age-specific block used its own log population offset, allowing the three curves to be compared directly on the notification-rate scale.

The model included microregion fixed effects, state-by-year, state-by-month, and state-by-pandemic-phase fixed effects, GeneXpert diagnostic coverage. Standard errors were estimated jointly across the three age groups using Conley spatial HAC standard errors with a 300 km cutoff. We used microregions rather than municipalities to stabilize age-specific counts, because paediatric and older-adult TB notifications are sparse in many small municipalities. Heterogeneity in the cumulative effect across age groups was evaluated using an omnibus Wald test, with working-age adults as the reference group, and summarized using Cochran's Q, tau-squared, and I-squared. The age distribution of TB notifications, nationally and by biome, is shown in Supplementary Fig. 44.

**Results.** The association between wildfire-related $PM_{2.5}$ and TB notifications increased with age (Supplementary Fig. 45, panels A-C). The estimated effect was largest among elderly (panel D): 14.5% (95% CI 10.4 to 18.8). The paediatric estimate was also elevated, at 5.6% (95% CI 0.2 to 11.3). In contrast, the working-age estimate was smaller and less certain, at 1.3% (95% CI -1.3 to 3.9). The 95% confidence intervals excluded the null for the older-adult and paediatric groups but included the null for working-age adults.

There was strong evidence of heterogeneity across age groups. The omnibus Wald test was statistically significant (chi-squared = 32.2 on 2 df, $p < 0.0001$), with working-age adults as the reference group. Cochran's Q was 28.6 on 2 df ($p < 0.0001$), and I-squared was 93%, indicating that most of the between-group variation was unlikely to be explained by sampling error alone.

The largest effect in older adults is consistent with a latent-reactivation pathway, because older adults are more likely to carry remotely acquired latent TB infection that could progress following pollutant-related immune disruption. The elevated paediatric estimate is also biologically plausible, given children's higher risk of progression after recent infection. By contrast, the smaller and less certain effect in working-age adults may reflect a larger contribution of recent transmission and other social determinants of TB risk. Overall, these results suggest that the TB burden attributable to wildfire-related $PM_{2.5}$ is concentrated especially among older adults.

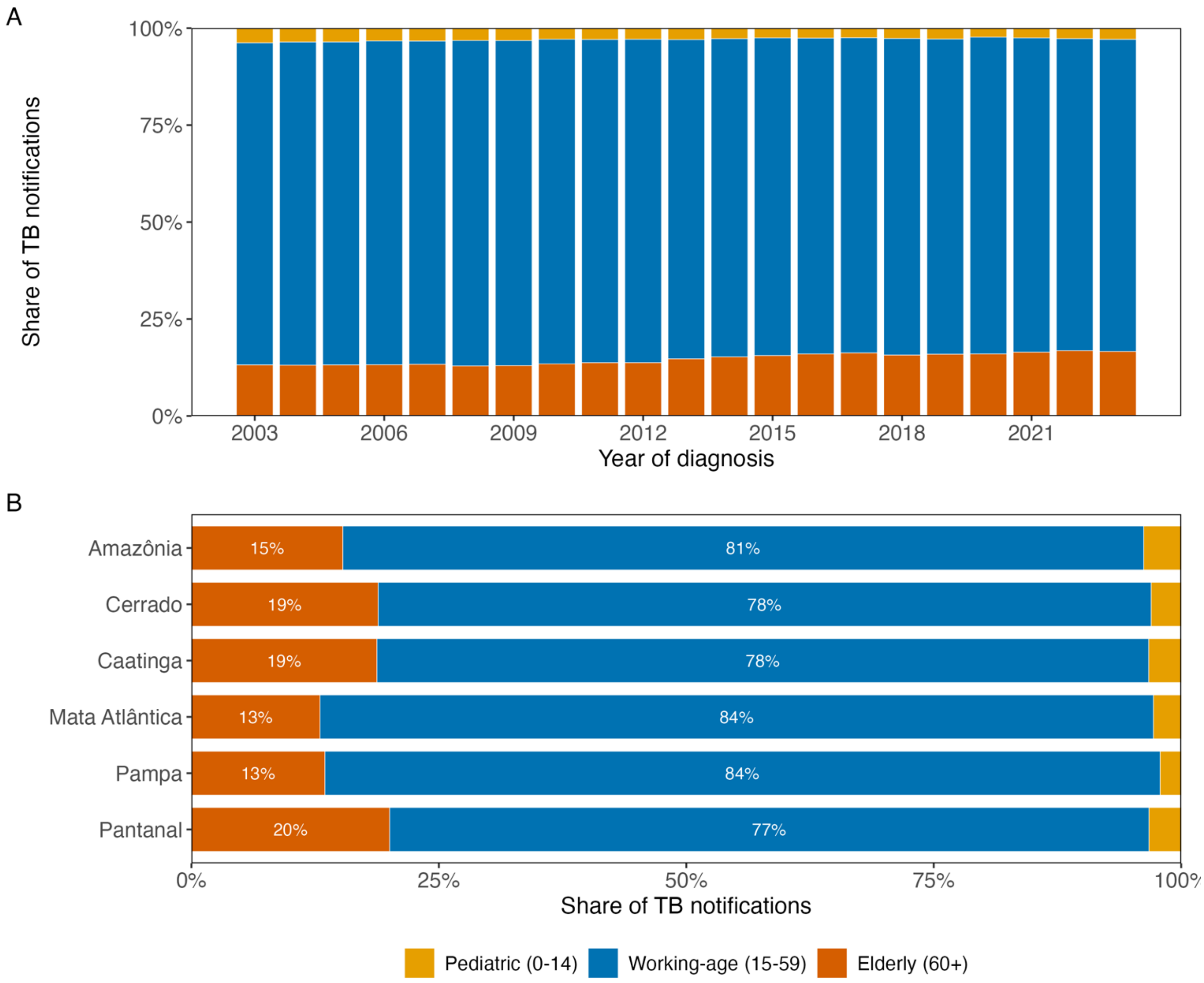


**Supplementary Fig. 44. Age distribution of tuberculosis notifications in Brazil, nationally and by biome.** Bars show the share of TB notifications falling in each of three age groups, Paediatric (0-14), Working-age (15-59) and Elderly (60+), among notifications with a valid recorded age. Counts are from the analysis-ready age-stratified municipality-month panel (GFED4.1s primary-exposure inventory), the same notification counts used in the age effect-modification analysis; age is year of diagnosis minus birth year, with implausible ages (< 0 or > 120 years) set to missing. Period: Jan 1, 2003 – Dec 1, 2023. (A) National age composition by year of diagnosis; each yearly bar sums to 100% of age-known notifications. (B) Age composition within each of Brazil's six biomes, pooled over the study period; municipalities that could not be assigned to a biome are excluded from this panel. Within each biome bar the segment labels give the rounded percentage of notifications in that age group (labels below 5% are suppressed).

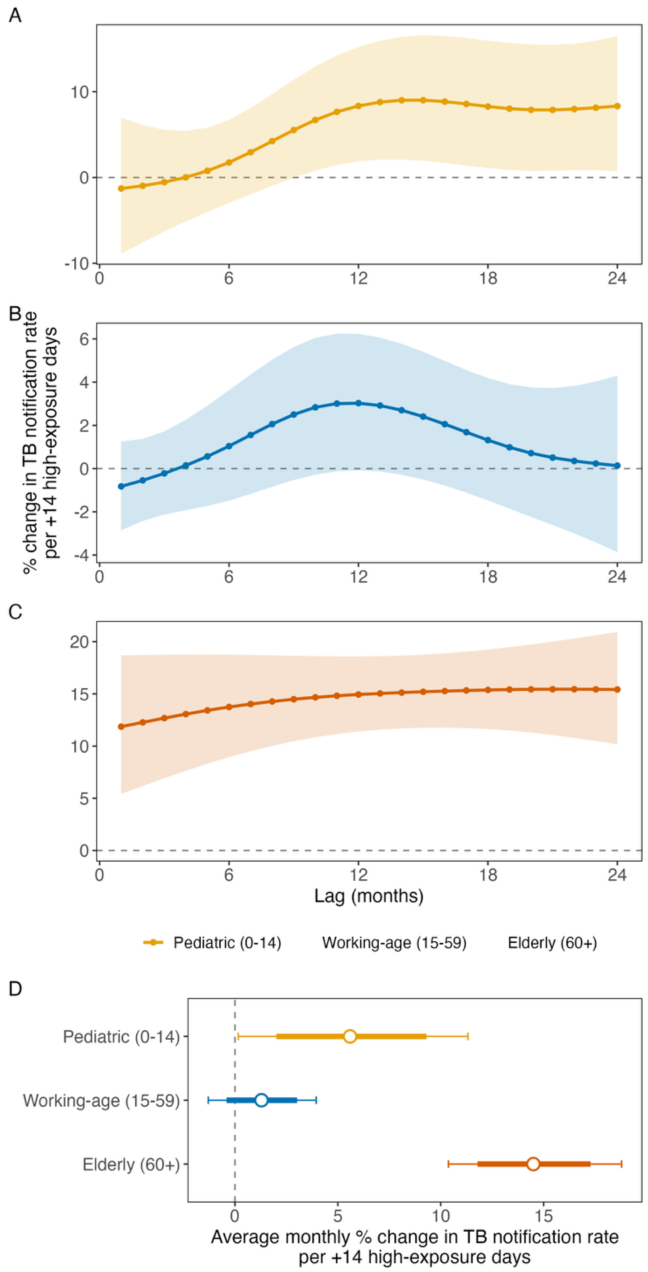


**Supplementary Fig. 45. Age group effect modification analysis of the lag response relationship between wildfire-related $PM_{2.5}$ and TB notification rates, micro-region sensitivity analysis.** Estimates are from a single penalized-spline distributed-lag interaction model (penalized P-spline lag basis, k = 8; lags 1-24 months) fitted across 558 IBGE micro-regions, with one age-specific $PM_{2.5}$ lag block per group plus an age main effect. Exposure is monthly days above 25 µg/m³ of GFED4.1s wildfire-related $PM_{2.5}$. The model includes micro-region, state-by-year and state-by-month fixed effects, a log age-specific population offset, the proportion of GeneXpert diagnoses and a pandemic-phase term; standard errors use a joint Conley spatial HAC (300 km) across the three age blocks. Period: Jan 1, 2005 – Dec 1, 2023. Heterogeneity of the cumulative $PM_{2.5}$ effect across age groups was assessed with an omnibus Wald test (reference: working-age) and summarized with Cochran's Q and I-squared. (A-C) Lag-response curves for (A) Paediatric (0-14), (B) Working-age (15-59) and (C) Elderly (60+). Lines are the mean percent change in TB notification rates per +14 additional high-exposure days at each monthly lag; shaded bands are 95% Conley confidence intervals. y-axes are scaled independently per panel because effect magnitudes differ markedly by age. The dashed line indicates no effect. (D) Average monthly effect per age group on a common axis: the per-month geometric-mean percent change over the 1-24 month window. Points are central estimates; thick bars are 80% and thin bars 95% Conley confidence intervals. The dashed line indicates no effect.

#### S4.2 Effect modification by HIV status

**Approach.** We estimated the primary specification with the exposure cross-basis interacted with HIV status, allowing the full lag–response to differ between HIV-positive and HIV-negative notifications within a single penalized P-spline distributed lag model; cases with missing or unknown HIV status were assigned to the HIV-negative stratum. Heterogeneity was assessed by a joint Wald test of the exposure × HIV-status interaction terms (df equal to the number of cross-basis functions).

**Results.** Over the whole study period there were 183,761 (10.4%) confirmed HIV-positive, 958,113 (54.5%) confirmed HIV-negative, and 617,108 (35.1%) TB cases with unknown/missing HIV status. The exposure × HIV-status interaction was significant (p=0.033), indicating that the lag–response differed between strata (Supplementary Fig. 46). HIV-positive notifications represent a small minority of the total, so the HIV-positive estimates carry substantially wider CIs (Supplementary Fig. 47).

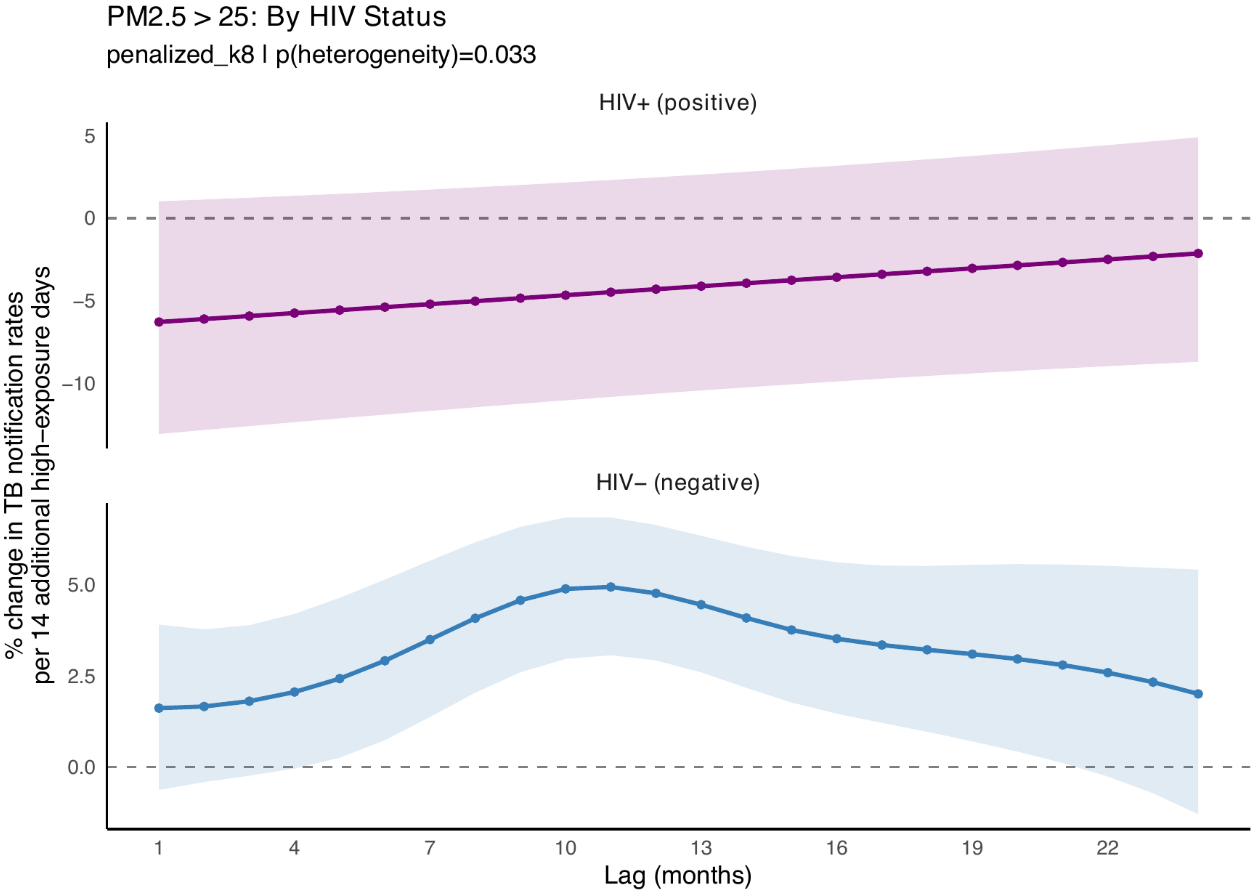


**Supplementary Fig. 46. HIV-status-specific lag-response curves for wildfire-related $PM_{2.5}$ exposure and TB notification rates, from a penalized pooled interaction model.** Curves show the modelled percentage change in TB notification rates per 14 additional high-exposure days across lags of 1–24 months, for HIV-positive (upper panel) and HIV-negative (lower panel) notifications. Points are lag-specific estimates and shaded bands the corresponding 95% CIs; the dashed horizontal line denotes no change.

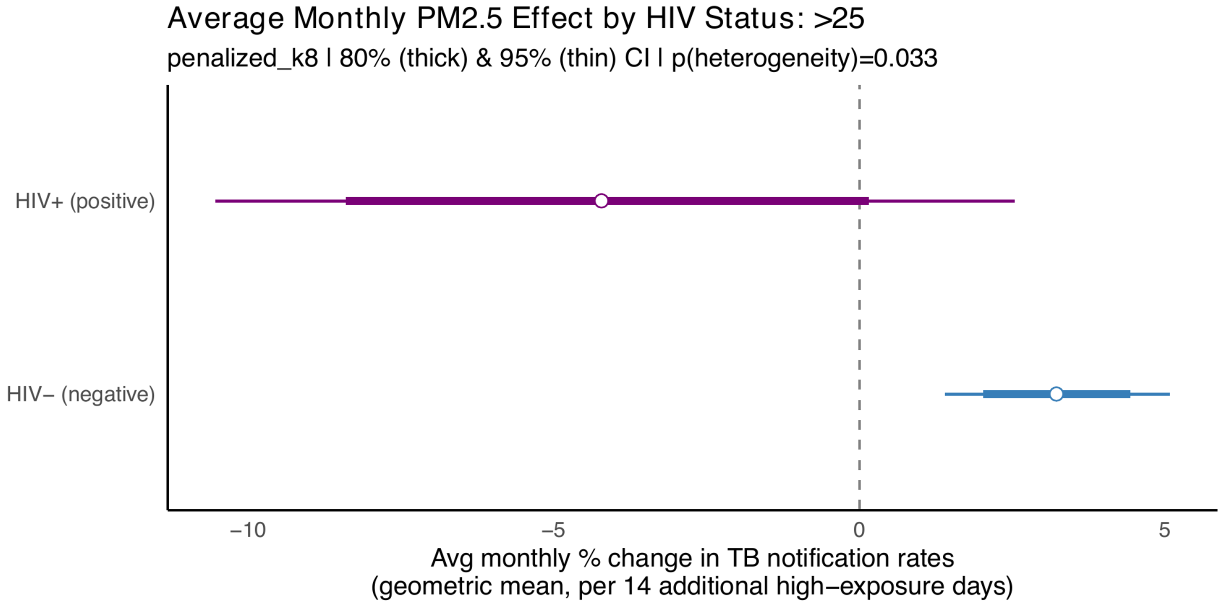


**Supplementary Fig. 47. Lag-averaged effect of wildfire-related $PM_{2.5}$ on TB notification rates by HIV status, from the penalized pooled interaction model.** Points are the geometric-mean monthly percentage change per 14 additional high-exposure days across lags of 1–24 months, with 80% (thick) and 95% (thin) intervals. Upper row, HIV-positive; lower row, HIV-negative; the dashed line denotes no change.

##### S4.3 Effect modification by biological sex

**Approach.** To assess whether the lag-response relationship between wildfire-related $PM_{2.5}$ and TB notification rates differs between men and women, we refitted the primary penalized-spline distributed-lag model separately for male and female TB notifications, holding the primary model structure fixed. Each stratum used a log sex-specific population offset interpolated between the 2010 and 2022 censuses, so the two curves are directly comparable on the notification-rate scale; notifications with missing or indeterminate recorded sex were excluded from the sex-specific counts. Because the strata are estimated as two independent fits rather than a single interaction model, this is a descriptive comparison, and we report no formal interaction test.

**Results.** The lag-response curves were closely aligned in shape and magnitude for men and women (Supplementary Fig. 48), with both following the delayed unimodal pattern observed in the pooled analysis. The average monthly effect was 2.9% (95% CI 0.2 to 5.8) for men and 3.1% (95% CI 0.7 to 5.5) for women, with widely overlapping confidence intervals. The posterior peak-lag distributions were also similar, centred at 13 months for men and 11 months for women, consistent with with the primary analysis. Overall, these results provide no evidence that the association between wildfire-related $PM_{2.5}$ exposure and TB notifications is modified by sex: the magnitude, lag structure, and timing of the peak effect were essentially the same in men and women. These findings support pooling sexes in the primary analysis and are consistent with an exposure-driven mechanism that does not depend on biological sex differences in baseline TB incidence in Brazil.

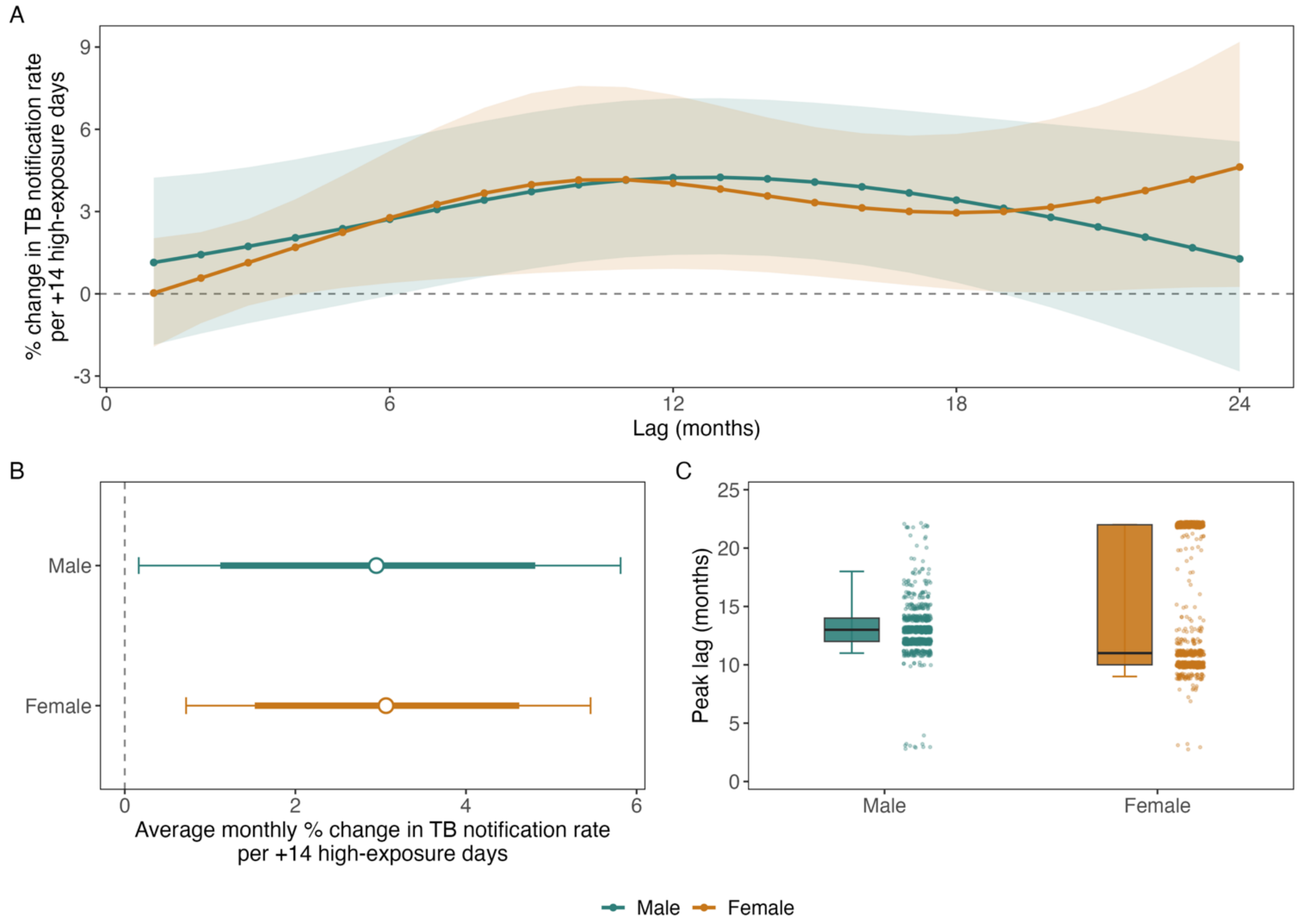


**Supplementary Fig. 48. Biological sex effect modification analysis of the lag-response relationship between wildfire-related $PM_{2.5}$ and tuberculosis notification rates.** Estimates are from two penalized-spline distributed-lag models fitted separately to male and female TB notifications (penalized P-spline lag basis, k = 8; lags 1-24 months). Exposure is the monthly number of days above 25 µg/m³ of GFED4.1s wildfire-related $PM_{2.5}$. Each model includes municipality, state-by-year and state-by-month fixed effects, a log sex-specific population offset and the proportion of GeneXpert diagnoses; standard errors use a Conley spatial HAC (300 km). Cases with missing or indeterminate sex are excluded from the sex-specific counts. Period: Jan 1, 2005 – Dec 1, 2023. (A) Lag-response curves for male (teal) and female (orange) TB notifications on a common axis. Lines are the mean percent change in the TB notification rate per +14 additional high-exposure days at each monthly lag; shaded bands are 95% Conley confidence intervals. The dashed line indicates no effect. (B) Average monthly effect per sex on a common axis: the per-month geometric-mean percent change over the 1-24 month window. Points are central estimates; thick bars are 80% and thin bars 95% Conley confidence intervals. The dashed line indicates no effect. (C) Posterior distribution of the peak lag (the lag at which the lag-response is maximal) per sex. For each sex, 10,000 coefficient vectors were drawn from the Bayesian posterior of the fit and mapped to a lag-response curve via the lag basis; the peak lag of each draw is its argmax. Each half-boxplot shows the 25th, 50th (thick line) and 75th percentiles (box) with whiskers at the 2.5th and 97.5th percentiles; jittered points to the right show a subsample of the individual posterior draws.

### S5. Model diagnostics

#### S5.1 QQ and residual-vs-predicted diagnostics

We assessed model fit using randomized-quantile scaled residuals, which should follow a uniform distribution when the fitted Poisson mean and response distribution are well specified. The QQ plot compares these residuals with their expected uniform quantiles, while the residual-versus-predicted plot assesses whether residuals remain stable across fitted values.

The residuals closely followed the uniform diagonal, and their quartiles were essentially flat across the predicted range, indicating good calibration of the Poisson distributed-lag model (Supplementary Fig. 49). The dispersion ratio was 1.13 and the zero-count ratio was 1.01, suggesting only mild overdispersion and no meaningful zero-inflation. Although formal tests were statistically significant, this likely reflects the very large number of municipality-months. Any residual overdispersion would mainly affect model-based standard errors and is accommodated by the Conley robust standard errors used for inference.

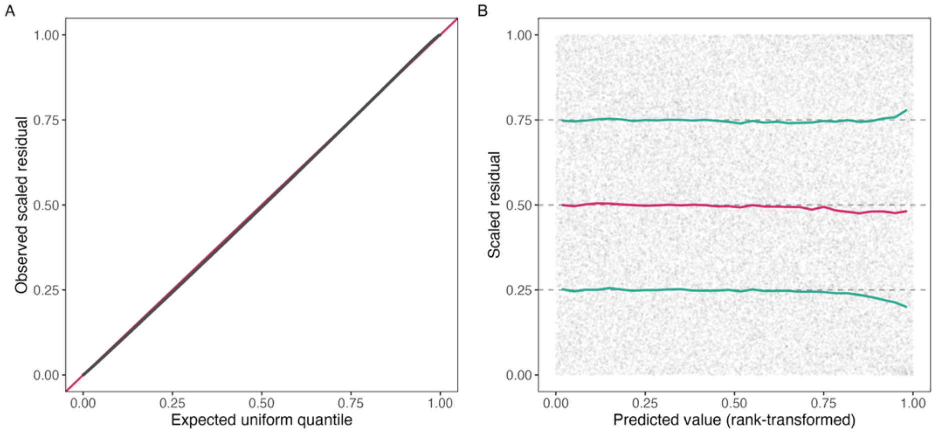


**Supplementary Fig. 49. Distributional diagnostics for the primary penalized distributed-lag model.** (A) Quantile-quantile plot of the randomized-quantile (scaled) residuals against the uniform reference distribution; points lying along the diagonal indicate agreement between the observed and modelled count distributions. (B) Scaled residuals against the rank-transformed predicted value, with the empirical lower-quartile, median, and upper-quartile curves overlaid; horizontal reference lines mark the quartiles expected under correct specification, so flat overlaid curves indicate stable dispersion and no residual trend across the fitted range. All diagnostics are evaluated on the primary analysis: a penalized-spline distributed-lag model of monthly tuberculosis notification counts across Brazilian municipalities (Jan 1, 2005 – Dec 1, 2023), with wildfire-related $PM_{2.5}$ entered as the monthly count of days exceeding 25 μg/m³ (GFED4.1s) over a 1-24 month lag window, with fixed effects for municipality, state-by-year, state-by-month, and state-by-pandemic-phase, a log-population offset, and post-hoc Conley spatial heteroskedasticity- and autocorrelation-consistent standard errors.

#### S5.2 Basis adequacy and smoothing parameter

We assessed whether the distributed-lag spline had adequate flexibility by comparing its effective degrees of freedom after penalization with the maximum allowed by the basis dimension. Values near the maximum would suggest that the basis was too restrictive, whereas values well below the maximum indicate sufficient flexibility. We also examined the estimated smoothing parameter on a log scale to check whether the penalty was at an extreme.

The distributed-lag term retained 4.3 effective degrees of freedom out of a possible 8, indicating adequate headroom and no evidence that the fitted lag shape was constrained by the basis ceiling (Supplementary Fig. 50A). The estimated smoothing parameter was also away from boundary values ($\log10\ \lambda \approx 6.8$), suggesting that the term was neither over-shrunk nor effectively unpenalized (Supplementary Fig. 50B). These diagnostics support the unimodal lag-response as a data-driven feature rather than an artefact of basis size or penalization.

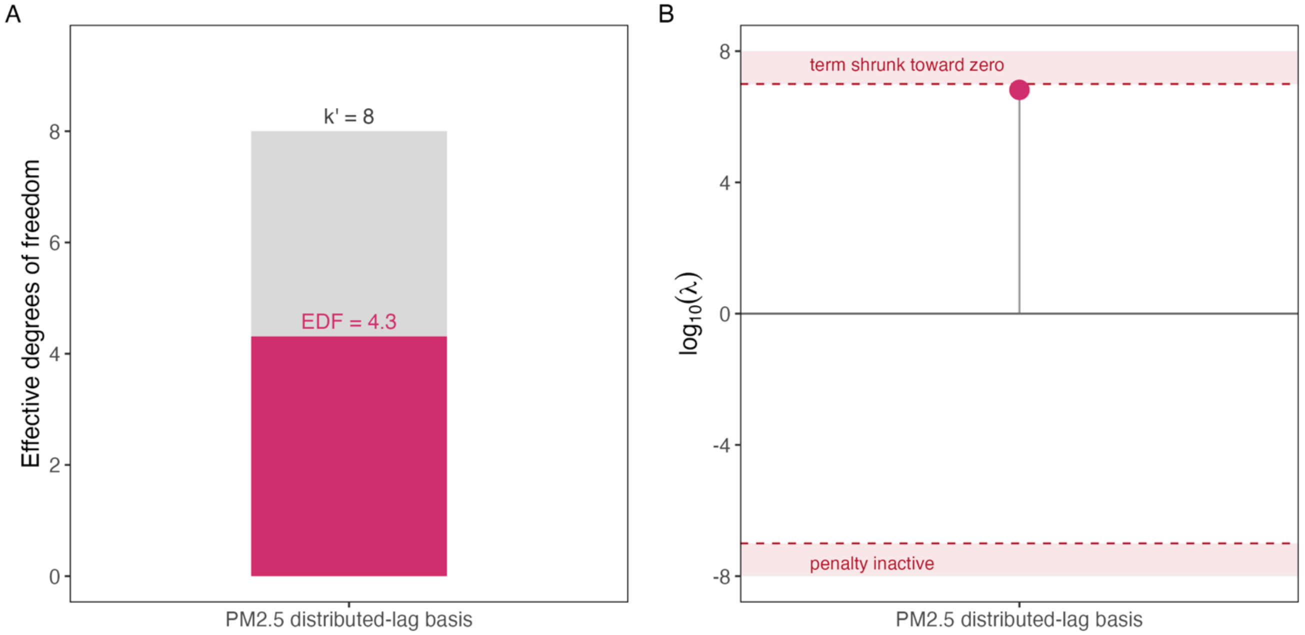


**Supplementary Fig. 50. Basis adequacy and smoothing-parameter diagnostics for the distributed-lag term.** (A) Effective degrees of freedom retained after penalization (solid bar) relative to the basis dimension (shaded bar), which is the maximum flexibility the basis allows. (B) Smoothing parameter on a base-ten logarithmic scale; the shaded bands and dashed lines mark the boundary region within which a penalty would be considered pathological, either shrinking the term toward zero or leaving it effectively unpenalized. All diagnostics are evaluated on the primary analysis: a penalized-spline distributed-lag model of monthly tuberculosis notification counts across Brazilian municipalities (Jan 1, 2005 – Dec 1, 2023), with wildfire-related $PM_{2.5}$ entered as the monthly count of days exceeding 25 µg/m³ (GFED4.1s) over a 1-24 month lag window, with fixed effects for municipality, state-by-year, state-by-month, state-by-pandemic-phase, a log-population offset, and post-hoc Conley spatial heteroskedasticity- and autocorrelation-consistent standard errors.

#### S5.3 Residual Temporal Autocorrelation

We assessed residual temporal dependence using the autocorrelation of the monthly cross-municipality mean residual, compared with a posterior-predictive envelope that accounts for uncertainty in the distributed-lag coefficients. We also repeated this check excluding 2020–2021, when pandemic-related disruptions may have affected notification patterns, and summarized within-municipality serial correlation using a pooled AR(1) estimate.

Most autocorrelations of the monthly mean residual fell within the posterior-predictive envelope, although 12 of 36 lags were outside it, indicating some residual seasonality beyond the seasonal fixed effects (Supplementary Fig. 51). Excluding 2020–2021 produced a similar pattern, with 8 of 36 lags outside the envelope, suggesting this was not driven solely by the pandemic. Within-municipality serial correlation was positive but very small (AR[1] = 0.027). Residual temporal dependence is therefore present but modest, and is addressed by the Conley variance estimator; it affects standard errors rather than the lag-response point estimates.

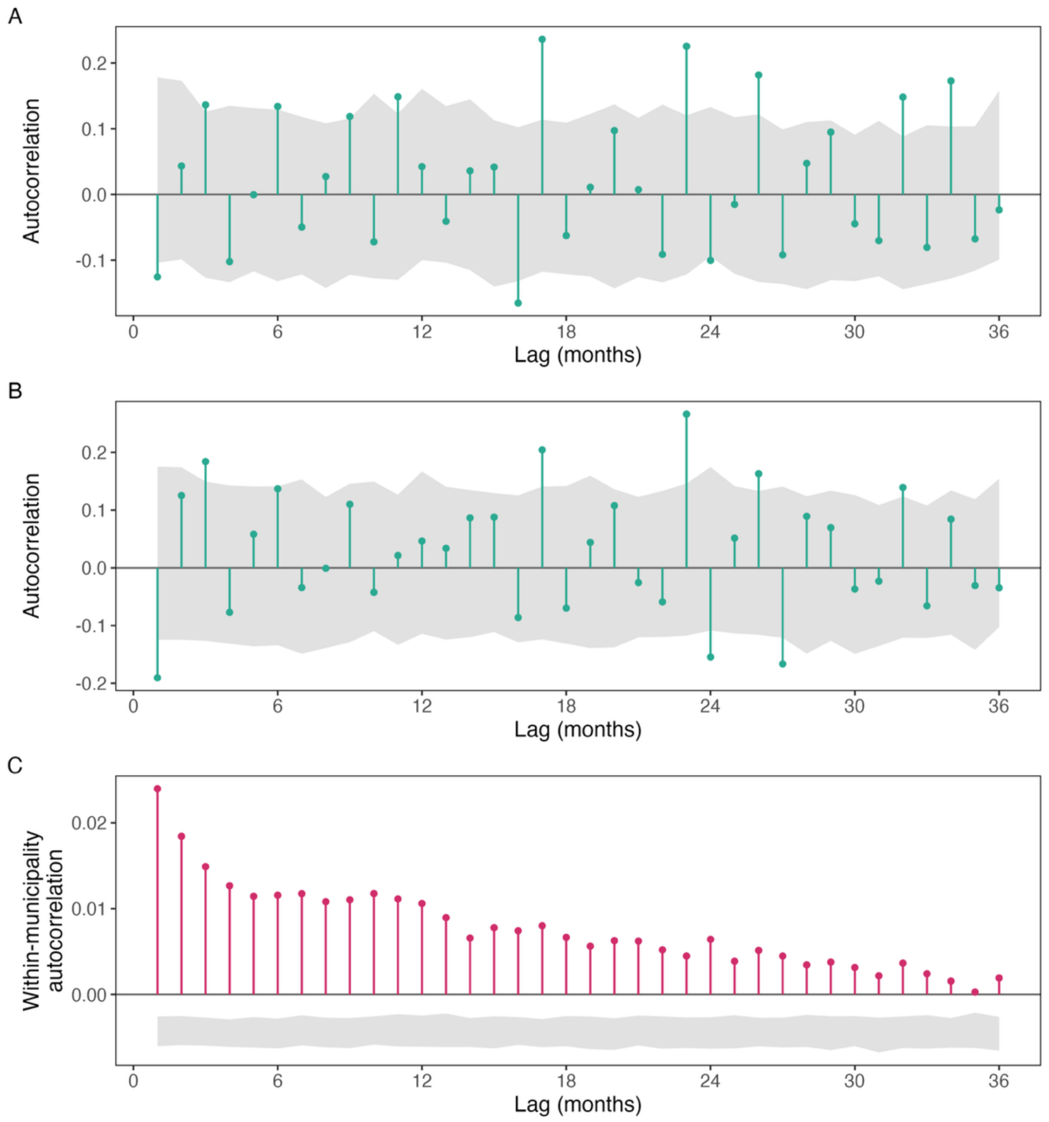


**Supplementary Fig. 51. Temporal autocorrelation of model residuals.** (A) Autocorrelation of the monthly cross-municipality mean scaled residual over the full study period, with the shaded band showing the posterior-predictive envelope constructed by drawing distributed-lag coefficients from their posterior, perturbing the fitted mean, and re-deriving the autocorrelation. (B) The same diagnostic with the 2020-2021 pandemic period excluded, isolating the pandemic notification disruption from seasonal-structure misfit. (C) Pooled within-municipality autocorrelation of the residual series, with its corresponding posterior-predictive envelope. In all panels the horizontal line marks zero autocorrelation and the horizontal axis is the lag in months. All diagnostics are evaluated on the primary analysis: a penalized-spline distributed-lag model of monthly tuberculosis notification counts across Brazilian municipalities (Jan 1, 2005 – Dec 1, 2023), with wildfire-related $PM_{2.5}$ entered as the monthly count of days exceeding 25 µg/m³ (GFED4.1s) over a 1-24 month lag window, with fixed effects for municipality, state-by-year, state-by-month, and state-by-pandemic-phase, a log-population offset, and post-hoc Conley spatial heteroskedasticity- and autocorrelation-consistent standard errors.

#### S5.4 Spatial correlation residuals

We assessed residual spatial dependence using Moran's I for municipality-mean scaled residuals. A distance-band correlogram summarized how spatial correlation changed with separation, while cumulative Moran's I over increasing-radius discs assessed how much correlation was captured within the Conley neighbourhood. Permutation tests were corrected across distance bands using the Holm procedure.

Residual spatial autocorrelation was weak and short-ranged. Moran's I was largest in the 0–100 km band (0.010) and declined to below 0.003 within a few hundred kilometres (Supplementary Fig. 52). After Holm correction, significant autocorrelation was confined to distances within 200 km, well inside the primary 300 km Conley cutoff. The cumulative Moran's I similarly declined as the radius widened, from 0.007 within 300 km to 0.002 within 1000 km. These diagnostics support the use of Conley spatial standard errors with a 300 km primary cutoff.

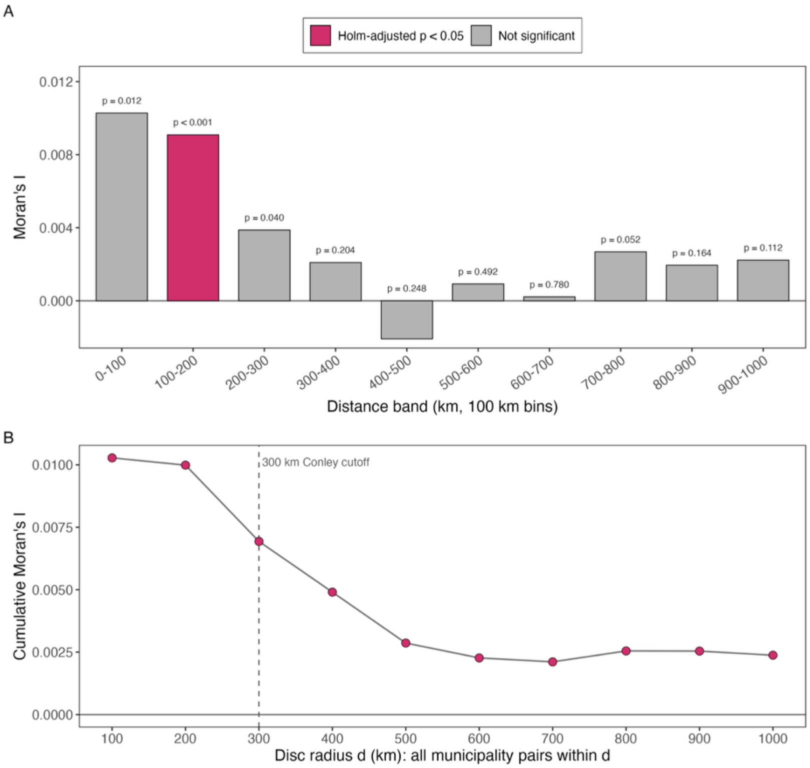


**Supplementary Fig. 52. Spatial autocorrelation of model residuals (Moran's I).** (A) Moran's I correlogram of the municipality-mean scaled residuals over uniform-width distance bands, with bars coloured by Holm-adjusted family-wise permutation significance and labelled with the raw permutation p-value; positive values denote spatial clustering of residuals at that separation. (B) Cumulative Moran's I computed over discs of increasing radius (all municipality pairs within the radius), matching the geometry of the Conley variance estimator, with points coloured by Holm-adjusted significance; the dashed vertical line marks the primary Conley cutoff. All diagnostics are evaluated on the primary analysis: a penalized-spline distributed-lag model of monthly tuberculosis notification counts across Brazilian municipalities (Jan 1, 2005 – Dec 1, 2023), with wildfire-related $PM_{2.5}$ entered as the monthly count of days exceeding 25 µg/m³ (GFED4.1s) over a 1-24 month lag window, with fixed effects for municipality, state-by-year, state-by-month, and state-by-pandemic-phase, a log-population offset, and post-hoc Conley spatial heteroskedasticity- and autocorrelation-consistent standard errors.

#### S5.5 Residual calibration against population

Because the model uses log population as a fixed-coefficient offset, we assessed whether residual calibration varied with municipality size. Municipality-mean scaled residuals were plotted against log population, with linear and locally smoothed fits; a flat relationship would support the proportional-rate assumption implied by the offset.

Mean scaled residuals were centred near 0.50 across the population range, indicating good overall calibration (Supplementary Fig. 53). A weak but statistically detectable trend with population was observed (slope −0.005

per tenfold population increase, $p < 0.001$), corresponding to only about a 0.02 change across the full population range and explaining roughly 2% of between-municipality variation. The largest municipalities showed slight over-prediction, but the magnitude was small and did not indicate a material violation of the log-population offset assumption.

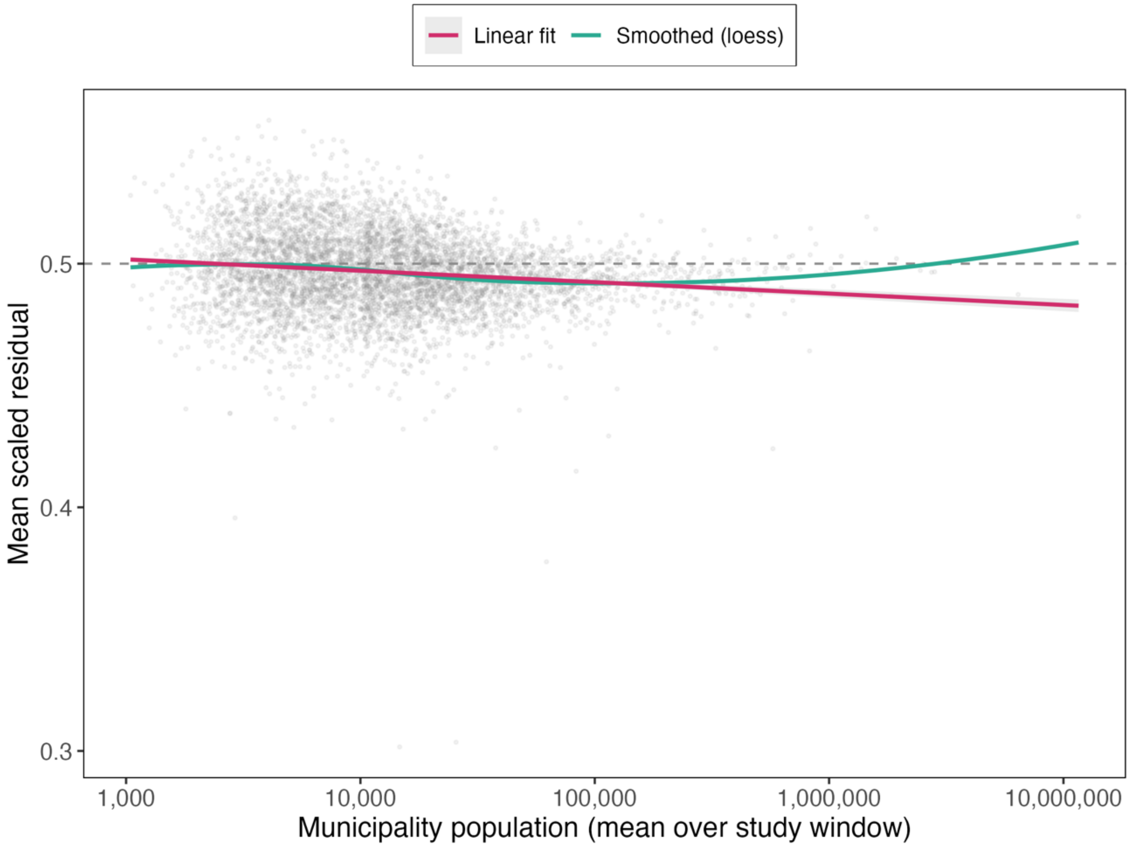


**Supplementary Fig. 53. Residual calibration against municipality population.** The horizontal axis is the municipality mean population over the study window on a logarithmic scale and the vertical axis is the per-municipality mean scaled residual; each point is one municipality. The dashed horizontal line marks the value expected under correct specification, and the overlaid linear and locally smoothed fits summarize any dependence of calibration on population size. All diagnostics are evaluated on the primary analysis: a penalized-spline distributed-lag model of monthly tuberculosis notification counts across Brazilian municipalities (Jan 1, 2005 – Dec 1, 2023), with wildfire-related $PM_{2.5}$ entered as the monthly count of days exceeding 25 μg/m³ (GFED4.1s) over a 1-24 month lag window, fixed effects for municipality, state-by-year, state-by-month, and state-by-pandemic-phase, a log-population offset, and post-hoc Conley spatial heteroskedasticity- and autocorrelation-consistent standard errors.

#### S6. Reporting and reproducibility

##### S6.1 Data and code availability details

Wildfire-related $PM_{2.5}$ data are publicly available from Zenodo:

- GFED4.1s at https://doi.org/10.5281/zenodo.15493914 and
- $QFED_{2.5}$ at https://doi.org/10.5281/zenodo.15496596.

The original dataset is described in Hu and colleagues.[4]

SINAN TB notification data are publicly available from the Brazilian Ministry of Health: https://portalsinan.saude.gov.br. IBGE population estimates and microregion/biome boundaries are publicly available from https://www.ibge.gov.br. ERA5-Land reanalysis data are available from the Copernicus Climate Data Store. CAMS EAC4 NO2 data are available from the Copernicus Atmosphere Data Store. MapBiomas Collection 10 data are available from https://brasil.mapbiomas.org.

All code for data processing and statistical analyses is available at: https://github.com/tm-pham/tb-wildfire-pm25.git. Analyses were run locally on a 14-inch MacBook Pro (Apple M4 Pro, 48 GB RAM, macOS Tahoe 26.5) and on Linux (Ubuntu 22.04) nodes of the University of Utah Center for High-Performance Computing cluster.

##### S6.2 Software versions

| Software / package | Version | Purpose |
|---|---|---|
| R | 4.4.1 | Statistical computing environment |
| fixest | 0.12.1 | Poisson PML with high-dimensional fixed effects; Conley HAC standard errors |
| mgcv | 1.9-1 | Penalized P-spline DLM via bam with paraPen and fREML |
| dlnm | 2.4.7 | Cross-basis construction for the concentration-based DLNM sensitivity |
| geobr | 1.9.1 | IBGE municipality, microregion, and biome boundaries |
| exactextractr | 0.10.0 | Area-weighted zonal statistics for gridded-to-polygon assignment |
| sf | 1.0-16 | Vector spatial operations |
| spdep | 1.3-5 | Neighbour structures for wind-weighted spillover analysis |
| data.table | 1.15.4 | Fast in-memory tabular operations |
| ggplot2 | 3.5.1 | Manuscript figure generation |
| metafor | 4.6-0 | Forest-plot computations for heterogeneity panels |
| terra | 1.7-78 | Raster operations on wildfire $PM_{2.5}$ NetCDF files |
